# Cost-Effectiveness of Vaccination Strategies to Control Future Mpox Outbreaks in England

**DOI:** 10.1101/2024.08.20.24312301

**Authors:** Xu-Sheng Zhang, Siwaporn Niyomsri, Sema Mandal, Hamish Mohammed, Miranda Mindlin, Bennet Dugbazah, Solomon Adjei, Andre Charlett, Jessica Edney, Elliot Sugars, Merav Kliner, Trish Mannes, Ellie Jewitt, Lorna Gilbert, Samihah Moazam, Claire Dewsnap, David Phillips, Gayatri Amirthalingam, Mary E. Ramsay, Peter Vickerman, Josephine Walker

## Abstract

**Background:** In 2022, a global outbreak of mpox occurred among gay and bisexual men who have sex with men (GBMSM). In England, the outbreak was controlled through reductions in sexual risk behaviour and vaccination of high-risk GBMSM. However, mpox continues to circulate and so future outbreaks could occur. We evaluated the most cost-effective vaccination strategy to minimise future mpox outbreaks among GBMSM in England.

**Methods:** A mathematical model of mpox transmission among GBMSM was developed to estimate the costs per quality-adjusted-life-year (QALY) gained for different vaccination strategies starting in 2024 (20-year time-horizon; 3.5% discount rate; willingness-to-pay threshold £20,000/QALY). The model was calibrated using English surveillance data from the 2022 outbreak and two community surveys. Reactive vaccination (only during outbreaks) and pre-emptive vaccination (continuous routine) strategies targeting high-risk GBMSM were compared to no vaccination. Baseline projections assumed vaccine effectiveness of 78%/89% for 5/10 years with 1/2 doses at £160/dose. Costs were estimated for case management, vaccination and public health responses (PHR) during an outbreak.

**Findings:** All vaccination strategies reduced costs and gained QALYs compared to no vaccination. Continuous pre-emptive vaccination (daily rate 41 doses) was most cost-effective, saving £39.56 million and gaining 547.6 QALYs over 20-years. Threshold analyses suggested vaccination of high-risk GBMSM is cost-effective if the vaccine costs <£701/dose. Pre-emptive vaccination remains the optimal strategy across numerous sensitivity analyses, but the optimal vaccination rate can vary. Reactive vaccination only becomes more cost-effective when PHR costs are not included.

**Interpretation:** Pre-emptive vaccination of high-risk GBMSM is a cost-saving strategy to prevent future mpox outbreaks.

**Funding:** NIHR

**Extended funding statement:** This study was funded by the NIHR Health Protection Research Unit in Behavioural Science and Evaluation at University of Bristol NIHR200877, in partnership with UK Health Security Agency (UKHSA). The views expressed are those of the author and not necessarily those of the NIHR, the Department of Health and Social Care, or UKHSA.

**Research in context:** *Evidence before this study:* The global outbreak of mpox in 2022 predominantly affected gay, bisexual, and other men who have sex with men (GBMSM). After a steep rise in cases over May to June 2022, the rate of cases of mpox decreased dramatically after July 2022, thought to be due to the roll-out of vaccination programmes in many countries and reductions in sexual risk behaviour among GBMSM. Despite this decline in cases, new infections of mpox have occurred among GBMSM in many countries in 2023, raising concerns that new outbreaks could occur especially if levels of vaccine-induced protection reduce over time. We searched PubMed, bioRxiv and medRxiv for articles published from beginning May 2022 to 28 June 2024 with the following keywords: ((“monkeypox” OR “mpox” OR “mpx”) AND (“model” OR “modelling” OR “modeling”) AND (“vaccine” OR “vaccination” OR “cost-effectiveness” OR “cost-effective”)). Although this search identified many articles involving transmission modelling that assessed the impact of various interventions on mpox transmission, only eight provided insights on what is needed to prevent future outbreaks, just one considered the cost implications of vaccinating for mpox, and none evaluated the cost-effectiveness of vaccination. Existing model analyses have evaluated what interventions are needed to control outbreaks showing that future outbreaks could be controlled by vaccinating close contacts of cases and individuals in large sexual networks, as well as pre-emptively vaccinating high-risk individuals prior to outbreaks occurring. None of these analyses used detailed data to calibrate their models to actual settings, reducing their real-world relevance. Conversely, other model analyses undertook detailed modelling for specific settings (Canada, Netherlands and England), and showed that existing levels of vaccine roll-out may have reduced the magnitude of future outbreaks. However, these analyses did not model possible future vaccination strategies. The only economic analysis for mpox compared the costs of vaccination to not vaccinating the general population in Jeddah, Saudi Arabia, suggesting that vaccination costs more than not vaccinating, although vaccinating the general population is an unrealistic strategy. Unfortunately, this economic analysis used implausible data (respiratory infection contact rates) to simulate the transmission of mpox, did not use recent data to estimate transmissibility, did not focus on GBMSM, and used very little data on the health-related costs of mpox disease.

*Added value of this study:* This economic analysis extends our understanding of what is needed to control future outbreaks of mpox among GBMSM in England and other settings. Combining a previously validated model of mpox infection in England with real data on the costs of care for mpox, vaccination and public health responses, we undertook an economic analysis to evaluate the most cost-effective future vaccination strategy to prevent future mpox outbreaks. We model either reactive (only vaccinate during outbreaks) or pre-emptive (routine vaccination irrespective of outbreaks) vaccination strategies targeting high-risk GBMSM. Our analyses show that all modelled vaccination strategies are likely to be cost-saving and improve quality of life compared to not vaccinating, with continuous pre-emptive vaccination at a low rate (daily rate 41 doses) being the most cost-effective strategy. This finding is robust over most sensitivity analyses with mpox vaccination remaining cost-effective if the vaccine price is less than £701 per dose.

*Implications of all the available evidence:* Ongoing importation of new sexually transmitted mpox cases in many non-endemic countries means that these countries need to be prepared for future mpox outbreaks if immunity levels fall or if the pool of unvaccinated people increases to a large extent. Our analyses give robust evidence that mpox vaccination is a cost-saving strategy for minimizing the likelihood of future mpox outbreaks in England and other comparable countries. These findings have been used as evidence by the UK Joint Committee on Vaccination and Immunisation to recommend a pre-emptive (routine) vaccination programme of high-risk GBMSM through sexual health services in the UK. Other countries should seriously consider similar strategies to prevent future outbreaks.

## Introduction

In May 2022, a global outbreak of mpox was detected in Western Europe and other non-endemic countries that primarily affected gay, bisexual, and other men who have sex with men (GBMSM) through their sexual networks(1). On 23 July 2022, the World Health Organization (WHO) declared a multi-country mpox outbreak. In response, many nations implemented measures to decrease the number of cases, including provision of vaccines and public health measures, such as isolation of cases, contact tracing and awareness raising(2). The global outbreak declined after July 2022, with 91,788 confirmed cases from 116 countries by October 2023(3). In England, cases peaked in mid-July (>60 cases/day) and then declined, with 3,412 reported cases by 16 September 2022, two thirds of which were in London(4).

The Modified Vaccinia Ankara (MVA) vaccine, originally developed for smallpox, was rolled out in England and other countries during the 2022 outbreak to reduce mpox transmission. Studies undertaken following this roll-out estimated that the vaccine offered ∼85% efficacy against mpox(5–7). However, although immunity can persist for decades following live smallpox vaccination(8), the duration of vaccine-induced protection for the non-replicating MVA vaccine is uncertain. Despite the protection provided by vaccination, model analyses suggest that the vaccines delivered to GBMSM at high-risk of mpox in England had limited impact on the mpox outbreak due to their late roll-out (late June 2022 onwards), but probably prevented outbreaks in 2023(9,10).

A low incidence of mpox has continued in England during 2023 (137 reported cases), with half being acquired overseas(6). Due to potentially waning immunity and turnover of the GBMSM population, future outbreaks could become more likely if there is no vaccination programme to maintain population immunity levels. We use modelling to determine the most cost-effective vaccination strategy for minimising future mpox outbreaks in England.

## Methods

We adapted our existing deterministic compartmental model of mpox transmission among GBMSM in England(10) to simulate the impact and cost-effectiveness of different vaccination strategies over 20-years from 2024. The model was parameterised and calibrated using surveillance data from the 2022 outbreak and two GBMSM behavioural surveys undertaken in 2021 and 2022(11,12). The cost per quality-adjusted life year (QALY) gained was used to compare various vaccination strategies targeting GBMSM at high-risk of mpox. A health services perspective was assumed for estimating costs.

### Model Structure

We initiated our mpox model(10) in April 2022 with a fully susceptible population. We then assumed a rate of imported mpox cases based on observed data (Poisson process). Susceptible individuals become infected through exposure to mpox cases, following which they become latently infected, but not infectious. They then enter the infectious period, which is stratified into three levels of disease severity (mild, moderate, and severe) based on observed cases in England(13). Following this, individuals either recover and develop immunity or are diagnosed and isolated after a specific time-period, defining the effective infectious period. To project the impact of vaccination, the model was stratified by vaccination status: none, first dose, and second dose. We calibrated the model to the England outbreak up to 12 August 2022 to estimate mpox-related model parameters(10). The model also incorporates recruitment of new GBMSM and waning immunity following vaccination and post-infection.

The model uses the number of men who self-identify as gay and bisexual in England as a proxy for the GBMSM population (769,000(14)). This is divided into four groups at lower and higher risk of mpox transmission and whether they recently attend sexual health services (SHS) or not. The risk strata are included because most mpox cases occurred among high-risk GBMSM in 2022(10) and vaccination is targeted to these individuals. Stratification by SHS status was included because vaccination strategies of GBMSM generally occur through SHS.

The three levels of disease severity capture differences in health care costs and quality-of-life. Cases with severe disease were defined as those having inpatient hospital care, while moderate disease cases required outpatient hospital care and mild disease cases had no hospital care but attended SHS. During the 2022 outbreak, the UK reported no deaths attributable to mpox(4), and so this was not included in the model. A simplified model schematic is in Figure 1, with a full model description in Appendix pp3-13.

**Figure 1.**
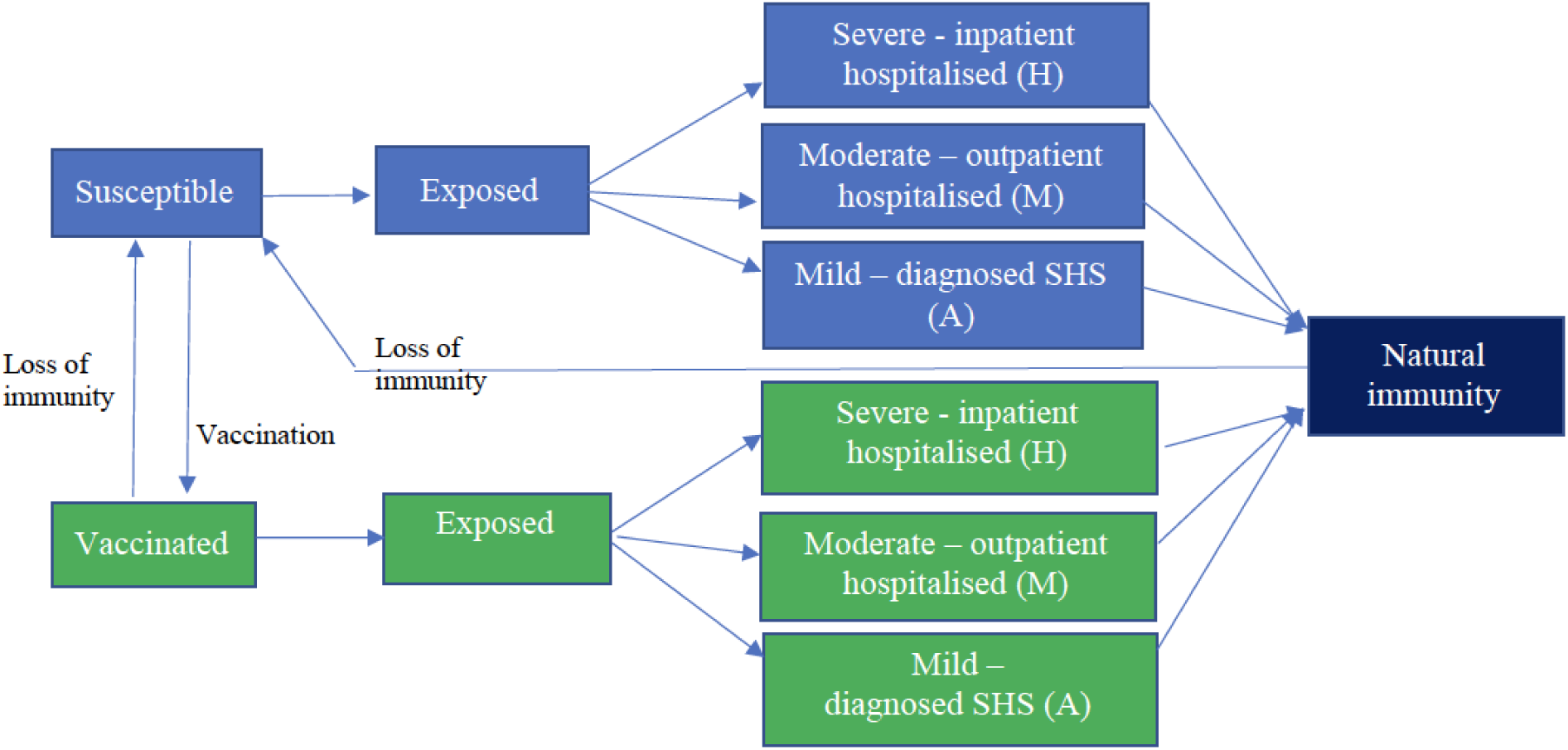
Schematic of the mpox transmission model among GBMSM for assessing the impact and cost-effectiveness of different future vaccination scenarios. Stratifications by low and high risk for mpox infection and attendance at sexual health services are not shown, and vaccination is only shown as one stratification although there are stratifications for 1 or 2 doses of vaccine in the model.

### Model parameterisation and calibration

The model was calibrated to the 2022 outbreak and then run till end of 2023, after which we modelled different vaccination scenarios from 2024. Data used to parameterise and calibrate the model to the 2022 outbreak are described elsewhere(10) and briefly here.

Up to 17 November 2022, data on actual imported cases were used in the model(10). Following this, we assumed an average importation rate (∼6 cases per month) based on the observed number of mpox cases in England acquired outside the UK in 2023 (n=73)(6).

Risk status was defined based on whether GBMSM have <10 or ≥10 anal sex partners in last four months, with the proportion in these categories and attending SHS being estimated from the 2021 Reducing Inequalities In Sexual Health (RiiSH) study, an internet-based survey of 1,039 GBMSM in England (Table 1)(11). Our previous analysis showed that reductions in sexual risk behaviour were important for calibrating this model to the 2022 outbreak in England, and so the same assumptions were made for this model.

**Table 1.** Posterior estimates of transmission parameters for model.

| Model parameter | Median (95%CrI) | Reference |
| --- | --- | --- |
| Transmission coefficient | 0.56 (0.44,0.71) | (10) |
| Timepoint of outbreak (days) after which contact rate and infectious period decrease | 50.56 (40.28,68.51) |  |
| Relative reduction in contact rate after timepoint in outbreak | 0.45 (0.25,0.56) |  |
| Effective infectious period (days) at start of outbreak | 3.02 (2.54,3.93) |  |
| Effective infectious period (days) after timepoint in outbreak | 2.38 (1.98,3.31) |  |
| Parameter defining the rate of change in behaviour and effective infectious period after timepoint in outbreak (Appendix pp7-8) | 0.016 (0.011,0.039) |  |
| Effectiveness of 1<sup>st</sup> dose of vaccine | 0.78 (0.54–0.89) | (5,6) |
| Effectiveness of 2<sup>nd</sup> dose of vaccine | 0.89 (0.78–0.99) | (16) |
| Effectiveness of natural infection for protecting against re-infections | 1.00 | Assumption |
| Duration of protection due to 1<sup>st</sup> dose of vaccine | 5.0 years | Assumption |
| Duration of protection due to 2<sup>nd</sup> dose of vaccine | 10 years | Assumption |
| Duration of protection due to natural infection | 10 years | Assumption |
| Mean rate of new infections entering population per month | 6 | (6) |
| Criteria for start of outbreak response | 120 cases in 3 months | Assumption |
| Criteria for end of outbreak response | 60 cases in 3 months | Assumption |
| Proportion of GBMSM that are high-risk for mpox | 11.9% | (12) |
| Mean contact rate for low and high risk GBMSM | 1.6 (low), 22.3(high) | (12) |

Vaccination for mpox started on 26 June 2022. Until end of 2023, vaccines were assigned in the model among high-risk GBMSM based on weekly vaccination numbers for first and second doses(15). Based on estimates from the 2022 outbreak, we assumed a vaccine effectiveness against mpox infection of 78% (95%CI: 54-89%)(5,6) for one dose and 89% (95%CI: 78-99%)(16) for two doses. However, the duration of protection provided by the vaccine is uncertain. Based on immunological principles and experience with similar vaccines, we expect protection to last over 5 years(17). For our baseline scenario, we assumed a duration of protection of 5 and 10 years for one and two doses, respectively, to prevent the model projecting outbreaks in 2023, which were not observed in England. The duration of immunity induced by natural infection was assumed to be equivalent to receiving two vaccine doses, with 100% effectiveness against infection. This allows for small levels of reinfection from 2023 onwards(18) because of waning natural protection.

Bayesian Markov chain Monte Carlo sampling was used to calibrate the model to case data among male individuals in England from 17 April to 12 August 2022. The model fits were validated against case data from 13 August to 16 November 2022. The model fits were used to project the impact of different vaccination strategies from 2024, with the range in estimates across the model fits being used to estimate the mean and 95% credibility interval (95%CrI) around all projections. More details on the model parameterisation and calibration are in Appendix pp7-10.

### Health Utilities

We estimated baseline QALY weights for GBMSM without mpox using health-related quality of life (HRQoL) data collected through EQ-5D-3L questionnaires in the RiiSH-MPOX survey undertaken in December 2022(12). As no QALY estimates were available for mpox, we assumed disutilities for mild, moderate, and severe mpox cases based on disability weights for mild (0.006 (0.002–0.012)), moderate (0.051 (0.032–0.074)), and severe (0.133 (0.088–0.190)) acute infectious disease from the Global Burden Diseases 2021 study(19). Disability weights were assumed to be the same as disutilities for calculating QALYs and subtracted from the baseline HRQoL data to estimate the impact over a year of infection with mpox (Table 2). Additionally, HRQoL utility weights from a UK study of herpes zoster (presumed similar pain levels to mpox)(20) were used to expand the uncertainty bounds around our estimates, assuming infection lasted 21 days. Further details in Appendix pp20.

**Table 2:** Cost and utility parameters. All costs are presented in 2022 British Pounds (£)

| Parameters | Base case value (lower–upper bound)* | Source/Comments |
| --- | --- | --- |
| **Unit Costs\*\*** |  |  |
| **Clinical case management costs** |  |  |
| Mild cases | £378.63 (£303.22 - £454.04) | (13,22-24) and Clinical experts. Patients with mild symptoms were assumed to be diagnosed and treated at sexual health services (SHS) and isolated at home. Mild symptoms typically presented with few lesions. All cases were confirmed with PCR by Rare and Imported Pathogens Laboratory (RIPL). |
| Moderate cases | £730.67 (£577.00 - £730.67) | (22,25,26). Patients with moderate level of symptoms were defined as those who attended hospital through the emergency department (ED) and were not admitted. An estimated 14.4% of these patients had attended SHS before visiting ED. |
| Severe cases | £3,991.22 (£3,776.00 – £3,991.22) | (21,22,25,26). Patients with severe mpox symptoms were defined as those who received inpatient hospital care. Based on analysis of Secondary Uses Service (SUS) and the emergency care data set (ECDS), 63.4% of patients admitted to hospital for mpox first attended ED prior to admission. We assumed the remainder (36.6%) attended SHS prior to hospital admission and ~4% of hospitalized patients required intensive care |
| **Vaccination administration costs** |  |  |
| First dose administration | £36.85 (fixed) | (22). Costs include staff time costs for registration, consultation, vaccine administration, and health promotion, as well as consumables such as gloves, plasters and cotton |
| Second dose administration | £25.78 (fixed) | (22). Costs assume a reduced consultation time compared to initial visit |
| Vaccine cost | £160.00 (fixed) | (27). Assumed cost of Shingrix vaccine |
| **Public health response costs** |  |  |
| Weekly outbreak costs | £81,290 (£73,714 - £88,866) | Public health response costs (pay and non-pay costs) were gathered from: i) the UKHSA regional teams where most Mpox cases occurred in 2022, ii) the central UKHSA National Response Centre, and iii) cross-UKHSA Enhanced incident costs from the UKHSA Directorate of Emergency Preparedness, Resilience and Response. This data was used to estimate the cost of a national response (for England) including both regional and central response activity. |
| Per case costs | £416 (£184 - £779) | Per case cost covers the cost associated with staffing mpox cell in London and across England. |
| **Utilities** |  |  |
| Baseline utility | 0.839 (fixed) | (12) |
| Mild Mpox | 0.833 (0.827 – 0.837) | (19,20) |
| Moderate Mpox | 0.788 (0.765 – 0.829) | (19,20) |
| Severe Mpox | 0.706 (0.649 – 0.821) | (19,20) |

\*All uncertainty ranges were sampled using triangle distribution.

### Costs

A health services perspective was used for estimating costs (2022 British pounds £) in the baseline analysis. Health-related costs for clinical management, public health responses during an outbreak and vaccination were included. Data on these costs were gathered from various sources. Uncertainty was associated with all cost estimates in our cost-effectiveness projections. Brief details of the different costs are given below and Table 2, with more details in Appendix pp10-13 and 14-19.

#### Clinical management costs

The proportion of mpox cases with each level of severity, and care accessed by these mpox patients was calculated from Secondary Uses Service (SUS), the emergency care data set (ECDS), clinical evidence(13,21–26), and checked with clinical experts. The estimated cost per case was £379 (£303-454), £731 (£577–731), and £3,991 (£3,776-3,991) for mild, moderate and severe mpox, respectively.

#### Vaccination costs

The cost of administering the mpox vaccine was obtained from the SHS tariff for mpox(22), with the estimated costs being £36.85 for the first dose and £25.78 for the second dose. The cost of the mpox vaccine is confidential, so we used the cost for the Shingrix vaccine (£160 per dose(27); thought to have a comparable cost) in the baseline analysis, and also estimated the maximum vaccine cost for each intervention to be cost-effective.

#### Public health response (PHR) costs

Costs for PHR measures taken by UKHSA during the outbreak include costs related to each case (e.g., contact tracing) and outbreak-related overhead costs. Data was obtained from three England regions and central UKHSA. The estimated PHR cost per case was £416 (£184-779) and the outbreak-related overhead cost was £81,290/week (£73,714-88,886).

### Future vaccination scenarios

From January 2024, we modelled the costs and impact of the following vaccination scenarios (Appendix pp8-10):

- **Counterfactual scenario:** No vaccination from 2024 onwards, although we assume vaccination in 2022-2023.
- **Pre-emptive vaccination irrespective of outbreak:** Vaccines are given continuously to high-risk GBMSM attending SHS at constant rates of 13, 27, 41, 54, 81, and 135 per day. These rates were chosen because they achieve a vaccination coverage of 5, 10, 15, 20, 30, and 50% in a year, respectively, if initiated from scratch.
- **Reactive vaccination when outbreak triggered:** Vaccines are given to high-risk GBMSM attending SHS at constant rate once an outbreak response is triggered and stopped when outbreak ends. We consider the rate achieved in the 2022 outbreak, 465 per day, and lower rates as for pre-emptive vaccination.

For reactive vaccination, our baseline analysis assumes that an outbreak response is triggered if >120 cases in 3 months and ends when <60 cases in 3 months. For all scenarios, we assume that a PHR occurs in the event of an outbreak, and that the same reduction in sexual risk behaviour occurs as in the 2022 outbreak(10). In all vaccination scenarios, we assume first doses are distributed among susceptible individuals and second doses are administered to those who have not yet received them, with 56% of vaccine doses being given as second doses (based on vaccine data(15)). However, if all high-risk GBMSM attending SHS become protected (vaccinated with first dose or have natural immunity), then remaining vaccinations are provided as second doses. The Appendix (pp9) give details of when vaccination levels saturate among high-risk GBMSM, which depends on the duration of protection. Fractional doses are considered as a sensitivity analysis.

### Cost-effectiveness analysis

We evaluated the incremental cost-effectiveness ratio (ICER) in terms of the incremental cost per QALY gained for each vaccination scenario, and net monetary benefit (NMB), over a 20-year period from January 2024. This involved comparing the estimated costs and QALYs for each vaccination scenario to the counterfactual scenario, as well as comparing different vaccination scenarios against each other to undertake a full incremental cost-effectiveness analysis. The latter analysis involves ordering the vaccination scenarios in terms of increasing cost, and calculating incremental costs and QALY gains, eliminating any scenario which are dominated (higher cost and fewer benefits) or extendedly dominated (higher ICER and fewer benefits). These comparisons were done to determine the most cost-effective vaccination scenario. In addition, the NMB was calculated for each scenario, calculated as QALYs multiplied by the willingness-to-pay (WTP) threshold minus the costs for that intervention. The highest NMB or incremental net benefit (INB, difference between the intervention NMB and counterfactual NMB) represents the most cost-effective scenario at a particular WTP threshold, with any NMB>0 being cost-effective at that threshold.

For all comparisons, probabilistic sensitivity analyses were undertaken that sampled model, utility and cost parameters 500 times to produce uncertainty bounds around the incremental costs and QALYs. Costs and QALYs were discounted at 3.5% per year and WTP thresholds of £20,000 or £30,000 per QALY were used, as recommended by the Joint Committee on Vaccination and Immunisation (JCVI)(28). We also estimated the maximum threshold vaccine price for reactive and pre-emptive vaccination to be cost-effective defined as 50% of probabilistic ICER estimates below the £20,000/QALY threshold or 90% below £30,000/QALY(28).

### Sensitivity analyses

Because of numerous parameter uncertainties and assumptions, we undertook sensitivity analyses to evaluate whether they changed which vaccination strategy was most cost-effective. Sensitivity analyses include the following changes, with all other parameters and assumptions remaining the same as in the baseline analysis:

1. Annual discount rate of 1.5% instead of 3.5%;
2. Only include direct health care costs so remove PHR costs from baseline;
3. Societal perspective - Include productivity losses due to mpox disease (absenteeism and presenteeism), estimated using data from the RiiSH-MPOX survey(12) and ONS data on employment and average salaries (Appendix pp19-20);
4. Long duration of vaccine protection of 10 and 20 years for one and two doses, respectively, instead of 5 and 10 years;
5. Short duration of vaccine protection of 2.5 and 5.0 years for one and two doses, respectively;
6. Low rate of 1 imported case per month instead of 6;
7. High rate of 10 imported cases per month;
8. Low outbreak response criteria of 96 cases in 3 months, which ends when <48 cases in 3 months (baseline outbreak criteria was 120 cases in 3 months, which ends when <60 cases in 3 months);
9. High outbreak response criteria of 144 cases in 3 months, which ends when <72 cases in 3 months;
10. No reductions in risk behaviour during outbreaks; baseline assumes decrease occurs during each outbreak;
11. Alternative definition of low- and high-risk GBMSM using data on rates of physical contact from the RiiSH-MPOX survey(12);
12. Assume breakthrough infections have half the chance of experiencing moderate and severe mpox disease than in baseline scenario;
13. Half of vaccinations are delivered as fractional (1/4) doses with assumed 25% lower effectiveness following 1 and 2 doses.

## Results

### Baseline impact projections

With no vaccination from 2024 (counterfactual scenario), the baseline model projects there will be outbreaks nearly every year (Figure S3), with the yearly number of cases increasing from 306 (95%CrI 257-485) in 2024 to 3,332 (95%CrI 2474-5998) in 2033 (Figure 2). Over 2024-2043, there would be an estimated 5,472 (95%CrI: 5,141–5676) outbreak days (when outbreak is triggered) and 54,891 (95%CrI: 26,246– 67,256) infections (Table 3). With vaccination, outbreaks are reduced. For instance, with pre-emptive vaccination strategies that deliver ≥41 vaccine doses per day, the yearly median number of cases remains below 300 over 2024-2043 (including incoming infections; Figure 2). This same level of new cases is also achieved with reactive vaccination strategies that deliver ≥81 vaccine doses per day when an outbreak is triggered.

**Figure 2.**
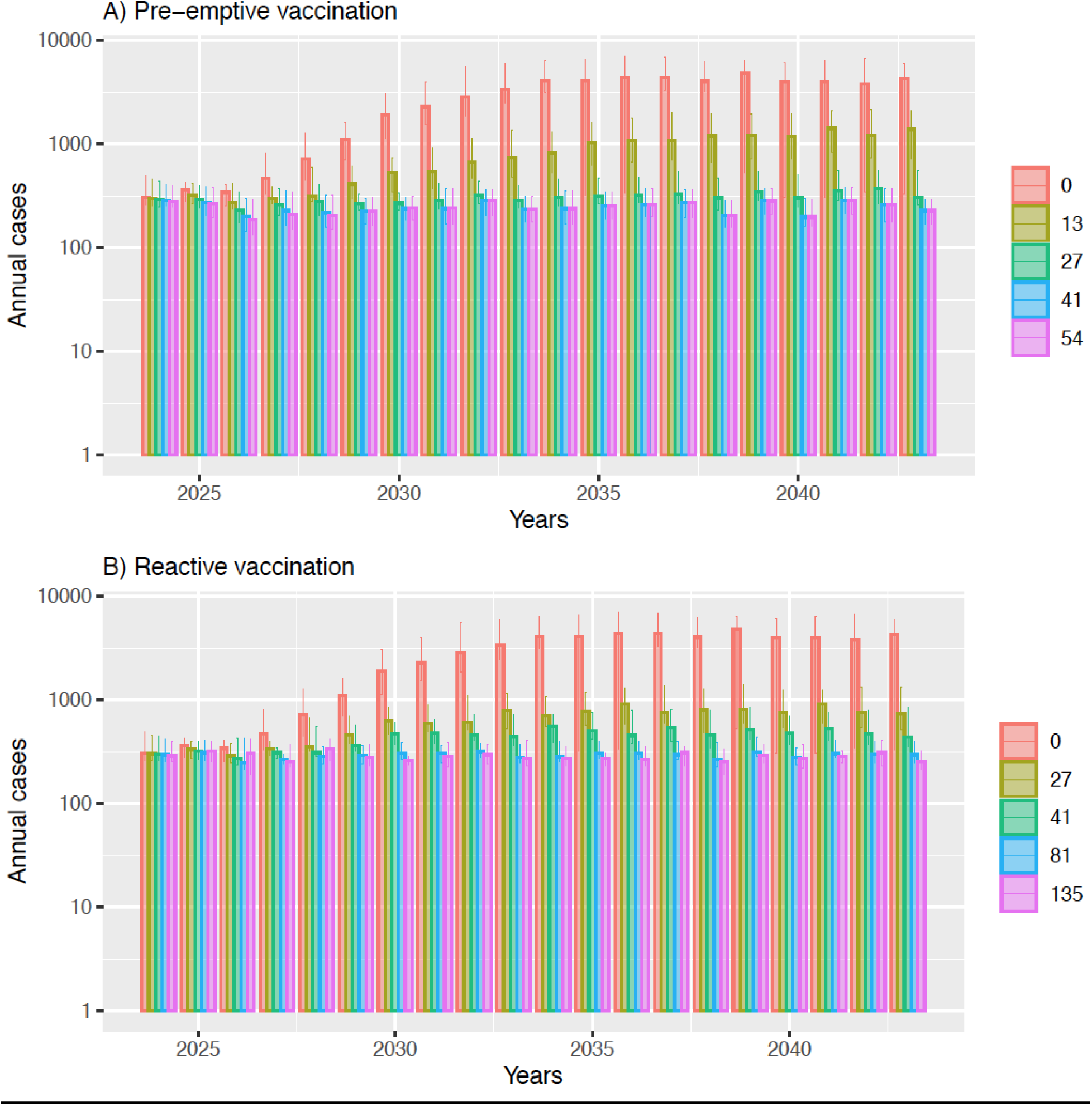
Annual cases over next 20 years (log scale) under different vaccination scenarios. Modelled vaccination rates are 13,27, 41 and 54 doses per day for pre-emptive vaccination and 27, 41, 81 and 135 doses per day for reactive vaccination. The error bars represent 95% CrI.

**Table 3.** Epidemic characteristics, costs and cost-effectiveness projections for different vaccination programmes over 2024-2043. Point values are means and ranges are 95% credibility intervals.

| Vaccination scenario | Total number of infections | Outbreak duration (days) | Total Cost (1000s £) | Total QALYs | Incremental costs compared to no vaccination (1000s £) | Incremental QALY compared to no vaccination | ICER compared to no vaccination | Fully incremental analysis | Incremental Net Benefit compared to no vaccination (£20,000/QALY) | Incremental Net Benefit compared to no vaccination (£30,000/QALY) |
| --- | --- | --- | --- | --- | --- | --- | --- | --- | --- | --- |
| No Vaccination counterfactual | 54891<br>[26246,67256] | 5472<br>[5141,5676] | 86127<br>(67671, 102039) | 9493798<br>(9493540, 9494085) | — | — | — | Dominated | — | — |
| Pre-emptive vaccination (PV) of GBMSM at high risk for mpox who attend SHS |  |  |  |  |  |  |  |  |  |  |
| PV 13 vaccines per day | 13099<br>[8224,18452] | 4219<br>[3494,4673] | 59184<br>(48271, 68408) | 9494228<br>(9494138, 9494307) | -26944<br>(-37747,-15225) | 429.59<br>(182.58,614.47) | Cost-saving | Dominated | 35535635<br>(17860583, 48118143) | 39831564<br>(19393579, 53736914) |
| PV 27 vaccines per day | 2143<br>[637,3860] | 2210<br>[802,3198] | 49320<br>(36731, 59342) | 9494331<br>(9494299, 9494358) | -36807<br>(-49976, -26202) | 532.62<br>(259.29,767.60) | Cost-saving | Dominated | 47459759<br>(32700262, 62948345) | 52785985<br>(35054011, 69276410) |
| **PV 41 vaccines per day** | **254**<br>**[0,1143]** | **369**<br>**[0,1719]** | 46527<br>(42497, 58089) | **9494345**<br>**(9494318, 9494369)** | **-39600**<br>**(-54355, -24480)** | **546.26**<br>**(278.56,780.80)** | **Cost-saving** | **Lowest cost scenario** | **50525563**<br>**(30140660, 67252007)** | **55988154**<br>**(33204347, 74114515)** |
| PV 54 vaccines per day | 234<br>[0,1101] | 340<br>[0,1641] | 47052<br>(43363, 58261) | 9494346<br>(9494319, 9494369) | -39075<br>(-53551,-24088) | 547.23<br>(279.58,780.85) | Cost-saving | Extendedly dominated | 50020084<br>(29279217, 66520901) | 55492428<br>(32351740, 73334039) |
| PV 81 vaccines per day | 221<br>[0,1064] | 325<br>[0,1594] | 47345<br>(43851, 58526) | 9494346<br>(9494319, 9494370) | -38782<br>(-53152,-23638) | 547.84<br>(280.23,782.34) | Cost-saving | 517,722/QALY compared to PV 41/day | 49739170<br>(29187177, 66097299) | 55217523<br>(31873113, 73308040) |
| PV 135 vaccines per day | 203<br>[0,1022] | 305<br>[0,1523] | 47487<br>(44244, 58324) | 9494346<br>(9494320, 9494370) | -38640<br>(-53376,-23279) | 548.10<br>(280.66,783.00) | Cost-saving | 546154/QALY compared to PV 81/day | 49602554<br>(28809626, 66134550) | 55083603<br>(31489695, 73493452) |
| Reactive vaccination (RV) of GBMSM at high risk for mpox who attend SHS |  |  |  |  |  |  |  |  |  |  |
| RV 27 vaccines per day | 9841<br>[7021,13064] | 4164<br>[3745,4450] | 58714<br>(50229, 65689) | 9494257<br>(9494192, 9494315) | -27413<br>(-38688,-13744) | 458.51<br>(202.78,658.39) | Cost-saving | Dominated | 36583355<br>(17518053, 50913077) | 41168421<br>(19561628, 56329832) |
| RV 41 vaccines per day | 5597<br>[4074,7416] | 3596<br>[3171,3909] | 55820<br>(47818, 62590) | 9494299<br>(9494257, 9494339) | -30308<br>(-42224,-16530) | 501.02<br>(231.93,722.24) | Cost-saving | Dominated | 40328247<br>(20703857, 55334814) | 45338482<br>(23216678, 61957122) |
| RV 81 vaccines per day | 2157<br>[1747,2723] | 2487<br>[2060,2842] | 52487<br>(43539, 60384) | 9494333<br>(9494306, 9494360) | -33640<br>(-45930,-21852) | 534.38<br>(253.95,772.71) | Cost-saving | Dominated | 44327652<br>(26708415, 59583806) | 49671409<br>(29281299, 66767847) |
| RV 135 vaccines per day | 1284<br>[1071,1605] | 1699<br>[1370,2233] | 49561<br>(40180, 60425) | 9494336<br>(9494311, 9494362) | -36566<br>(-49521,-24543) | 537.53<br>(256.33,780.15) | Cost-saving | Dominated | 47317034<br>(30162311, 62626566) | 52692312<br>(32879876, 69889759) |
| RV 204 vaccines per day | 960<br>[729,1565] | 1328<br>[966,2148] | 48913<br>(38452, 61210) | 9494338<br>(9494315, 9494362) | -37215<br>(-50720,-25782) | 539.28<br>(259.57,777.96) | Cost-saving | Dominated | 48000194<br>(32646847, 62458815) | 53392963<br>(35283426, 70202720) |
| RV 465 vaccines per day | 811<br>[462,1530] | 1130<br>[630,2111] | 49996<br>(41530, 61447) | 9494339<br>(9494315, 9494363) | -36132<br>(-49989,-23834) | 540.67<br>(262.46,778.53) | Cost-saving | Dominated | 46944924<br>(30143265, 62114353) | 52351584<br>(33129719, 68782845) |

Cost-saving scenarios save money and gain QALYs compared to the no vaccination counterfactual scenario. Dominated scenarios have higher mean costs and save fewer QALYs (just comparison of mean) than another scenario. Extended dominated scenarios have a higher mean ICER and save fewer QALYs (just comparison of mean) than another scenario. The scenario in bold is the most cost-effective vaccination scenario – it saves most money and no other scenario that saves more QALYs is cost-effective compared to it.
Abbreviations: QALY, Quality Adjusted Life Years; ICER, incremental cost-effectiveness ratio; PV, pre-emptive vaccination; RV, reactive vaccination.

### Baseline cost-effectiveness projections

Over the 20-year period, the counterfactual scenario projects 9,493,798 (95%CrI 9,493,540–9,494,085) QALYs (discounted 3.5% per year). All vaccination scenarios produce greater QALYs. For example, pre-emptive vaccination at 41 vaccine doses per day gains 546 (95%CrI 279–781) QALYs compared to the counterfactual scenario, while reactive vaccination at 81 vaccine doses per day gains 534 (95%CrI 254– 773) QALYs (Table 3).

The total discounted costs of the counterfactual no vaccination scenario is £86,127,476 (95%CrI £67,671,131–102,039,870; Table 3) over 20 years, with clinical case management and PHR costing £25,042,869 (29.5%) and £61,084,608 (70.5%), respectively (Table S13). All the vaccination scenarios decrease the total costs compared to the counterfactual as savings in clinical care management and PHR offset the vaccination cost. Because all vaccination scenarios also gain QALYs, they are cost-saving compared to the counterfactual. The most cost-effective vaccination scenario (highest NMB; Table 3) is pre-emptive vaccination at 41 vaccine doses per day, which saves £39,600,381 (95%CrI 24,480,101-54,355,415) and gains 546 (95%CrI 279-781) QALYs compared to the counterfactual. All other vaccination scenarios have an ICER above £30,000/QALY gained compared to this scenario (Table 3), including all reactive vaccination strategies.

### Threshold vaccine price

For pre-emptive vaccination at 41 vaccine doses per day, the threshold vaccine price for this strategy to be cost-effective is £400 or £368 per dose for the £20,000/QALY and £30,000/QALY WTP criteria, respectively (Table 4). Similarly, for reactive vaccination at 204 vaccine doses per day, the threshold vaccine prices are £426 and £394 per dose for the £20,000/QALY and £30,000/QALY criteria. Threshold vaccine prices are lower at higher vaccination rates and for pre-emptive vaccination (versus reactive vaccination at same vaccination rate; Figure S8).

**Table 4.** Maximum threshold vaccine price (British pounds) for meeting the JCVI willingness-to-pay (WTP) criteria for being cost-effective. Two JCVI WTP criteria are used: V50 represents the vaccine price at which the median ICER projection is below £20,000 per QALY saved, and V90 is the vaccine price at which 90% of ICER projections are below £30,000 per QALY saved. The vaccination scenario is cost-effective below these threshold prices.

| Pre-emptive vaccination of GBMSM attending SHS that are high risk for mpox |  |  | Reactive vaccination of GBMSM attending SHS that are high risk for mpox |  |  |
| --- | --- | --- | --- | --- | --- |
| Vaccination rate | V50 | V90 | Vaccination rate | V50 | V90 |
| 13 doses per day | £701 | £573 | 27 doses per day | £616 | £567 |
| 27 doses per day | £485 | £455 | 41 doses per day | £546 | £509 |
| 41 doses per day | £400 | £368 | 81 doses per day | £464 | £437 |
| 54 doses per day | £392 | £362 | 136 doses per day | £445 | £410 |
| 81 doses per day | £388 | £358 | 204 doses per day | £426 | £394 |
| 136 doses per day | £385 | £354 | 465 doses per day | £396 | £371 |

### Sensitivity analyses

The effect of the sensitivity analyses on the incremental costs and QALYs for each vaccination scenario are shown in Appendix Tables S14-S28 and described in Appendix pp21-25. These sensitivity analyses change the incremental costs and QALYs saved, but most do not change which vaccination scenario is most cost-effective. Pre-emptive vaccination remains the most cost-effective scenario when we assumed a longer duration of vaccine protection or lower rate of imported infections, although a lower vaccination rate is optimal (27 vaccine doses per day instead of 41). Similarly, pre-emptive vaccination remains the most cost-effective scenario, but at a higher vaccination rate (54, 135 or 271 vaccine doses per day), when we assume a shorter duration of vaccine protection, higher rate of imported infections or no reduction in risk behaviour during future outbreaks. Assuming different outbreak response criteria has no effect on the preferred vaccination scenario, nor does changes in the discount rate, how we define high-risk GBMSM, assuming breakthrough infections have less severe symptoms, or including productivity losses from mpox. The only sensitivity analysis that changes the preferred vaccination strategy is when we don’t include PHR costs, where the most cost-effective approach becomes reactive vaccination at 27 vaccine doses per day.

## Discussion

Utilising an mpox transmission model, we evaluated the cost-effectiveness of different vaccination strategies for controlling future mpox outbreaks among GBMSM in England. If the vaccine has moderate cost (£160 per dose), our analyses project that vaccinating for mpox is always better than not vaccinating; it reduces the number and size of outbreaks, saves money considerably and gains QALYs. Baseline projections suggest the preferred strategy is routine pre-emptive vaccination of high-risk GBMSM at a low rate (41 doses per day), with pre-emptive vaccination remaining the preferred strategy over most sensitivity analyses, although the optimal vaccination rate can change. Vaccination only becomes not cost-effective if the vaccine cost is high (>£701 per dose).

### Strengths and Limitations

To our knowledge, this is the first cost-effectiveness analysis of undertaking vaccination for mpox. Strengths of the analysis include its use of detailed cost data from the 2022 outbreak in England and the use of a published and validated model calibrated to the England outbreak(10), both of which increases the validity of our projections.

However, there were uncertainties associated with our analysis. Extensive sensitivity analyses and probabilistic uncertainty analyses were undertaken to assess the robustness of our findings (described below), with these analyses showing that pre-emptive vaccination is the preferred strategy unless public health response costs are not included.

There is uncertainty around the duration of protection provided by the vaccine, which should be assessed through longer-term follow-up of vaccinated individuals. Our sensitivity analyses show that assuming shorter or longer duration of protection affects the impact achieved by vaccination, but the most cost-effective strategy remains pre-emptive vaccination, although the optimal vaccination rate decreases if the duration of protection is longer than we assumed or increases if duration of protection is shorter. Uncertainty in the future importation rate of new mpox cases has a similar effect. Pre-emptive vaccination remains the optimal strategy, but the optimal vaccination rate decreases for lower importation rates and increases for higher importation rates.

Uncertainty exists in the sexual risk behaviour of high-risk GBMSM and whether risk behaviour will reduce during future outbreaks, as it did in 2022(12). Interestingly, there was no change to our optimal vaccination scenario when we used different data to define high-risk GBMSM. Conversely, when we assumed no reduction in risk behaviour during future outbreaks, then the size of the outbreaks increased dramatically without vaccination. This results in higher vaccination rates being needed to control these outbreaks, and more costs being saved from doing so. Despite these changes, pre-emptive vaccination remains the optimal strategy but at a higher vaccination rate.

Other uncertainties related to whether specific costs should be incorporated. For instance, if we do not include public health response costs during each outbreak, as done in economic evaluations of vaccine initiatives for endemic diseases, then less costs are saved from vaccination and only low vaccination rates are cost-effective with reactive vaccination being the preferred strategy. Conversely, if we include societal costs such as productivity losses related to mpox, then the preferred vaccination scenario does not change but more costs are saved from vaccination. It is also uncertain which criteria in terms of new cases will be used to trigger an mpox outbreak response, however, we found this did not affect the preferred vaccination strategy.

Evidence suggests that the clinical features of re-infections or breakthrough infections after vaccination are less pronounced than was found in the 2022 outbreak(29), and so their management costs could be less than we estimated. However, when we assumed breakthrough infections had half the chance of experiencing moderate and severe mpox disease, we found that the optimal vaccination scenario remained unchanged.

There were no mpox-specific HRQoL utility weights and so we used general utility weights for infectious diseases from the Global Burden of Disease study(19) which may not relate well to mpox disease. To counter this issue, we included large uncertainty ranges around our utility estimates, also using utility weights for Herpes Zoster(20), and our model projections were robust despite this.

Lastly, we did not include worse outcomes among people living with HIV(30) because most GBMSM living with HIV in England are virally suppressed(31). In other settings, vaccination for mpox may be more cost-effective because it also reduces the severity of infection among people with HIV(32).

### Comparison with literature

Numerous model analyses of the mpox outbreak in 2022(9,10,33) have suggested that the existing roll-out of vaccination has been important for preventing future resurgences. Other model analyses have suggested that future outbreaks could be controlled through vaccinating close contacts of cases(34) or individuals with many sexual contacts(35), especially if done pre-emptively(36). Our current study builds on these analyses by evaluating the cost-effectiveness of future vaccination strategies. This is crucial new evidence for determining the optimal vaccination strategy going forward.

### Implications

Our analysis shows that pre-emptive routine vaccination of high-risk GBMSM attending sexual health services is likely to be a cost-saving strategy for preventing future mpox outbreaks in England. These findings underpinned the evidence considered by the UK JCVI for recommending this vaccination strategy in England(37). The robustness of our findings suggest similar vaccination strategies should be considered by other high-income countries for minimising mpox outbreaks going forward. In more resource limited countries with ongoing mpox transmission, the cost of the vaccine needs to be minimised to enable wide-spread vaccination campaigns. Future studies need to evaluate the optimal vaccination strategies in such countries.

## Data Availability

This analysis and modelling were undertaken for health protection purposes under permissions granted to UKHSA to collect and process confidential patient data under Regulation 3 of The Health Service (Control of Patient Information) Regulations 2020 and Section 251 of the National Health Service Act 2006. All data were pseudonymised during analysis, and records were stored securely. As such, authors cannot make the underlying datasets publicly available for ethical and legal reasons, particularly due to the sensitive information included. Applications for relevant anonymised data should be submitted to the UKHSA Office for Data Release at https://www.gov.uk/government/publications/accessing-ukhsa-protected-data. The model code and projections for this paper will be shared with interested parties upon reasonable request, which will be decided by Peter Vickerman, Josephine Walker and Xu-Sheng Zhang.

## Contributors

PV, JW, XSZ, SM and HM conceived and designed the study. XSZ developed the model and performed all model analyses. JGW and SN led and undertook the costing analysis and oversaw the cost-effectiveness analysis. Numerous authors provided data and/or undertook additional data analyses for costing the interventions and calibrating the model. PV oversaw the overall analysis. XSZ wrote the initial draft of the manuscript with PV. All authors contributed to guiding the overall analysis plan, interpreting interim and final results, and critically reviewing the final version of the manuscript. XSZ, JGW and PV have directly accessed and verified the underlying data of this research article, and PV made the final decision to submit the manuscript.

## Acknowledgements

PV received funding for this analysis from the Health Protection Research Unit for Evaluation of Interventions and Behavioural Science [NIHR200877] funded by the UK National Institute for Health Research. PV is also funded by the Wellcome Trust (WT 220866/Z/20/Z). The study sponsors had no involvement in the study design; in the collection, analysis, and interpretation of data; in the writing of the report; and in the decision to submit the paper for publication.

## References

1. Thornhill JP, Barkati S, Walmsley S, et al. Monkeypox Virus Infection in Humans across 16 Countries - April-June 2022. N Engl J Med. 2022;387(8):679–91.

2. World Health Organisation. Responding to the global mpox outbreak - Ethics issues and considerations: A policy brief (https://iris.who.int/bitstream/handle/10665/371405/WHO-Mpox-Outbreak-response-Ethics-2023.1-eng.pdf?sequence=1.). 2023.

3. World Health Organisation. 2022-23 Mpox (Monkeypox) Outbreak: Global Trends 2024 (https://worldhealthorg.shinyapps.io/mpx_global/). 2024.

4. UK Health Security Agency. Investigation into monkeypox outbreak in England: technical briefing 8 (https://www.gov.uk/government/publications/monkeypox-outbreak-technical-briefings) https://www.gov.uk/government/publications/monkeypox-outbreak-technical-briefings 2022 [

5. Bertran M, Andrews N, Davison C, et al. Effectiveness of one dose of MVA-BN smallpox vaccine against mpox in England using the case-coverage method: an observational study. Lancet Infect Dis. 2023.

6. Charles H, Thorley K, Turner C, et al. Post-peak mpox in England: epidemiology, reinfection, and vaccine effectiveness – data from 2023 (). medRxiv (https://doiorg/101101/2024022624303362). 2024.

7. Dalton AF, Diallo AO, Chard AN, et al. Estimated Effectiveness of JYNNEOS Vaccine in Preventing Mpox: A Multijurisdictional Case-Control Study - United States, August 19, 2022-March 31, 2023. MMWR Morb Mortal Wkly Rep. 2023;72(20):553–8.

8. Kennedy RB, Ovsyannikova IG, Jacobson RM, Poland GA. The immunology of smallpox vaccines. Curr Opin Immunol. 2009;21(3):314–20.

9. Brand SPC, Cavallaro M, Cumming F, et al. The role of vaccination and public awareness in forecasts of Mpox incidence in the United Kingdom. Nat Commun. 2023;14(1):4100.

10. Zhang XS, Mandal S, Mohammed H, et al. Transmission dynamics and effect of control measures on the 2022 outbreak of mpox among gay, bisexual, and other men who have sex with men in England: a mathematical modelling study. Lancet Infect Dis. 2024;24(1):65–74.

11. Brown JR, Reid D, Howarth AR, et al. Sexual behaviour, STI and HIV testing and testing need among gay, bisexual and other men who have sex with men recruited for online surveys pre/post-COVID-19 restrictions in the UK. Sex Transm Infect. 2023.

12. Ogaz D, Enayat Q, Brown JRG, et al. Mpox Diagnosis, Behavioral Risk Modification, and Vaccination Uptake among Gay, Bisexual, and Other Men Who Have Sex with Men, United Kingdom, 2022. Emerg Infect Dis. 2024;30(5):916–25.

13. Fink DL, Callaby H, Luintel A, et al. Clinical features and management of individuals admitted to hospital with monkeypox and associated complications across the UK: a retrospective cohort study. Lancet Infect Dis. 2022.

14. Office of National Statistics. Sexual orientation in the UK from 2012 to 2020 by region. (https://www.ons.gov.uk/datasets/sexual-orientation-by-region/editions/time-series/versions/2) 2022 [

15. National Health Service England (NHS-England). Vaccinations: Mpox (https://www.england.nhs.uk/statistics/statistical-work-areas/vaccinations-for-mpox/) 2024 [

16. Xu M, Liu C, Du Z, Bai Y, Wang Z, Gao C. Real-world effectiveness of mpox (monkeypox) vaccines: a systematic review. J Travel Med. 2023.

17. Berry MT, Khan SR, Schlub TE, et al. Predicting vaccine effectiveness for mpox. Nat Commun. 2024;15(1):3856.

18. Li T, Li Z, Xia Y, Long J, Qi L. Mpox reinfection: A rapid systematic review of case reports. Infect Med (Beijing). 2024;3(1):100096.

19. Global Burden of Disease Forecasting Collaborators. Burden of disease scenarios for 204 countries and territories, 2022-2050: a forecasting analysis for the Global Burden of Disease Study 2021. Lancet. 2024;403(10440):2204–56.

20. Gater A, Abetz-Webb L, Carroll S, Mannan A, Serpell M, Johnson R. Burden of herpes zoster in the UK: findings from the zoster quality of life (ZQOL) study. BMC Infect Dis. 2014;14:402.

21. Dugbazah B, Mindlin M. Monkeypox hospitalisation interim report. 2023.

22. Phillips D. Tariff Calculator for Monkeypox activities in Sexual Health Services using the Integrated Sexual Health Tariff (ISHT) Methodology. 2022.

23. National Health Service (NHS). Chief Medical Officer Alert: Tecovirimat as a Treatment for Patients Hospitalised due to Monkeypox Viral Infection (https://www.google.com/url?sa=t&source=web&rct=j&opi=89978449&url=https://www.cas.mhra.gov.uk/ViewandAcknowledgment/ViewAttachment.aspx%3FAttachment_id%3D104081&ved=2ahUKEwiDtKWF1OWGAxVoQUEAHcs3DaQQFnoECA8QAQ&usg=AOvVaw1BozWk0DGCQbiRkvEHHFi4 2022 [

24. National Health Service England (NHS-England). NHS Drug Tariff (https://www.drugtariff.nhsbsa.nhs.uk/#/00837338-DC/DD00837329/Home) 2023 [

25. National Health Service England (NHS-England). National Cost Collection: National Schedule of NHS Costs - Year 2021/22 - NHS trusts and NHS foundation trusts 2023 [

26. Jones K, Weatherly H, Birch S, Castelli A, Chalkley M, Dargan A. Unit Costs of Health and Social Care 2022 Manual. Technical report. (doi:10.22024/UniKent/01.02.100519). Personal Social Services Research Unit (University of Kent); Centre for Health Economics (University of York) K, UK., editor2022.

27. National Institute for Health and Care Excellence (NICE). British National Formulary (BNF; https://bnf.nice.org.uk/drugs/) 2023 [

28. Department of Health and Social Care. Cost-effectiveness methodology for vaccination programmes (https://assets.publishing.service.gov.uk/media/5afd3c17ed915d0de80ffd6c/cemipp-consultation-document.pdf). 2018.

29. Hazra A, Zucker J, Bell E, et al. Mpox in people with past infection or a complete vaccination course: a global case series. Lancet Infect Dis. 2024;24(1):57–64.

30. Mitja O, Alemany A, Marks M, et al. Mpox in people with advanced HIV infection: a global case series. Lancet. 2023;401(10380):939–49.

31. UK Health Security Agency. HIV Action Plan monitoring and evaluation framework (https://www.gov.uk/government/publications/hiv-monitoring-and-evaluation-framework/hiv-action-plan-monitoring-and-evaluation-framework) 2022 [

32. Schildhauer S, Saadeh K, Vance J, et al. Reduced Odds of Mpox-Associated Hospitalization Among Persons Who Received JYNNEOS Vaccine - California, May 2022-May 2023. MMWR Morb Mortal Wkly Rep. 2023;72(36):992–6.

33. Guzzetta G, Marziano V, Mammone A, et al. The decline of the 2022 Italian mpox epidemic: Role of behavior changes and control strategies. Nat Commun. 2024;15(1):2283.

34. Yuan P, Tan Y, Yang L, et al. Modeling vaccination and control strategies for outbreaks of monkeypox at gatherings. Front Public Health. 2022;10:1026489.

35. Knight J, Tan DHS, Mishra S. Maximizing the impact of limited vaccine supply under different early epidemic conditions: a 2-city modelling analysis of monkeypox virus transmission among men who have sex with men. CMAJ. 2022;194(46):E1560–E7.

36. Gan G, Janhavi A, Tong G, Lim JT, Dickens BL. The need for pre-emptive control strategies for mpox in Asia and Oceania. Infect Dis Model. 2024;9(1):214–23.

37. Joint Committee on Vaccination and Immunisation (JCVI). JCVI statement on mpox vaccination as a routine programme (https://www.gov.uk/government/publications/mpox-vaccination-programme-jcvi-advice-10-november/jcvi-statement-on-mpox-vaccination-as-a-routine-programme). 2023.

