## Supplementary Material for "Cost-Effectiveness of Vaccination Strategies to Control Future Mpox Outbreaks in England"

1. Statistics, Modelling and Economics, Data, Analytics & Surveillance, UK Health Security Agency, London, UK
2. Population Health Sciences, University of Bristol, Bristol, UK
3. Immunisation and Vaccine Preventable Diseases Division, UK Health Security Agency, London, UK
4. Blood Safety, Hepatitis, Sexually Transmitted Infections and HIV Division, UK Health Security Agency, London, UK
5. Institute for Global Health, University College London, London, UK
6. The National Institute for Health and Care Research Health Protection Research Unit in Blood Borne and Sexually Transmitted Infections at University College London in partnership with the UK Health Security Agency, London, United Kingdom
7. Health Protection Operations, UK Health Security Agency, London, UK
8. NHS ARDEN AND GREATER EAST MIDLANDS COMMISSIONING SUPPORT UNIT
9. People Delivery Unit, UK Health Security Agency
10. Northwest Health Protection Team, UK Health Security Agency, Manchester, UK
11. Regional Deputy Director, UKHSA South East
12. South London and Maudsley NHS Foundation Trust, UK.
13. Directorate of Emergency Preparedness, Resilience and Response, UK Health Security Agency, London, UK
14. Sheffield Teaching Hospitals NHS Foundation Trust, Sheffield, UK.
15. Croydon Health Services NHS Trust, Croydon, Surrey, UK
16. The National Institute for Health and Care Research Health Protection Research Unit in Behavioural Science and Evaluation at the University of Bristol in partnership with the UK Health Security Agency, Bristol, United Kingdom

*joint senior authors

### Supplementary I: Mpox Model and Equations

#### Mpox Transmission Model

The transmission model was started on 17 April 2022 (date of symptom onset for first mpox case in England (1)), with all individuals susceptible to mpox infection. When exposed under the force of infection ($\lambda$), individuals become latently infected (E, Exposed) but not yet infectious. After a latent period of $L$ days, they become infectious (I, infectious). Individuals recover and become immune (R, recovered) or are diagnosed and isolated after an effective infectious period of $d$ days. This effective infectious period, during which cases can transmit mpox, is shorter than the full infectious period due to isolation.

Two identical states of $E$ (exposure) and $I$ (infectious) are introduced so that the distributions for the latent period ($L$) and infectious period ($d$) follow gamma distributions of shape 2 rather than exponential distribution. This approach is based on empirical knowledge that these time intervals follow a bell-shaped distribution (2). A duration of immunity, $L_{\text{imm0}}$, reflects waning immunity induced by infection, which is the duration individuals stay in the recovered class before they become susceptible again. To model the potential difference in infectiousness and latent/infectious period induced by vaccination, the susceptible, exposed and infectious classes are further stratified by vaccination status: none, 1st dose, and 2nd dose.

The GBMSM population in England, assumed to be 769,000 individuals (3), is divided into four groups based on the risk of acquiring mpox (low and high-risk) and recent attendance at sexual health services (SHS) or not. Low and high-risk statuses were defined based on having fewer than 10 or at least 10 anal sex partners in last four months, comprising 11.9% and 88.1% of GBMSM, with mean contact rates of 22.3 and 1.6 per 4 months, respectively (4) . The 2021 RiiSH survey estimated that 36.8% of low-risk GBMSM and 69.3% of high-risk GBMSM attended SHS over the last 4 months recently. This results in the proportions in the following four groups as listed in Table S1.

**Table S1:** Proportion of GBMSM in low and high-risk groups and that attend SHS or not

| **Group (j)** | **Definition** | **Proportion** |
|---|---|---|
| **J=1** | Low-risk, not attending SHS | (1-0.119)×(1-0.368)=55.7% |
| **J=2** | Low-risk, attending SHS | (1-0.119) ×0.368 =32.4% |
| **J=3** | High-risk, not attending SHS | 0.119×(1-0.693) =3.7% |
| **J=4** | High-risk, attending SHS | 0.119×0.693=8.2% |

To capture differences in health care costs, productivity losses, and quality of life utilities for different severities of mpox disease, individuals infected with mpox were further categorised into three classes: severe (hospitalised, H), moderate (outpatient, M), and mild/minimal (not hospitalised, A) symptoms. Infected individuals progress to either H, M, or A with probabilities $p_1$, $p_2$, and $1-p_1-p_2$, respectively (Figure S1). The model also includes waning immunity from vaccination and recruitment of new GBMSM. To reflect the potential complexity of mpox natural history, 4×30 compartments are included in the model and definitions of model compartments and parameters are given in Table S2.

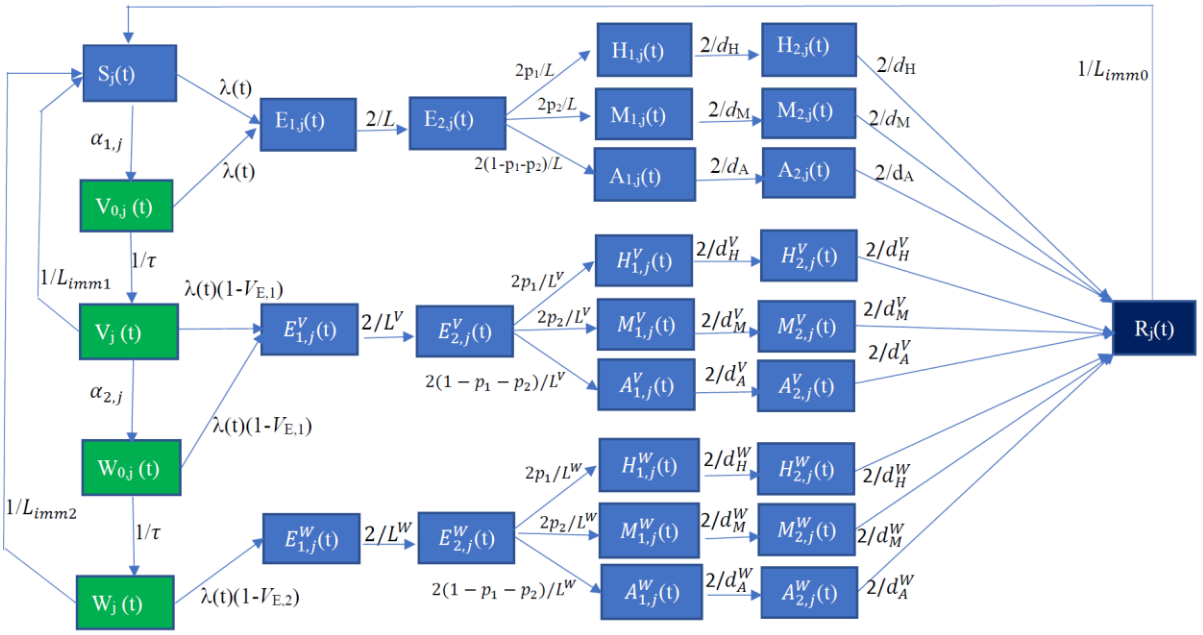

**Figure S1:** Schematic of the full mpox transmission model among GBMSM for assessing the impact and cost-effectiveness of different vaccination strategies. Stratifications by low and high risk for mpox infection and attendance at sexual health services are not shown.

**Table S2** Definitions of Variables and Parameters in Figure S1 and Equation S1. Subscript j denotes the risk level of an individual and whether they attend SHS or not (Table S1).

| **Variable** | **Definition** |
|---|---|
| $S_j$ | Number of the susceptible GBMSM in group *j* |
| $V_{0,j}$ | Number of those having 1st dose of vaccine but not yet being protected in group *j* |
| $V_j$ | Number of those having 1st dose of vaccine and being protected with $V_{E,1}$ in group *j* |
| $W_{0,j}$ | Number of those having 2nd dose of vaccine but being protected with $V_{E,1}$ in group *j* as $V_j$ |
| $W_j$ | Number of those having 2nd dose of vaccine and being protected with $V_{E,2}$ in group *j* |
| $E_{1,j}$ $(E^V_{1,j}, E^W_{1,j})$ | Number of exposed GBMSM in group *j* (first stage) (Corresponding number exposed after 1st and 2nd doses of vaccine) |
| $E_{2,j}$ $(E^V_{2,j}, E^W_{2,j})$ | Number of exposed GBMSM in group j (second stage) (Corresponding number exposed after 1st and 2nd doses of vaccine) |
| $H_{1,j}$ $(H^V_{1,j}, H^W_{1,j})$ | Number of severe inpatient hospitalised infections in group *j* (first stage) (Corresponding number of severe infections after 1st and 2nd doses of vaccine) |
| $H_{2,j}$ $(H^V_{2,j}, H^W_{2,j})$ | Number of severe inpatient hospitalised infections in group *j* (second stage) (Corresponding number of severe infections after 1st and 2nd doses of vaccine) |
| $M_{1,j}$ $(M^V_{1,j}, M^W_{1,j})$ | Number of moderate outpatient hospitalised infections in group *j* (first stage) (Corresponding number of moderate infections after 1st and 2nd doses of vaccine) |
| $M_{2,j}$ $(M^V_{2,j}, M^W_{2,j})$ | Number of moderate outpatient hospitalised infections in group *j* (second stage) (Corresponding number of moderate infections after 1st and 2nd doses of vaccine) |
| $A_{1,j}$ $(A^V_{1,j}, A^W_{1,j})$ | Number of mild and not hospitalised infections in group *j* (first stage) (Corresponding number of mild infections after 1st and 2nd doses of vaccine) |
| $A_{2,j}$ $(A^V_{2,j}, A^W_{2,j})$ | Number of mild and not hospitalised infections in group *j* (second stage) (Corresponding number of mild infections after 1st and 2nd doses of vaccine) |
| $R_j$ | Number of recovered infections or isolated infections in group *j* |

| Variable | Definition |
|---|---|
| **Parameter** | **Definition** |
| $\alpha$ | The recruitment rate assuming the age of GBMSM ranges from 15 to 49 years old (= $1/(365 \times (49 - 15))$ per day) |
| $\lambda_j$ | Force of infection for group *j* (see equation (S2)) |
| $\beta$ | Transmission coefficient of mpox |
| $V_{E,1}$, $V_{E,2}$ | Effectiveness of 1st and 2nd doses of vaccine against mpox infection (effect of vaccination on susceptibility) |
| $\xi_1$, $\xi_2$ | The relative infectiousness of infected individuals previously vaccinated with 1 and 2 doses relative to the unvaccinated infected individuals – set to one in model to assume no reduced infectivity |
| $\omega(t)$ | The relative contact rate which reduces during mpox outbreaks |
| $\alpha_{1,j}$, $\alpha_{2,j}$ | Vaccination rate for 1st dose and 2nd dose among group *j*. The actual allocations of vaccines up to the end of 2023 are given in Equation (S4) (i.e., $F_{1,j}(\alpha_{1,j};t)$, $F_{2,j}(\alpha_{2,j};t)$) and for the next 20 years they were given as equation (S5) below |
| $t_{vacc}$ | The discrete time over which vacciantions occur – they are allocated discretely each day |
| $\tau$ | Time duration from vaccination to when protected against mpox infection (14 days (5)) |
| $\Omega_j$ | Daily number of imported cases in group *j* |
| $t^{import}$ | The discrete time over which importation occur – they are allocated discretely each day |
| $L$, $L^V$, $L^W$ | Latent period for new infections amongst those that have either not been vaccinated or been vaccinated one or two times (assumed to be the same in model) |
| $d_H, d_H^V, d_H^W$ | Average infectious period for severe infections amongst those that have either not been vaccinated or been vaccinated one or two times (assumed to be the same in model) |
| $d_M, d_M^V, d_M^W$ | Average infectious period for moderate infections amongst those that have either not been vaccinated or been vaccinated one or two times (assumed to be the same in model) |
| $d_A, d_A^V, d_A^W$ | Average infectious period for mild infections amongst those that have either not been vaccinated or been vaccinated one or two times (assumed to be the same in model) |
| $p_1$ | Proportion of infections that are severe (6.1%) (6) |
| $p_2$ | Proportion of infections that are moderate (8.3%) (6) |
| $L_{imm0}$, $L_{imm1}$, $L_{imm2}$ | Duration of immunity induced by either infection, vaccination of 1 or 2 doses, respectively |

The model is approximated by a set of differential equations; for SHS and sexual activity groups *j* (=1,2,3,4), they are

$$\frac{dS_j(t)}{dt} = -\lambda_j(t)S_j(t) - \Omega_j(t)\delta\left(t - t^{import}\right) - \delta(t - t_{vacc})F_{1,j}\left(\alpha_{1,j};t\right) + \alpha(N_j - S_j(t)) + \frac{V_j(t)}{L_{imm1}} + \frac{W_j(t)}{L_{imm2}} + \frac{R_j(t)}{L_{imm0}}$$

$$\frac{dV_{0,j}(t)}{dt} = -\lambda_j(t)V_{0,j}(t) + \delta(t - t_{vacc})F_{1,j}\left(\alpha_{1,j};t\right) - (\alpha + \tfrac{1}{\tau})V_{0,j}(t)$$

$$\frac{dV_j(t)}{dt} = -\lambda_j(t)\left(1 - V_{E,1}\right)V_j(t) + \tfrac{1}{\tau}V_{0,j}(t) - \delta(t - t_{vacc})F_{2,j}\left(\alpha_{2,j};t\right) - \alpha V_j(t) - \frac{V_j(t)}{L_{imm1}}$$

$$\frac{dW_{0,j}(t)}{dt} = -\lambda_j(t)\big(1 - V_{\mathrm{E},1}\big)W_{0,j}(t) + \delta(t - t_{vacc})F_{2,j}\big(\alpha_{2,j}; t\big) - (\alpha + \tfrac{1}{\tau})W_{0,j}(t)$$

$$\frac{dW_j(t)}{dt} = -\lambda_j(t)\big(1 - V_{\mathrm{E},2}\big)W_j(t) + \frac{1}{\tau}W_{0,j}(t) - \alpha W_j(t) - \frac{W_j(t)}{L_{imm2}}$$

$$\frac{dE_{1,j}(t)}{dt} = \lambda_j(t)[S_j(t) + V_{0,j}(t)] - \frac{2}{L}E_{1,j}(t) - \alpha E_{1,j}(t)$$

$$\frac{dE_{2,j}(t)}{dt} = \frac{2}{L}E_{1,j}(t) - \frac{2}{L}E_{2,j}(t) + \Omega_j(t)\delta(t - t^{import}) - \alpha E_{2,j}(t)$$

$$\frac{dA_{1,j}(t)}{dt} = \frac{2(1-p_1-p_2)}{L}E_{2,j}(t) - \frac{2}{d_A}A_{1,j}(t) - \alpha A_{1,j}(t)$$

$$\frac{dA_{2,j}(t)}{dt} = \frac{2}{d_A}A_{1,j}(t) - \frac{2}{d_A}A_{2,j}(t) - \alpha A_{2,j}(t)$$

$$\frac{dM_{1,j}(t)}{dt} = \frac{2p_2}{L}E_{2,j}(t) - \frac{2}{d_M}M_{1,j}(t) - \alpha M_{1,j}(t)$$

$$\frac{dM_{2,j}(t)}{dt} = \frac{2}{d_M}M_{1,j}(t) - \frac{2}{d_M}M_{2,j}(t) - \alpha M_{2,j}(t)$$

$$\frac{dH_{1,j}(t)}{dt} = \frac{2p_1}{L}E_{2,j}(t) - \frac{2}{d_H}H_{1,j}(t) - \alpha H_{1,j}(t)$$

$$\frac{dH_{2,j}(t)}{dt} = \frac{2}{d_H}H_{1,j}(t) - \frac{2}{d_H}H_{2,j}(t) - \alpha H_{2,j}(t)$$

$$\frac{dE^V_{1,j}(t)}{dt} = \lambda_j(t)\big(1 - V_{E,1}\big)[V_j(t) + W_{0,j}(t)] - \frac{2}{L^V}E^V_{1,j}(t) - \alpha E^V_{1,j}(t)$$

$$\frac{dE^V_{2,j}(t)}{dt} = \frac{2}{L^V}E^V_{1,j}(t) - \frac{2}{L^V}E^V_{2,j}(t) - \alpha E^V_{2,j}(t)$$

$$\frac{dA^V_{1,j}(t)}{dt} = \frac{2(1-p_1-p_2)}{L^V}E^V_{2,j}(t) - \frac{2}{d^V_A}A^V_{1,j}(t) - \alpha A^V_{1,j}(t)$$

$$\frac{dA^V_{2,j}(t)}{dt} = \frac{2}{d^V_A}A_{1,j}(t) - \frac{2}{d^V_A}A^V_{2,j}(t) - \alpha A^V_{2,j}(t) \qquad \text{(S1)}$$

$$\frac{dM^V_{1,j}(t)}{dt} = \frac{2p_2}{L^V}E^V_{2,j}(t) - \frac{2}{d^V_M}M^V_{1,j}(t) - \alpha M^V_{1,j}(t)$$

$$\frac{dM^V_{2,j}(t)}{dt} = \frac{2}{d^V_M}M^V_{1,j}(t) - \frac{2}{d^V_M}M^V_{2,j}(t) - \alpha M^V_{2,j}(t)$$

$$\frac{dH^V_{1,j}(t)}{dt} = \frac{2p_1}{L^V}E^V_{2,j}(t) - \frac{2}{d^V_H}H^V_{1,j}(t) - \alpha H^V_{1,j}(t)$$

$$\frac{dH^V_{2,j}(t)}{dt} = \frac{2}{d^V_H}H^V_{1,j}(t) - \frac{2}{d^V_H}H^V_{2,j}(t) - \alpha H^V_{2,j}(t)$$

$$\frac{dE^W_{1,j}(t)}{dt} = \lambda_j\big(1 - V_{\mathrm{E},2}\big)W_j(t) - \frac{2}{L^W}E^W_{1,j}(t) - \alpha E^W_{1,j}(t)$$

$$\frac{dE^W_{2,j}(t)}{dt} = \frac{2}{L^W}E^W_{1,j}(t) - \frac{2}{L^W}E^W_{2,j}(t) - \alpha E^W_{2,j}(t)$$

$$\frac{dA^W_{1,j}(t)}{dt} = \frac{2(1-p_1-p_2)}{L^W}E^W_{2,j}(t) - \frac{2}{d^W_A}A^W_{1,j}(t) - \alpha A^W_{1,j}(t)$$

$$\frac{dA^W_{2,j}(t)}{dt} = \frac{2}{d^W_A}A^W_{1,j}(t) - \frac{2}{d^W_A}A^W_{2,j}(t) - \alpha A^W_{2,j}(t)$$

$$\frac{dM^W_{1,j}(t)}{dt} = \frac{2p_2}{L^W}E^W_{2,j}(t) - \frac{2}{d^W_M}M^W_{1,j}(t) - \alpha M^W_{1,j}(t)$$

$$\frac{dM_{2,j}^W(t)}{dt} = \frac{2}{d_M^W} M_{1,j}^W(t) - \frac{2}{d_M^W} M_{2,j}^W(t) - \alpha M_{2,j}^W(t)$$

$$\frac{dH_{1,j}^W(t)}{dt} = \frac{2p_1}{L^W} E_{2,j}^W(t) - \frac{2}{d_H^W} H_{1,j}^W(t) - \alpha H_{1,j}^W(t)$$

$$\frac{dH_{2,j}^W(t)}{dt} = \frac{2}{d_H^W} H_{1,j}^W(t) - \frac{2}{d_H^W} H_{2,j}^W(t) - \alpha H_{2,j}^W(t)$$

$$\frac{dR_j(t)}{dt} = \frac{2}{d_H} H_{2,j}(t) + \frac{2}{d_H^V} H_{2,j}^V(t) + \frac{2}{d_H^W} H_{2,j}^W(t) + \frac{2}{d_M} M_{2,j}(t) + \frac{2}{d_M^V} M_{2,j}(t) + \frac{2}{d_M^W} M_{2,j}^W(t) + \frac{2}{d_A} A_{2,j}(t) + \frac{2}{d_A^V} A_{2,j}^V(t) + \frac{2}{d_A^W} A_{2,j}^W(t) - (\alpha + \frac{1}{L_{immo}}) R_j(t)$$

**Force of infection:** In equation (S1), $\lambda_j(t)$ is the force of infection for group j and given by:

$$\lambda_j(t) = \beta\omega(t) \sum_{i=1}^{4} \Pi_{j,i} \, [A_{1,i}(t) + A_{2,i}(t) + M_{1,i} + M_{2,i}(t) + H_{1,i}(t) + H_{2,i}(t) + \xi_1(A_{1,i}^V(t) + A_{2,i}^V(t) + M_{1,i}^V + M_{2,i}^V(t) + H_{1,i}^V(t) + H_{2,i}^V(t)) + \xi_2(A_{1,i}^W(t) + A_{2,i}^W(t) + M_{1,i}^W + M_{2,i}^W(t) + H_{1,i}^W(t) + H_{2,i}^W(t))] \quad \text{(S2)}$$

Here, $\beta$ is the transmission coefficient, $\xi_j$ is the relative infectiousness of vaccinated infected individuals relative to the unvaccinated infected individuals (assumed to be one in model), and $\omega(t)$ represents the relative contact rate which reduces during mpox outbreaks.

In Equation (S2), the matrix $\Pi_{j,i}(t) = \boldsymbol{M}_{j,i}/\Lambda$ is a scaled version of the mixing matrix, with $\Lambda$ being the dominant eigenvalue of the next generation matrix $\boldsymbol{M}^*$, whose element $(j,i)^{th}$ is given by $M_{j,i}^* = N_j M_{j,i}$, where $N_j$ is the population size within group $j$ (7) . $\beta\Pi_{j,i}$ gives the infection pressure exerted on a susceptible individual within group $j$ by a single infectious individual in group $i$. The baseline matrix among four groups (Table S1) is based on anal sex contact data from the 2021 RiiSH survey(4),

$$\boldsymbol{M} = M_{Risk} \otimes M_{SHS} = \begin{pmatrix} 0.070 & 0.363 & 0.181 & 0.936 \\ 0.067 & 0.366 & 0.172 & 0.945 \\ 0.882 & 4.562 & 2.731 & 14.125 \\ 0.838 & 4.606 & 2.596 & 14.260 \end{pmatrix},$$

#### Responsive behavioural change

To account for behavioural changes during an outbreak, we assume that the induced public health response will result in a reduction in sexual contact rates ($\omega(t)$) and the effective infectious period ($d$(t)) as occurred in the 2022 outbreak(7). That is, both $\omega(t)$ and $d(t)$ reduce in the form of a sigmoidal function with the relative sexual contact rate $\omega$ decreasing from 100% when outbreak is declared to $\omega_o$=55.5% (95%CrI: 44.2–75.1%), and the effective infectious period $d$ reducing from $D_1$ = 3.0 (95%CrI: 2.5–3.9) to $D_2$ = 2.4 (95%CrI: 2.0–3.3) days. Both parameters stay at these reduced values until the outbreak ends, after which they return to their baseline values as MPOX-RiiSH data suggests occurred in England(8). The formula for ω(t) and d(t) are given below:

$$\omega(t) = \begin{cases} 1 & when\, t < 0 \\ 1 + \frac{(\omega_O - 1)}{1+\exp(-(t-t_c)C))} & otherwise \end{cases} \quad \text{(S3A)}$$

and the infectious period shortens as

$$d(t) = \begin{cases} D_1 & when\, t < 0 \\ D_1 + \frac{(D_2 - D_1)}{1+\exp(-(t-t_c)C))} & otherwise \end{cases} \quad \text{(S3B)}$$

with the midpoint of change at day $t_c$= 51 (95%CrI: 40–69) from the day of outbreak alert, and speed of reduction $C$= 0.016 (95%CrI: 0.011–0.039) per day (estimated through calibrating model to the outbreak data up to 12 August 2022 (see (7); Table 1).

#### Vaccination

##### Vaccination up to end 2023

Up to 31st December 2023, vaccines were distributed to GBMSM susceptible for mpox for 1st dose or those that were singly vaccinated for their 2nd dose (9). Vaccines were allotted proportionately among the two high risk groups ($j$=3,4), with there being specific rates $\alpha_{1,j}(t)$ for 1st dose and $\alpha_{2,j}(t)$ for 2nd dose, estimated from the weekly vaccination data (Figure S2 (9)) which were assumed to be evenly delivered over each day of one week. This implies that on day $t$ (=1,2,3,…,624), the first doses of vaccine are delivered among the susceptible of two high risk groups as:

$$F_{1,j}(\alpha_{1,j};t) = 0, j = 1,2$$
$$F_{1,j}(\alpha_{1,j};t) = \frac{\alpha_{1,j}}{S_3(t)+S_4(t)} S_j(t), j = 3,4 \quad \text{(S4A)}$$

And the second doses of vaccines are delivered among those singly vaccinated as

$$F_{2,j}(\alpha_{2,j};t) = 0, j = 1,2$$
$$F_{2,j}(\alpha_{2,j};t) = \frac{\alpha_{2,j}}{V_3(t)+V_4(t)} V_j(t), j = 3,4 \quad \text{(S4B)}$$

Notice that day 624 represents the date 31 December 2023 from 17 April 2022.

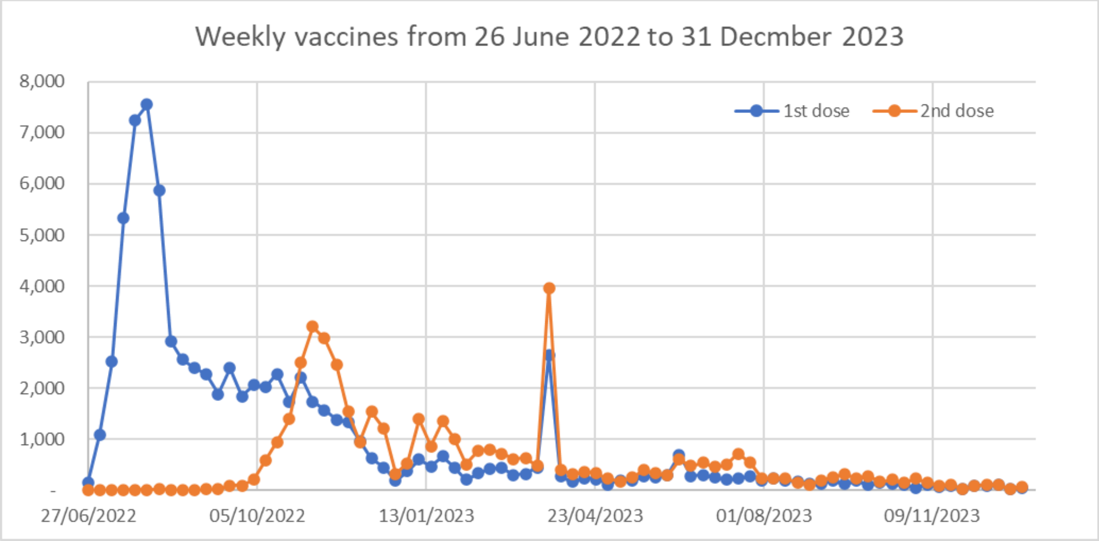

**Figure S2:** Weekly number of vaccines delivered from 27 June 2022 to 31 December 2023 in England.

##### Vaccination scenarios from start 2024

To assess the impact and cost-effectiveness of various vaccination strategies, we consider the following three scenarios:

1) **No Vaccination:** There are no vaccinations from 1 January 2024 (day 625) to 31 December 2043 (day 7,929). This is regarded as the counterfactual scenario:

$$\alpha_{1,j}(t) = \alpha_{2,j}(t) = 0, j = 1,2,3,4; t = 625, \ldots, 7929$$

In this scenario, a reduction in risk behaviour and the effective infectious period still occurs during each outbreak as observed in 2022.

2) **Pre-emptive Vaccination (Constant rate to high risk GBMSM attending SHS):** Continuous current vaccination of high-risk SHS attendees (group $j$=4) at constant daily rates: $\alpha_{1,j}$ for 1st dose and $\alpha_{2,j}$ for 2nd dose per day. Let $Const_V$ denote the total daily number of vaccines given and $q$ the assumed proportion of 2nd doses (0.561 estimated from data up to December 2023), then vaccines will be delivered during the period from $t$ = 625 (1 January 2024) to t=7929 (31 December 2043) as follows:

$$F_{1,4}(t) = \alpha_{1,4} \equiv \frac{Const_V}{1+q}, \text{if } S_4(t) \geq \frac{Const_V}{1+q}$$
$$F_{1,4}(t) = \alpha_{1,4} \equiv S_4(t), \text{if } 0 \leq S_4(t) < \frac{Const_V}{1+q} \quad \text{(S5A)}$$

$$F_{2,4}(t) = \alpha_{2,4} \equiv Const_V - \alpha_{1,4}, \text{if } V_4(t) \geq Const_V - \alpha_{1,4}$$
$$F_{2,4}(t) = \alpha_{2,4} \equiv V_4(t), \text{if } 0 \leq V_4(t) < Const_V - \alpha_{1,4} \quad \text{(S5B)}$$

To manage the saturation of population vaccination, the following rule is enforced here: 1st doses of vaccines will be given out only if there are susceptible individuals in group 4; 2nd doses of vaccines will be given out if not all the members of group 4 have been doubly vaccinated. If first doses are saturated, then the excess vaccines will be given out as second doses until these are saturated. Because vaccination may be saturated, it is expected that the total number of vaccines given out over 20 years could be less than $Const_V$*(7929-624) and the actual proportion having 2nd doses of vaccines may differ from $q$=56.1%. Indeed, projections show that vaccination will saturate when the rate is above some specific rate (41 doses per day in baseline scenario), with this rate being higher if the duration of protection is shorter (75 doses per day if assume 2.5/5 year protection after 1/2 doses) or the rate at which new individuals join the high-risk GBMSM population is higher (81 doses per day if assume larger high-risk group – sensitivity analysis 11). Note that the first subscript in $F_{1,j}(t)$ and $F_{2,j}(t)$ refers to first or second dose and the second subscript refers to the sub-group of GBMSM (SHS and risk class; Table S1).

3) **Reactive Vaccination (Constant rate to high risk GBMSM attending SHS)**: Reactive vaccination of GBMSM SHS attendees at high risk for mpox delivered at a constant rate of $Const_V$ per day only when an outbreak is signalled. When an outbreak response starts, the vaccines will be given as in equation (S5), otherwise vaccination will stop.

For the reactive vaccination scenarios, it is assumed that vaccines will only be delivered if an outbreak response occurs. The threshold for the start of an outbreak response was decided through consultation with UKHSA as 120 indigenously generated cases within three months. The threshold for the end of an outbreak response was decided as ≤60 indigenous case over three months. Low (20% lower) and High (20% higher) thresholds were assumed in sensitivity analyses.

The total vaccines given out up to 31 December 2023 is 124,464, among which 79,743 were for 1st dose and 44,721 were for 2nd dose so ($q$=) 56.1% got 2nd dose of vaccines (9). The mean daily vaccination rates from July to December 2022 (regarded as the outbreak period) was 465.4 per day (354.4 for 1st dose and 111.0 for 2nd dose). This daily rate $Const_V$ = 465.4 per day was defined as the high rate in our future reactive vaccination scenario to consider the early ramp up in delivery and access to vaccines. Other vaccination rates were also investigated to identify the most cost-effective vaccination programs. For reactive vaccination initiatives, this included vaccination rates of 27, 41, 81, 135, 204, doses per day, which correspond to achieving a vaccination coverage of 10%, 15%, 30%, 50% and 75% after a year if there had been no prior vaccination. For pre-emptive vaccination initiatives, vaccination rates of 13, 27, 41, 54, 81, 135 doses per days were modelled, which correspond to achieving a vaccination coverage of 5%, 10%, 15%, 20%, 30% and 50% after a year. Figure S4 shows the annual vaccinations under the two schemes.

**Duration of Immunity**

Uncertainty in the duration of vaccine protection is an ongoing issue because it is a relatively new widescale deployment of a vaccine, and so no robust published data beyond 2 years exist, although longer term follow up studies including looking antibody responses post infection vs vaccination (including 1 vs 2 vaccine doses) are underway.

A short duration of protection was assumed in our previous study (7) because of recent outbreak data in France and USA suggesting that many of the new cases are among double vaccinated individuals (10, 11). However, the fact that no significant outbreaks occurred in 2023 in England (12) suggests that the duration of protection could be long. A recent theoretical investigation (13) indicates that MVA-BN VE will remain >59% protective for up to 10 years even after a single dose. Based on this information, we considered the following three choices (Table S3) for the duration of protection following vaccination and natural infection, with the medium duration of protection being assumed in the baseline scenario and the others being used in the sensitivity analyses.

**Table S3**: Duration of protection following vaccination and natural infection

| **Duration** | **$L_{imm1}$ (induced by one dose)** | **$L_{imm2}$ (induced by two doses)** | **$L_{imm0}$ (induced by natural infection)** |
|---|---|---|---|
| Short | 2.5 years | 5 years | 5 years |
| Medium (baseline) | 5 years | 10 years | 10 years |
| Long | 10 years | 20 years | 20 years |

**Including Costs and Health benefits**

**Health benefits**

The health-related quality of life (HRQoL) for individuals infected with mpox is categorised into three categories:

- Mild/minimal symptoms (A): Managed at home, not hospitalised.
- Moderate symptoms (M): Managed as outpatients, not admitted to the hospital.
- Severe symptoms (H): Require hospitalisation, with some needing intensive care.

Utility weights for uninfected GBMSM and for those GBMSM with mild, moderate, and severe mpox are listed in **Table S12 in Supplementary II – Estimates of Costs and Utilities**. These weights were used to calculate the Quality Adjusted Life Years (QALYs) over 20 years. The total QALY for each year was calculated by summing the product of the number of cases with each level of disease and their respective utility weights, and adding to that the product of the number of uninfected GBMSM and the utility weight for healthy individuals. The formula for accumulated QALYs over 20 years is:

$$QALYs = \sum_{y=0,1,\ldots,19} \frac{UW_H H_y + UW_M M_y + UW_A A_y + UW_0(N - H_y - M_y - A_y)}{(1+\theta)^y} \quad \text{(S6)}$$

where $\theta$ =0.035 is the baseline annual discount rate (14, 15) and $N$ =769,000 represents the total GBMSM population in England. $A_y$, $M_y$ and $H_y$ are the number of those GBMSM with mild, moderate, and severe mpox in year $y$, and $UW_A$, $UW_M$ and $UW_H$ are their respective utility weights (Table S12). $N-H_y-M_y-A_y$ is the number of uninfected GBMSM and $UW_0$ is their utility weight.

**Productivity losses**

Based on a recent survey (RiiSH) among GBMSM (8), productivity losses due to mpox were estimated (Details are given in **Supplementary II – Estimates of Costs and Utilities**). The formula to calculate total productivity losses (PL) over 20 years is:

$$PL = \sum_{y} \frac{(A_y PL_A + M_y PL_M + H_y PL_H)}{(1+\theta)^y}, \; y \in 0,1,\ldots,19 \quad \text{(S7)}$$

Here, $PL_A$, $PL_M$, $PL_H$ represent the per capita productivity losses for mild, moderate, and severe cases of mpox, respectively.

**Costs of clinical case managements (CCM)**

The total costs of CCM over the 20 years are calculated as:

$$Cost_{CCM} = \sum_{y=0}^{19} \frac{U_A A_y + U_M M_y + U_H H_y}{(1+\theta)^y} \quad \text{(S8)}$$

Here, $U_A$, $U_M$, $U_H$ represent the per capita costs for mild, moderate, and severe cases of mpox, respectively. The details of estimation are given in **Supplementary II – Estimates of Costs and Utilities**.

**Vaccination costs**

The total costs due to vaccination over 20 years are calculated as:

$$Cost_{\text{Vacc}} = \sum_{y=0}^{19} \frac{(V_{\text{price}} + P_{adm1})T_{1,y} + (V_{\text{price}} + P_{adm2})T_{2,y}}{(1+\theta)^y}$$
$$= \sum_{y=0}^{19} \frac{P_{adm1}T_{1,y} + P_{adm2}T_{2,y}}{(1+\theta)^y} + V_{\text{price}} \sum_{y=0}^{19} \frac{T_{1,y} + T_{2,y}}{(1+\theta)^y}$$

(S9)

where $T_{1,y}$ and $T_{2,y}$ are the number of 1st and 2nd doses administered each year, and $P_{adm1}$ and $P_{adm2}$ are the administration costs for each dose. For the baseline projections, the vaccine price $V_{price}$ was assumed to be same as the new Shingrix vaccine (16) due to confidentiality over the real cost of the mpox vaccine. Costing details are given in **Supplementary II – Estimates of Costs and Utilities.**

**Public Health Responses costs (PHR)**

Public health response costs include contact tracing costs, related responses for each case including training and coordination ($Cost_{Case}$), and fixed outbreak costs for the public health response, including strategic costs such as for incident management teams and cells ($Cost_{Duration}$). These are summed up as PHR costs:

$Cost_{PHR} = Cost_{Duration} + Cost_{Case}$,

with:

$$Cost_{Duration} = \sum_{y=0}^{19} \frac{C_{week} \times (D_{OTB,y}/7)}{(1+\theta)^y} \quad \text{(S10A)}$$

$$Cost_{Case} = \sum_{y=0}^{19} \frac{(A_y + M_y + H_y) C_{Case}}{(1+\theta)^y} \quad \text{(S10B)}$$

where $D_{OTB,y}$ is the number of days of outbreaks, $(A_y+M_y+H_y)$ is the total number of cases during outbreaks within year y, $C_{Week}$ is the weekly cost, and $C_{Case}$ is the cost per case estimated from the public health response in the 2022 outbreak. Details are given in **Supplementary II – Estimates of Costs and Utilities.**

**Probabilistic sensitivity analysis**

Model parameters, including vaccine efficacy ($V_{E,1}$, $V_{E,2}$), utility weights ($UW_A$, $UW_M$, $UW_H$), CCM costs ($U_A$, $U_M$, $U_H$), and PHR costs ($C_{week}$, $C_{case}$), were varied across their uncertainty ranges. Triangular distributions were chosen to describe the uncertainties using R function rtriangle(n,a,b,c) to generate *n* random variables that has median c and a and b as its lower and upper levels. Five-hundred samples of these parameters were randomly sampled. These samples were further combined with the best 500 sets of transmission parameters obtained in our previous model calibration to the 2022 outbreak data (7) to investigate the uncertainties in the total costs and benefits.

**Cost-effectiveness of vaccination strategies compared to no vaccination**

The total costs of mpox outbreaks under vaccination include vaccination costs ($Cost_{Vacc}$), costs for clinical case management (CCM), and public health response (PHR) costs:

$$C = Cost_{Vacc} + Cost_{CCM} + Cost_{PHR} \quad \text{(S11)}$$

The effectiveness of control measures is measured by QALYs gained:

$$E = QALYs. \quad \text{(S12)}$$

The incremental cost-effectiveness ratio (ICER) is calculated as:

$$ICER = (C_A - C_B)/(E_A - E_B) \quad \text{(S13)}$$

where A is the vaccination scenario and B is the no-vaccination scenario. An option is deemed cost-effective if the median ICER is below the willingness to pay threshold of £20,000/QALY, with 90% of ICER projections below £30,000/QALY.

Two other measurements for the total costs were considered in sensitivity analysis: one ignores the contribution from PHR so the total costs are calculated as

$$C = Cost_{\mathrm{Vacc}} + Cost_{\mathrm{CCM}} \tag{S14}$$

The second one includes productivity losses (PL) in the total costs as,

$$C = Cost_{\mathrm{Vacc}} + Cost_{\mathrm{CCM}} + Cost_{\mathrm{PHR}} + PL \tag{S15}$$

#### Supplementary II: Estimates of Costs and Utilities

##### Overall Costing Approach

We gathered cost data from a variety of sources, including micro-costing where data were available, and referring to the National Schedule of NHS costs and published literature. We aimed to estimate the following unit costs to be used in the transmission model of mpox in the England (Table  S4):

**Table S4:** Costing Methods Used for Mpox cost-effectiveness analysis

| **Cost item** | **Source/Method** |
|---|---|
| Clinical case management costs: mild case | Sexual Health services tariff for mpox (17), assuming mild cases are diagnosed and treated in sexual health clinics; British National Formulary for medication costs (16) |
| Clinical case management costs: moderate case | National Schedule of NHS costs (18), Unit Costs of Health and Social Care (19), assuming moderate cases attend A&E but are not admitted to hospital |
| Clinical case management costs: severe case | National Schedule of NHS costs (18), assuming severe cases receive hospital inpatient care |
| Vaccine administration and vaccine costs | Sexual Health services tariff for mpox (17) and BNF NICE (16) |
| Public health response costs | UKHSA staffing costs and response costs from three regions |
| Productivity losses | RiiSH-MPOX survey among GBMSM(8) and Office of National Statistics employment and salary information (20, 21) |

In addition, we gathered data on health-related quality of life (HRQoL) in order to estimate QALY weights for the healthy population, and mild, moderate, and severe cases of mpox. The costing analysis was conducted from a health system perspective with costs presented in 2022 British Pounds (£). The proportion of mpox cases at each level of severity was calculated from Secondary Uses Service (SUS), the Emergency Care Data Set (ECDS), and clinical evidence (6). Clinical information was based on the UK standard guidance and discussions with clinical experts (22-24).

##### Clinical case management costs

*Mild cases*

Patients with mild or minimal symptoms were assumed to be diagnosed and receive care through sexual health services, and isolated at home. Mild symptoms typically presented with a few lesions. For patients experiencing pain, medications such as codeine in combination with paracetamol and local anaesthetic gel for lesion pain were administered. Additionally, fever relief medications such as paracetamol or ibuprofen were prescribed for patients with fever(25). All cases were confirmed with PCR by Rare and Imported Pathogens Laboratory (RIPL) as part of the testing and physical examination process.

The costs for sexual health services were derived from the Tariff Calculator for mpox activities in Sexual Health Services using the Integrated Sexual Health Tariff (ISHT) Methodology Spreadsheet (17). Details are shown in Table S5. Activities included initial phone consultation or triage, mpox test and physical exam, and phone call with positive results. In addition, the costs of medications were included, with unit costs for each medication being drawn from the British National Formulary (BNF) Tariff (26).

Uncertainty around these overall costs for care of mild cases relates primarily to the staff costs, depending on what level of staff are interacting with patients. To account for this, we varied the overall cost of care per mild case by +/- 20%, keeping medication costs constant. Staff cost estimates were based on expert opinion of resources used, and are not estimates that can be used to construct payment mechanisms between providers and commissioners of services.

**Table S5:** The Costs of a Mild Mpox Case Diagnosed in Sexual Health Services and Medications

| **Item** | **Unit cost (£, 2022)** | **Proportion receiving this activity** | **Cost per case (£, 2022)** | **Ref** |
|---|---|---|---|---|
| Phone consultation | 25.11 | 0.95 | 23.86 | (17) |
| Walk-in triage | 25.11 | 0.05 | 1.26 | (17) |
| Phone follow-up on positive results | 45.32 | 1.00 | 45.32 | (17) |
| Mpox test and physical examination | 306.63 | 1.00 | 306.63 | (17) |
| *Total activity costs:* | | | 377.06 | |
| Med: Codeine + paracetamol | 3.40 | 0.06 | 0.19 | (26),CE |
| Med: Local anaesthetic gel | 10.50 | 0.12 | 1.22 | (26),CE |
| Med: Paracetamol or Ibuprofen | 0.30 | 0.52 | 0.16 | (26),CE |
| *Total medication costs:* | | | 1.57 | |
| **Cost per case in SHS** | | | **378.63** | |
| **Lower bound assumption (activity -20%)** | | | **303.22** | |
| **Upper bound assumption (activity +20%)** | | | **454.04** | |

**Abbreviations:** SHS, sexual health services; MPX, mpox; Med, medication; A, assumption; CE, clinical expert.

*Moderate cases*

Patients with a moderate level of symptoms were defined as those who attended hospital through the emergency department (ED) and were not admitted. The costs of ED visits are presented in **Table S6.**

To determine the cost of ED visits, we referred to the National schedule of NHS costs (18), and calculated the weighted average estimates per visit. The NHS reference costs provided procurement currency codes or Healthcare Resource Groups (HRGs), which were estimated based on the setting and complexity of cases. The cost of the entire service received is bundled in the estimates (18) . No specific costs for mpox are available, so we included the weighted average of currency codes for emergency medicine, category 3 investigation with category 1-4 treatment (VB02Z, VB03Z), under service code T01NA for non-admitted patients to type 1 facilities (emergency departments) (details are presented in Table S7).

Based on the sexual health services tariff (17), we estimate 14.4% of patients attended sexual health services prior to attending ED. As an alternative costing source, we used the reported cost of a day case for a genitourinary medicine infection from the report on Unit Costs of Health and Social Care 2022 (19). This gave a lower bound cost estimate.

**Table S6:** Cost of a Moderate Mpox Case Based on Emergency Department Visit Costs

| Item | Unit cost (£, 2022) | Proportion receiving this activity | Cost per case (£, 2022) | Ref |
|---|---|---|---|---|
| Weighted average for emergency care (non-admitted) | 673.65 | 1 | 673.65 | (18, 19) |
| Transferred from sexual health services: Activity costs from Table 1 plus £30.22 for referral | 407.29 | 0.144 | 57.02 | (17) |
| **Baseline total cost per emergency visit, non-admitted (baseline and upper bound)** | | | **730.67** | |
| **Alternative cost for day case for GUM infection (lower bound)** | | | **577** | (19) |

**Abbreviations:** GUM, genitourinary medicine.

**Table S7:** NHS Currency Code and Description Used to Calculate Hospitalisation Costs (18)

| Index | Code | Description |
|---|---|---|
| Emergency department | VB02Z | Emergency Medicine, Category 3 Investigation with Category 4 Treatment |
| | VB03Z | Emergency Medicine, Category 3 Investigation with Category 1-3 Treatment |
| Inpatient care | WJ04Z | Genito-Urinary Medicine (GUM) Infections |
| | WJ01A | Complex Infectious Diseases with Multiple Interventions |
| | WJ02A | Major Infectious Diseases with Multiple Interventions |
| | WJ01B | Complex Infectious Diseases with Single Intervention |
| | WJ02B | Major Infectious Diseases with Single Intervention |
| | WJ01C | Complex Infectious Diseases without Interventions, with CC Score 6+ |
| | WJ02C | Major Infectious Diseases without Interventions, with CC Score 6+ |
| | WJ01D | Complex Infectious Diseases without Interventions, with CC Score 3-5 |
| | WJ02D | Major Infectious Diseases without Interventions, with CC Score 3-5 |
| | WJ01E | Complex Infectious Diseases without Interventions, with CC Score 0-2 |
| | WJ02E | Major Infectious Diseases without Interventions, with CC Score 0-2 |
| Critical care | CCU01 | Non-specific, general adult critical care PATIENTS predominate, 0 Organs Supported |
| | CCU02 | Surgical adult PATIENTS (unspecified specialty), 0 Organs Supported |
| | CCU03 | Medical adult PATIENTS (unspecified specialty), 0 Organs Supported |
| | CCU90 | Non-standard LOCATION using a WARD area, Adult Critical Care, 0 Organ Supported |
| | CCU91 | Non-standard LOCATION using the operating department, Adult Critical Care, 0 Organ Supported |

*Severe cases*

Patients with severe mpox symptoms were defined as those who received inpatient hospital care. Based on the findings reported in Fink et al (2022) regarding disease complications, rectal and perianal pain was a common indication for hospital admission (27). Furthermore, the symptoms could progress into more complex conditions, with some patients requiring surgery and some patients experiencing secondary bacterial infections. The average duration of hospital stay for mpox cases was 5.5 days (IQR 1-8) based on data from SUS and ECDS (6). We estimated the unit cost of hospitalisation by using a range of HRG codes including major infectious diseases and complex infectious diseases, and assuming only non-elective hospitalisations apply. We also incorporated the HRG code for Genitourinary Medicine (GUM) infections in the unit cost analysis and reported a weighted average as the base case cost per hospitalisation. Based on this assumption, we used the HRG codes WJ04Z, WJ01/02/03A, WJ01/02B, WJ01/02C, WJ01/02D, and WJ01/02E to estimate the costs associated with hospitalisation. We included the costs for both short and long stay non-elective inpatients (18).

Based on analysis of SUS and ECDS data, 63.4% of patients admitted to hospital for mpox treatment first attended ED prior to admission (6). We assume the remainder (36.6%) attended SHS prior to hospital admission.

Approximately 4% of hospitalised patients were assumed to require intensive care according the data from SUS:ECDS (6). Based on the evidence presented in Fink et al (2022), it was found that all mpox cases in the ICU did not require organ support (27) . Accordingly, we estimated the ICU costs based on the National NHS Cost using the Critical Care (CC) sheet and the following HRG codes CCU01, CCU02, CCU03, CCU90, and CCU91 (18) .

During the outbreak period until June 2022, mpox was classified as a high consequence infectious disease (HCID). This classification necessitated the consideration of additional costs, including the costs associated with special isolation rooms, personal protective equipment (PPE), and staff training respond to the outbreaks. As of January 2023, HCID care is still needed for cases due to Clade I MPXV, while the recent outbreak was due to Clade II (22) . We assume that future cases will not require HCID care.

We also use an alternative estimate of the hospitalisation cost for GUM infections from the Personal Social Services Research Unit (PSSRU) for sensitivity analysis. This gave us a lower bound cost estimate (19) .

The costs of inpatient care for mpox are presented in Table S8, and details on NHS currency code and description used to calculate hospitalisation costs are provided in Table S7.

**Table S8:** Hospitalisation Costs Weighted Averages

| **Parameter/Activity** | **Unit cost (£, 2022)** | **Proportion** | **Cost per case (£, 2022)** | **Ref** |
|---|---|---|---|---|
| Weighted average for long stay non-elective inpatients | 4,803.73 | | | (18) |
| Weighted average for short stay non-elective inpatients | 731.80 | | | (18) |
| Weighted average across short and long stay | 3,346.86 | 1 | 3,346.86 | (18) |
| Weighted average ICU cost per patient | 1,705.06 | 0.04 | 68.2 | (18) |
| Transfer from emergency department | 673.65 | 0.634 | 427.09 | (6, 18) |
| Referral from sexual health services | 407.29 | 0.366 | 149.07 | (17) |
| **Total cost per hospitalisation with 4% ICU (baseline and upper bound)** | | | **3,991.22** | |
| **Alternative cost for GUM infection from PSSRU (lower bound)** | | | **3,776** | (19) |

**Noted:** Weighted averages are calculated based on number of activities attributed to each HRG code. We assume that patients who are in intensive care also have time hospitalised on the ward, so the ICU cost is in addition to the hospital stay.

**Abbreviations:** ICU, intensive care unit; GUM, genitourinary medicine.

##### Vaccination Administration Costs

Vaccination administration costs are presented in Table S9. The cost of administering the first and second doses of the mpox vaccine was obtained from the Sexual Health Services tariff for mpox (17). Since data on the cost per vial of the mpox vaccine was not available due to confidentially, in the base case we use the costs for the Shingrix vaccine as provided by the BNF Tariff (16), and we also conduct a threshold analysis (see Supplementary III—Sensitivity analysis) to identify the threshold cost for the mpox vaccine at which each vaccination scenario would be cost-effective at £20,000/QALY or 30,000/QALY. We do not account for uncertainty in the administration costs.

**Table S9:** Vaccination Administration Costs

| Parameter | Unit cost (£, 2022) | Ref |
|---|---|---|
| Administration for first dose MPX vaccine<br>(Including: staff time costs for registration, consultation, vaccine administration, and health promotion, as well as consumables such as gloves, plasters, and cotton) | 36.85 | (17) |
| Administration for second dose MPX vaccine<br>(Reduced consultation time) | 25.78 | (17) |
| **Vaccine cost**<br>Assume cost of Shingrix | 160 | (16) |

##### Public Health Response Costs

To estimate the costs of the public health response to the outbreak, we gathered staffing and other costs from UKHSA attributed to the outbreak. From the national response, costs were accounted for on the emergency planning response and resilience (EPRR) incident budget code across the whole outbreak period. In addition, we gathered response costs from three regions (London, and two anonymous regions 2 and 3), where the majority of mpox cases occurred in 2022. Each of the three regions reported strategic costs and per case costs. We assumed that response costs in the remaining regions of England were similar to the non-London regional data we had.

Potential explanations for cost differences by region include reporting variations, clinical variations, and case allocation differences. Reporting variations were due to the availability of local information, data collected retrospectively, and requests being made amidst various competing year-end pressures. Clinical variations included differences in the contribution of external stakeholders to case management and patient pathways, as well as differences in their expectations of UKHSA senior support and engagement. Case allocation was determined using postcode allocation, available for 60% of Second Generation Surveillance System (SGSS) samples; the others were allocated by the GP postcode. The estimations of total costs per week and total costs per case are listed in Table S10 and S11, respectively.

**Table S10:** Fixed Outbreak Strategic Costs (17 weeks)

| Category | Description | Total cost for 17 weeks (£, 2022) | Cost per week (£, 2022) |
|---|---|---|---|
| Central UKHSA | Includes agency pay, lab consumables, lab equipment, other payroll costs, conferences, travel and subsistence, this excludes the cost of those diverted from business-as-usual (BAU) work. | £819,999 | £48,235 |

| | | | |
|---|---|---|---|
| London strategic costs | Includes meeting attendance (national and local), protocol development, staff training, developing protocols, liaison with external stakeholders. | £72,600 | £4,271 |
| Strategic costs for region 2 | Estimated staff costs | £77,265 | £4,545 |
| Strategic costs for region 3 | Cost for consultant | £45,067 | £2,651 |
| The remaining 6 regions | Assumed average total of region 2 & 3 (two rows above) and multiply by 6 | £366,996 | £21,588 |
| **Total per week** | | | **£81,290** |
| **Sensitivity analysis** | **Assumed region 2 (higher) or 3 (lower) staff costs apply to all non-London regions** | | **£73,714 - £88,866** |

**Table S11:** Per Case Costs

| Region | Description | Cost per case (£, 2022) |
|---|---|---|
| London | Includes staff costs from response cell rota including admin staff, health protection practitioners, and consultants. | £184 |
| Region 2 | Includes staff costs from response cell rota including admin staff, health protection practitioners, and consultants. | £1,308 |
| Region 3 | Costs for staffing the mpox cell. | £401 |
| 8 non-London regions | Weighted the average costs between region 2 and 3 based on the total number of reported cases in each region (212 cases in region 2 and 297 cases in region 3), to get an average per case cost for all 8 non-London regions | £779 |
| **Weighted average cost per case** | **61% of cases were in London. We weighted between London and all other regions. Calculated as £184 * 0.61 + £779 * 0.39.** | **£416** |
| **Sensitivity analysis** | **For lower bound, we assumed London cost for all based on best economy of scale on response and assumed regional average costs for all to estimate the upper bound** | **£184 - £779** |

**Productivity Loss Costs**

To estimate the costs of productivity loss due to mpox infection, we used data from the RiiSH -MPOX survey from December 2022 (8), which provided insights into the patterns of absenteeism and productivity loss due to self-reported mpox infections. The survey assumed that:

- 75% (n=3/4) of severe inpatient cases took an average of 20 days off work.
- 68.6% (n=24/35) of moderate cases took an average of 14.8 days off work.
- 67.7% (n=21/31) of mild cases took an average of 13.1 days off work.

Additionally, the RiiSH data suggests that while people continued to work during their illness, mpox cases experienced a 45.7% reduction in productivity over an average of 10.8 days (8). We based our calculations on an employment rate of 75.6%, using data from the Office for National Statistics (ONS) for 2022 for individuals aged 16-64, and an average weekly wage of £683 (20, 21). This estimated the productivity losses per case as follows:

- For non-hospitalised cases (i.e., mild cases), the productivity loss cost is approximately £1,535.
- For outpatient cases (i.e., Moderate cases), the productivity loss cost is approximately £1,709.
- For inpatient cases (i.e., Severe cases), the productivity loss cost is approximately £2,376.

#### Health-Related Quality of Life

No suitable QALY weights (estimated using EQ-5D surveys) are available for mpox. This is an area of uncertainty in evaluating the cost-effectiveness of interventions related to mpox. We obtained baseline health-related quality of life (HRQoL) data from an EQ5D questionnaire for GBMSM who had never tested positive for mpox, in the RiiSH-MPOX survey undertaken in December 2022 (8). We then applied the disability weights from the Global Burden Diseases (GBD) 2021 study to calculate utility scores for different severity levels of mpox (28). The disability weights were derived from infectious diseases at mild (0.006 (0.002 – 0.012)), moderate (0.051 (0.032 – 0.074)), and severe levels (0.133 (0.088 – 0.190)) (28) . The disability weights were subtracted from the baseline HRQoL data for uninfected GBMSM (8) to estimate the impact over a year of infection with mpox at each level (Table S12).

In addition, we reviewed the literature for health-related quality of life measures in patients with herpes zoster, which we assumed would lead to similar pain levels as mpox. We found 8 studies, with the additional limitation that all studies included only patients at least aged 50 or older, which is different from the demographic of patients with mpox. One study was conducted in the UK, and this also broke down HRQoL scores based on whether the patient was experiencing mild (0.7), moderate (0.66), or severe pain (0.52), based on EQ-5D questionnaires (29) . Assuming a duration of infection of 21 days for all cases, we calculated the annual HRQoL average for patients with mpox assuming for the remainder of the year they had the baseline utility weight of 0.839.

The uncertainty bounds around the GBD HRQoL weights were then combined with the values from herpes zoster, resulting in wider uncertainty bounds for moderate and severe mpox. Details on utility scores are presented in Table S12.

**Table S12:** Utility scores calculated by adapting disability weights

| **Health state** | **Disability weight (per year in which infection occurred)** | **HRQoL from herpes zoster, for 21 days out of the year** | **Final HRQoL weights for year of infection with mpox** | **Ref** |
|---|---|---|---|---|
| Baseline utility | 0.839 | | $UW_0$=0.839 | RiiSH-MPOX survey (8) |
| Mild MPOX | 0.833 (0.827 – 0.837) | 0.831 | $UW_A$= 0.833 (0.827 – 0.837) | (28, 29) |
| Moderate MPOX | 0.788 (0.765 – 0.807) | 0.829 | $UW_M$=0.788 (0.765 – 0.829) | (28, 29) |
| Severe MPOX | 0.706 (0.649 – 0.751) | 0.821 | $UW_H$=0.706 (0.649 – 0.821) | (28, 29) |

**Abbreviations:** HRQoL, health-related quality of life.

#### Supplementary III: Sensitivity analyses

As in the baseline analysis, the following parameter value and ranges are assumed: Durations of protection induced for 1 dose and 2 doses of vaccine are set to $L_{imm1}$=5 years and $L_{imm2}$= 10 years respectively; effectiveness of 1 dose and 2 doses of vaccine are $V_{E,1}$=78% (54 – 89%) and $V_{E,2}$= 89% (78 – 100%)(5, 30). Immunity induced by natural infection is assumed to be 100% against infection during the immunity duration of $L_{imm0}$= 10 years. The contact matrix is based on data on anal sex contact from the 2021 RiiSH survey(4). The seeding rate of imported infections during the next 20 years is assumed to be 6 cases per month(12) and the annual discount for QALY and costs is $q$ = 3.5%(15). Vaccine price is set at 160.00 pound per dose based on the cost of the Shingrix vaccine (16). The total costs summarise the costs due to clinical case management, vaccination, and public health responses. The results for impact and cost-effectiveness of the vaccination scenarios under the baseline analysis are given in Table 3 in the main text with Table S13 giving more details of the cost breakdown.

Because of numerous parameter uncertainties and assumptions, we undertook 13 sensitivity analyses to see how they affected our cost-effective projections for the different modelled reactive and pre-emptive vaccine scenarios. In the following, the terms "increase" and "decrease" are referred to the comparisons of the sensitivity analysis scenarios with the equivalent vaccination scenarios from the baseline analysis.

##### 1. Lower annual discount rate (1.5%) on costs and utilities

Lowering the discount rate to 1.5% (from 3.5%) increases the incremental QALYs and costs saved for any vaccination scenario by 26–32% (compared to the same vaccination scenarios in the baseline), but doesn't change which is the optimal vaccination scenario (pre-emptive vaccination at 41 per day; Table S14). All scenarios are cost-saving at the baseline vaccine cost, with vaccination remaining cost-effective unless the vaccine is very expensive (>£658 per dose) (Table S15).

##### 2. Remove public health response costs from the baseline

Excluding public health response costs results in vaccination saving less costs for all vaccination scenarios copared to the baseline scenario (Table S16). Vaccination is only cost saving when daily number of vaccine doses is ≤27 for pre-emptive vaccination, or ≤41/day for reactive vaccination. The optimal scenario is now reactive vaccination at 27 doses per day for the baseline vaccine cost. Vaccination remains cost-effective (although at a lower rate) if the vaccine price is less than £356 per dose (Table S15).

##### 3. Include productivity losses due to mpox disease

Including productivity losses in the total costs makes each vaccination scenario more cost-saving by 140–170% (compared to the same vaccination scenarios in the baseline) but doesn't change the QALYs saved or which vaccination scenario is optimal (Table S17). Vaccination remains cost-effective (although at a lower vaccination rate) unless vaccine price is very expensive (> £1351 per dose; Table S15).

#### 4. Long duration of protection

Increasing the duration of vaccine protection (Limm1= 10 years and Limm2=Limm0=20 years) reduces the number of outbreak days and the outbreak size, which consequently decreases the total costs and increases the QALYs with no vaccination, but decreases the incremental QALYs by 47-47% and cost savings by 21-40% when comparing scenarios to no vaccination (compared to the same vaccination scenarios in the baseline; Table S18). All the modelled scenarios remain cost-saving compared to no vaccination, but the optimal scenario changes to pre-emptive vaccination at 27 doses per day. Vaccination remains cost-effective (although at a lower rate) if the vaccine price is less than £514 per dose (Table S15).

#### 5. Short duration of protection

Decreasing the duration of vaccine protection (Limm1=2.5 years and Limm2=Limm0=5.0 years) increases the number of outbreak days and the outbreak size (except PV81 and PV135), which consequently increases the total costs and decreases the QALYs with no vaccination, but increases the incremental QALYs and decreases the cost savings when comparing vaccination scenarios to no vaccination (compared to the same vaccination scenarios in the baseline; Table S19). All the modelled scenarios remain cost-saving compared to no vaccination, but the optimal scenario changed to pre-emptive vaccination at 54 doses per day. Vaccination remains cost-effective (although at a lower rate) if the vaccine price is less than £621 per dose (Table S15).

#### 6. Low importation rate

Under the low importation rate (seeding rate=1 case per month), both the number of outbreak days and the outbreak size become smaller with the QALYs saved from vaccination slightly increasing and the incremental costs decreasing for pre-emptive vaccination but remaining very similar for reactive (compared to the same vaccination scenarios in the baseline; Table S20). All the modelled programs remain cost-saving as in baseline scenarios; but the most cost-effective scenario changes to pre-emptive vaccination at 27 doses per day. Vaccination remains cost-effective (although at a lower rate for reactive) if the vaccine price is less than £734 per dose (Table S15).

#### 7. High importation rate

Without vaccination, the high importation rate (seeding rate=10 cases per month) increases the number of outbreak days but decreases the outbreak size compared to the baseline model (Table S21). This is due to the interaction between importation and responsive behaviour: under the high importation rate of 10 cases per month, an outbreak can be maintained for a long period under the outbreak criterion (starts with 120 cases and ends with less than 60 cases within three months) (Figure S5). With vaccination, the QALYs gained reduces by 32-39% and less costs are saved by 37-59% (compared to the same vaccination scenarios in the baseline; Table S21), but all scenarios remain cost-saving compared to no vaccination. The optimal scenario changes to pre-emptive vaccination at 135 doses per day. Vaccination remains cost-effective (although at a lower rate for reactive) if the vaccine price is less than £506 per dose (Table S15).

#### 8. Low outbreak criteria

With no vaccination, lowering the outbreak criteria by 20% (an outbreak starts with >=96 cases and ends with ≤48 cases in 3 months) increases the number of outbreak days but decreases the outbreak size, which increases slightly the QALYs but decreases the total costs. With pre-emptive vaccination (except for a very low vaccination rate such as PV13), lowering the outbreak criteria increases both the outbreak size and duration (compared to baseline), while for reactive vaccination it increases the outbreak duration but increases outbreak size only for high vaccination rates (>135 doses per day). For reactive and pre-emptive vaccination, the incremental QALYs and cost savings from vaccination decrease by 21-32% (compared to the same vaccination scenarios in the baseline; Table S22) when the outbreak criteria is lowered but the optimal vaccination scenario remains pre-emptive vaccination at 41 doses per day. Vaccination remains cost-effective (although at a lower rate) if the vaccine price is less than £564 per dose (Table S15).

##### 9. High outbreak criteria

With no vaccination, increasing the outbreak criteria by 20% (an outbreak starts with >=144 cases and ends with ≤72 cases in 3 months) decreases outbreak days but increases the outbreak size, which decreases slightly the QALYs but increases the total costs. With pre-emptive vaccination (except for that of very low vaccination rate such as PV13), increasing the outbreak criteria decreases both outbreak size and duration (compared to baseline), while for reactive vaccination it decreases the outbreak duration but increases the outbreak size when vaccination <465 dose per day. For reactive and pre-emptive vaccination, the incremental QALYs and cost savings from vaccination increase by 9-16% when the outbreak criteria is increased (compared to the same vaccination scenarios in the baseline), but the optimal vaccination scenario remains pre-emptive vaccination at 41 doses per day (Table S23). Vaccination remains cost-effective (although at a lower rate) if the vaccine price is less than £715 per dose (Table S15).

##### 10. No responsive behaviour changes during outbreaks

Without responsive behaviour change during each outbreak, both the duration and size of the outbreak increases and mpox becomes endemic (Figure S6) with no vaccination, and consequently QALYs decrease and the total costs increase (Table S24). However, vaccination can save much more costs due to CCM and PHR to balance the costs due to vaccination and all the modelled scenarios are cost-saving with the most cost-effective scenario being pre-emptive vaccination at a higher vaccination rate of 271 doses per day. Vaccination remains cost-effective (although reactive and at a lower rate) if the vaccine price is up to £2131 per dose (Table S15).

##### 11. Using alternative data to define the contact matrix

If we use physical contact data (not anal sex contact data) from the RiiSH MPOX survey in December 2022 to parameterise our model instead of anal sex contact data from the 2021 RiiSH survey then we have a larger proportion of high-risk SHS GBMSM (23.3% versus 11.9% for the baseline scenario). For this sensitivity analysis, the model needs to be re-calibrated to case data from the 2022 outbreak. If we use the same methods to do this as for the baseline model, then the new model projects that mpox incidence will increase once sexual behaviours revert to the levels before the 2022 outbreak (Figure S7). This is not what we observed during 2023. To avoid these projections, this version of the model is calibrated to observational data up to 10 January 2023. Under the new estimates of model parameters (Table S25), a potential outbreak in 2023 is avoided (Figure S7).

To conduct the sensitivity analysis using this new contact matrix, we use estimates of the transmission model calibrated to 10 January 2023 as listed in Table S25. The cost-effectiveness of vaccination strategies based on these parametrisations is shown in Table S26. With no vaccination, the model incorporating this new contact matrix projects more outbreak days but smaller outbreak size (compared to baseline), which consequently increases the QALYs and decreases the total costs compared to the baseline model.

With pre-emptive vaccination, both the incremental QALYs and costs (compared to baseline projections) always decreases; however, the incremental QALYs always increase with vaccination rate while the incremental costs first decrease then increase when vaccination rate ≥54 doses per day. This is due to the larger proportion of high-risk GBMSM (23.3% versus 11.9% for the baseline scenario) so more vaccines can be delivered before saturation occurs. This makes pre-emptive vaccination at rates above 54 doses per day and below 27 doses per day not cost saving.

With reactive vaccination, both outbreak size and duration increase (compared to baseline projections) when vaccination rate ranges from 41 to 203 doses per day, while the incremental QALYs and costs decrease compared to baseline projections. Incremental QALYs always increase with vaccination rate and costs saved decrease when vaccination rate is smaller than 465 doses per day. Most of the modelled scenarios of reactive vaccination are still cost-saving compared to the no vaccination counterfactual, except at low vaccination rates of <41 per day. Irrespective, pre-emptive vaccination at 41 doses per day is still the most cost-effective scenario as in the baseline scenario. Vaccination remains cost-effective (at the same rate) if the vaccine price is up to £209 per dose (Table S15).

12. **Breakthrough infections have less severe symptoms**

Empirical observations indicate that clinical features and outcomes of re-infection and breakthrough infections after vaccination appear to be less clinically severe than those described in 2022 case literature (31). To test whether this affects our cost-effectiveness estimates, this sensitivity analysis assumes breakthrough infections have half the chance of experiencing moderate and severe mpox disease compared to the baseline scenario, that is, $p_1$=3.05% and $p_2$=4.15%. The results in Table S27 show that without vaccination, QALYs increase by 3 and total costs decrease by £62,000 and the changes in both incremental QALYs and costs are small (compared to projections for the baseline model). The most cost-effective vaccination scenario remains pre-emptive vaccination at 41 doses per day. Vaccination remains cost-effective (at the same rate) if the vaccine price is up to £650 per dose (Table S15).

13. **Fractional vaccination doses**

During the 2022 outbreak, fractional were delivered in some settings because of limitations in vaccine supply, with evidence suggesting that this does not substantially reduce the effectiveness of the vaccine(32). Going forward, busier SHS clinics may use fractional doses to reduce vaccine costs. We undertook a sensitivity analysis to determine whether the use of fractional doses for half of all vaccinations would affect our cost-effectiveness estimates. To be conservative, we assumed that fractional vaccination would reduce the effectiveness of the vaccine by 25% in relative terms following 1 or 2 doses. In comparison to the baseline scenario, the results in Table S28 show that using fractional doses results in more costs being saved from vaccinating, but fewer QALY being gained because of the hypothesised lower vaccine effectiveness. This means that higher vaccination rates are preferred with the optimal vaccination scenario being pre-emptive vaccination at 271 doses per day.

### Supplementary IV: Figures and Tables

#### Figures S3-S8

**Figure S3** Epidemic projections of the baseline model for different vaccination scenarios over 20 years from 2024 to 2043. Four different rates of two vaccination schemes are shown. Pre-emptive vaccination (PV) at rates of 0, 27, 41 and 81 doses per day and reactive (RV) of 27, 81, 135, and 203 doses per day. The figures also show the number of vaccines given out as first and second doses and evolution of susceptibility in the model. At low rates of pre-emptive vaccination (<41 doses per day), 1st dose was never getting saturated and the allocation for 1st and 2nd dose followed the assumed ratio of 1.00:0.56. When vaccination rate reach or exceeds 41 doses per day, 1st dose got saturated within about one year, then it is followed by a transient period during which more vaccines are given out as 2nd dose; once 2nd dose is also saturated, the vaccine doses are given out evenly as 1st and 2nd doses.

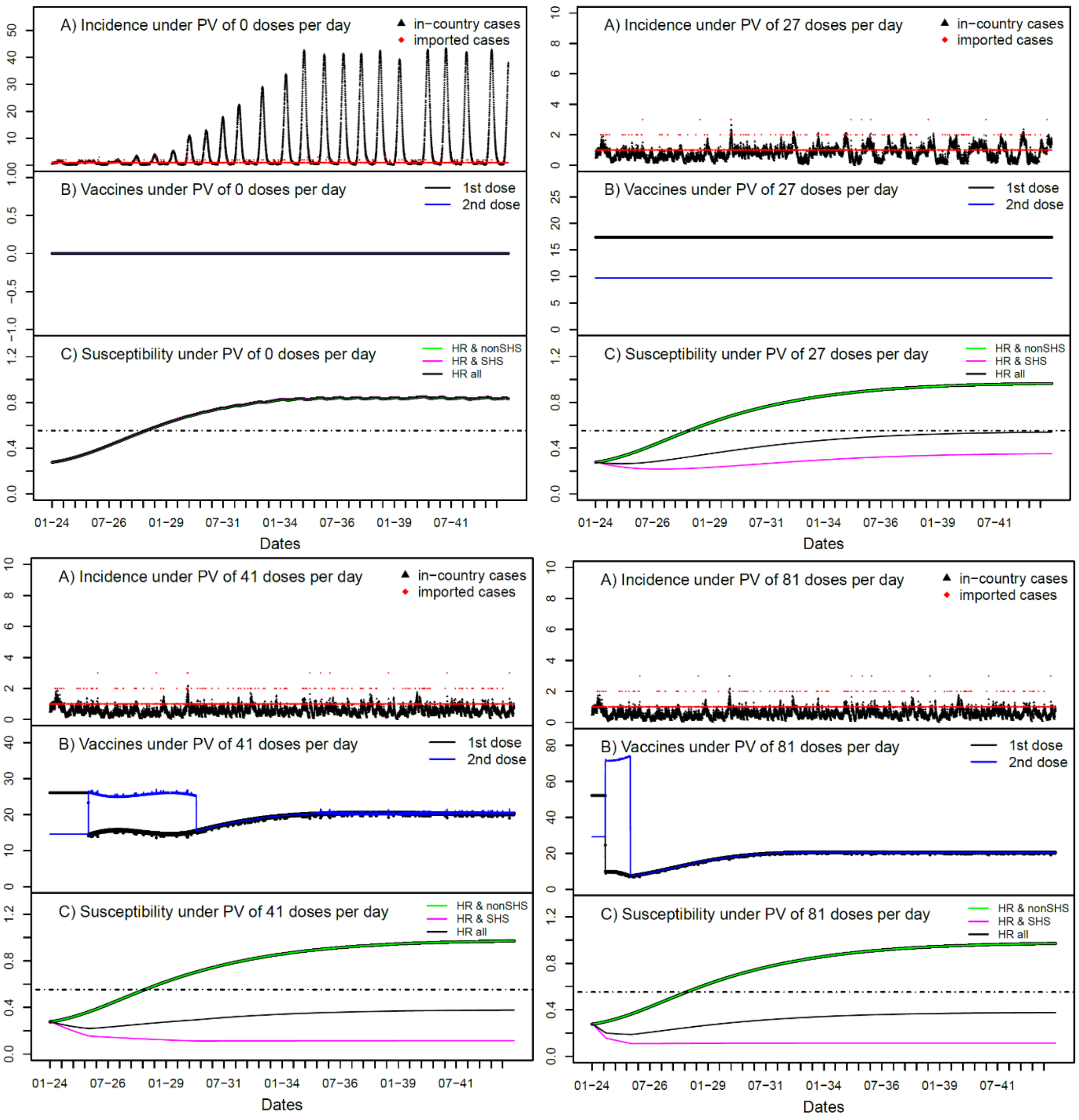

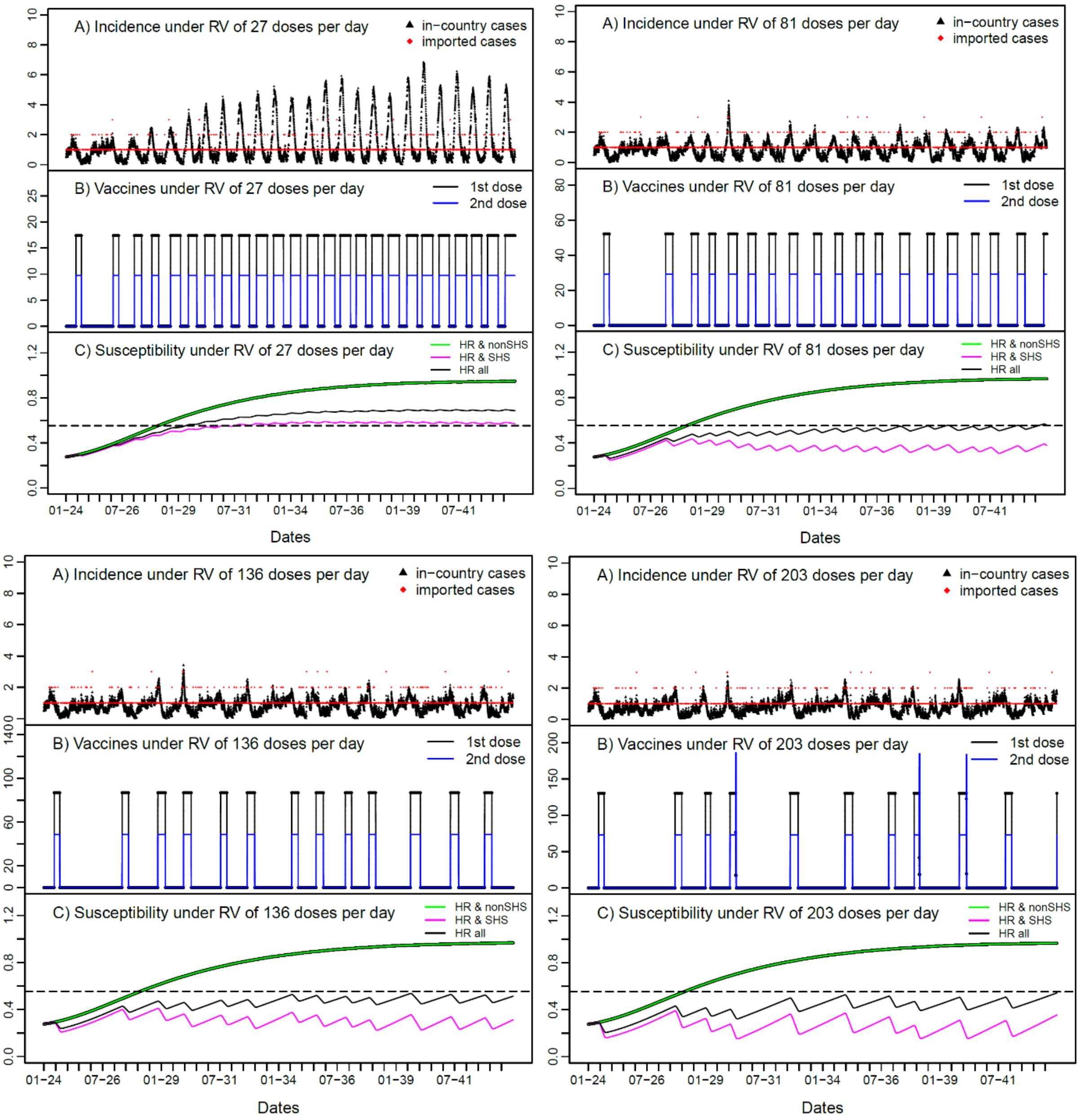
A) Incidence under RV of 27 doses per day
in−country cases
imported cases
B) Vaccines under RV of 27 doses per day
1st dose
2nd dose
C) Susceptibility under RV of 27 doses per day
HR & nonSHS
HR & SHS
HR all
A) Incidence under RV of 81 doses per day
B) Vaccines under RV of 81 doses per day
C) Susceptibility under RV of 81 doses per day
A) Incidence under RV of 136 doses per day
B) Vaccines under RV of 136 doses per day
C) Susceptibility under RV of 136 doses per day
A) Incidence under RV of 203 doses per day
B) Vaccines under RV of 203 doses per day
C) Susceptibility under RV of 203 doses per day
01−24
07−26
01−29
07−31
01−34
07−36
01−39
07−41
Dates

**Figure S4** Annual number of vaccines given during the 20-year period from 2024 under baseline scenario: A) the annual number of total vaccines given out under six rates of pre-emptive vaccination (PV) (13, 27, 40, 54, 81 and 135 doses per day); B) the annual number of vaccines given out under six rates of reactive vaccination (RV) (27, 41, 81, 135, 204 and 465 doses per day).

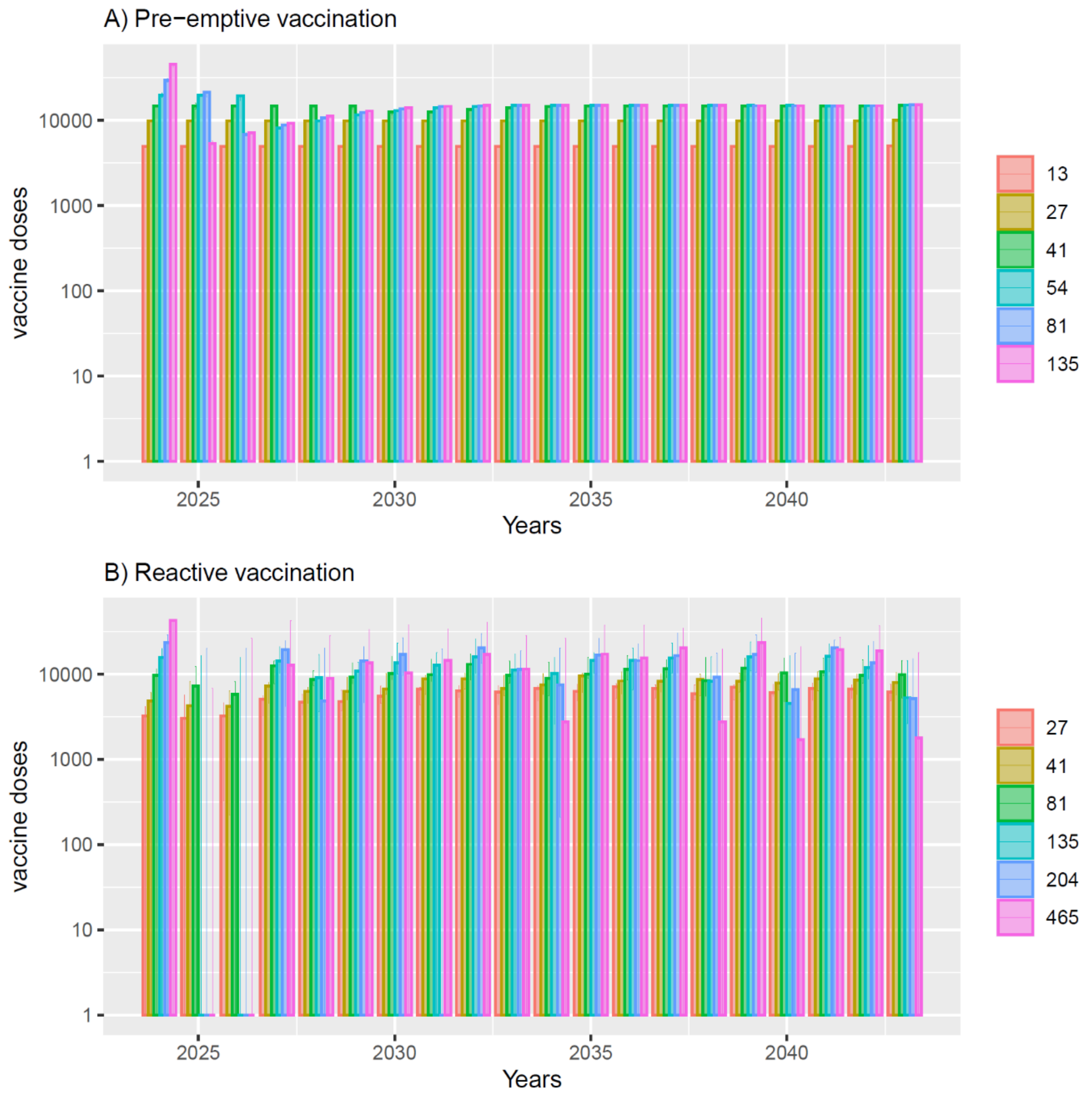

**Figure S5** Impact of importation rate on the epidemic outbreaks with responsive behaviour change. No vaccination was assumed. Importation rate: A) one case per month, B) six cases per month (baseline), and C) ten cases per month. The red dots represent imported cases. The thick black line represents the median model projections, and the grey shading is their 95% CrI.

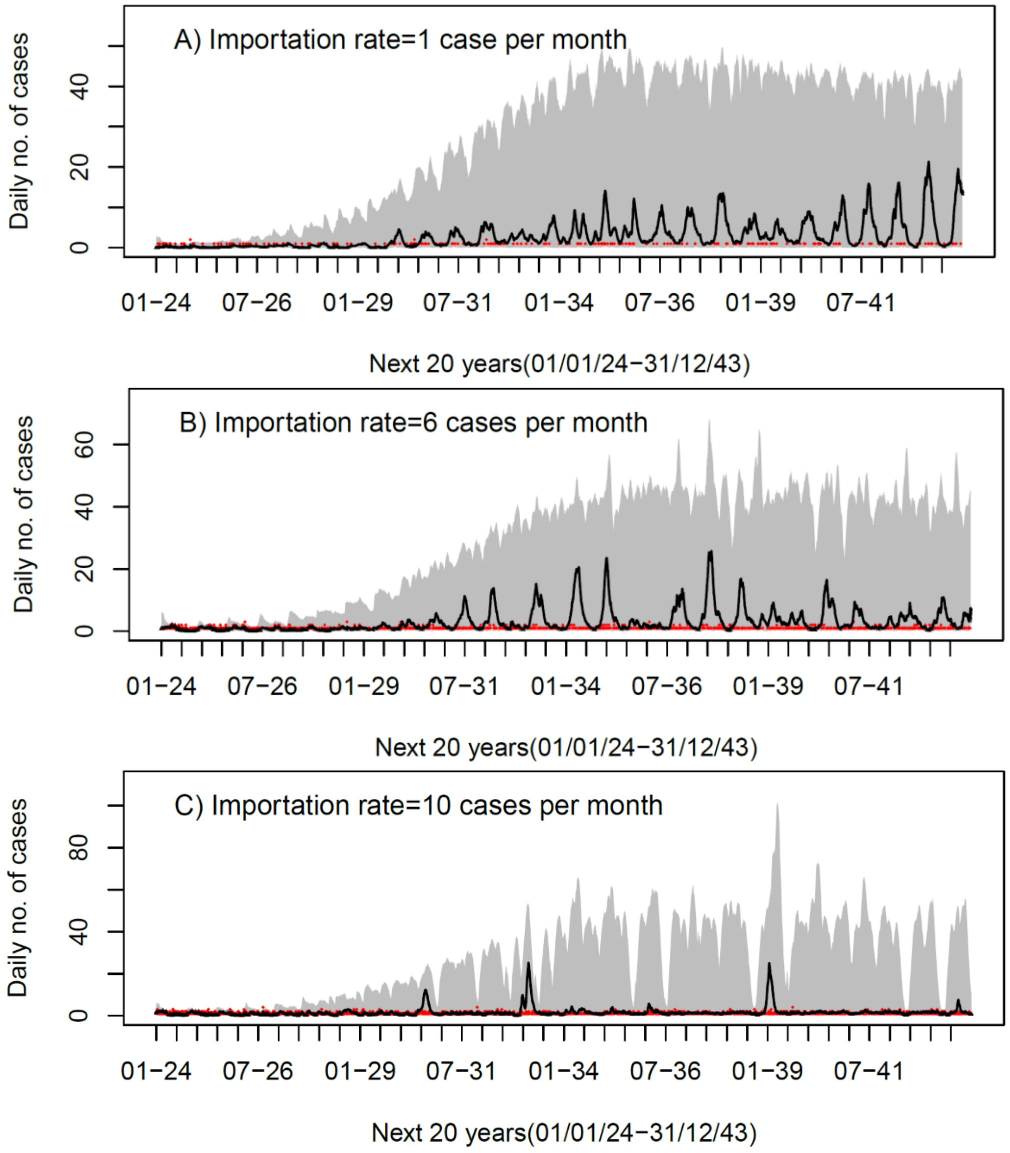

**Figure S6** Impact of having no responsive behaviour change on the epidemic outbreaks (compared to baseline of having responsive behaviour change). Three scenarios were considered: No vaccination (first row), pre-emptive vaccination at 41 doses per day (2nd row) and reactive vaccination at 203 doses per day (3rd row) A) with B) without responsive behaviour change. The red dots represent imported cases. The thick black line represents the median model projections, and the grey shading is their 95% CrI.

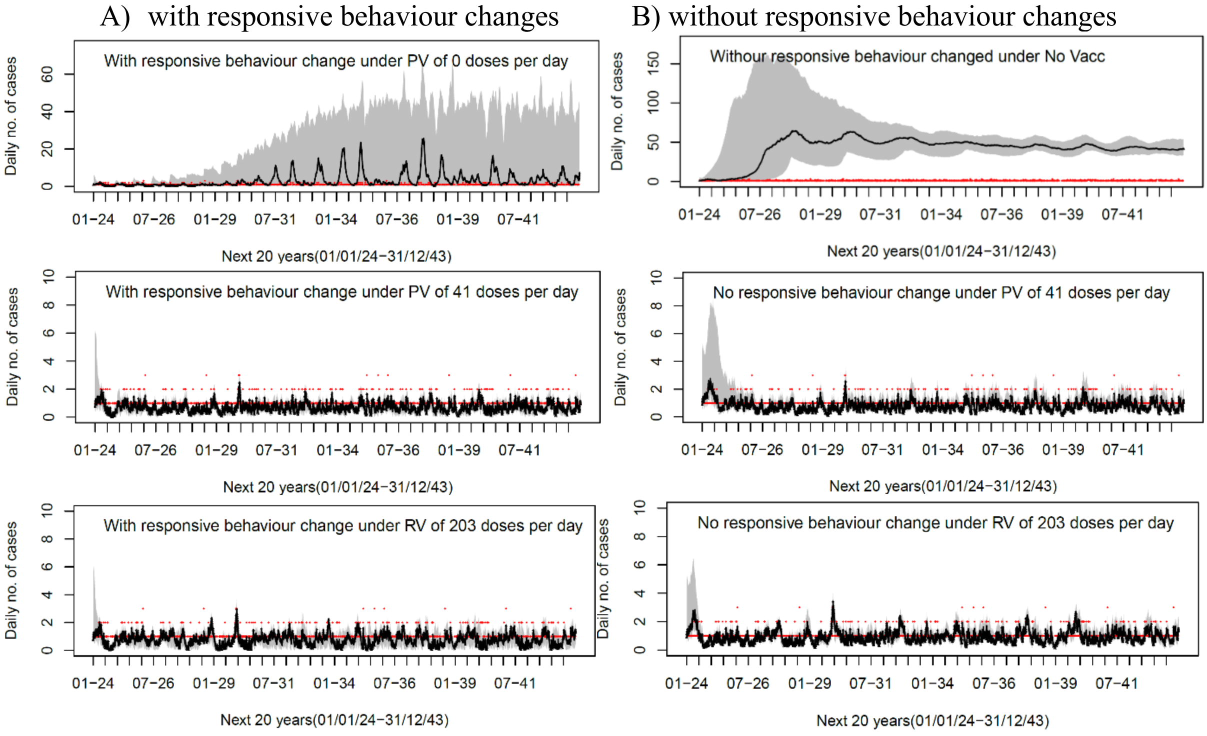

**Figure S7** Model predictions up to December 2023 for the transmission model using contact matrix constructed using the 2022 RiiSH-MPOX physical contact data(8). The predictions in A), based on the model calibration up to 12 August 2022, show that there would be a large outbreak in 2023; the predictions in B), based on the model calibration up to 10 January 2023, show that an outbreak in 2023 is avoided. The blue triangles and red diamonds represent in-country and imported cases, respectively. The thick black line represents the median model projections and grey shading is their 95% CrI. The four vertical lines represent the middle turning point of contact rate (5 and 21 June 2022; red), start of vaccination (27 June; black), date up to which observational data used for model calibration (12 August 2022 and 10 January 2023; purple) and date up to which data used for model validation and from when the contact behaviour reverts to normal (16 November; pink).

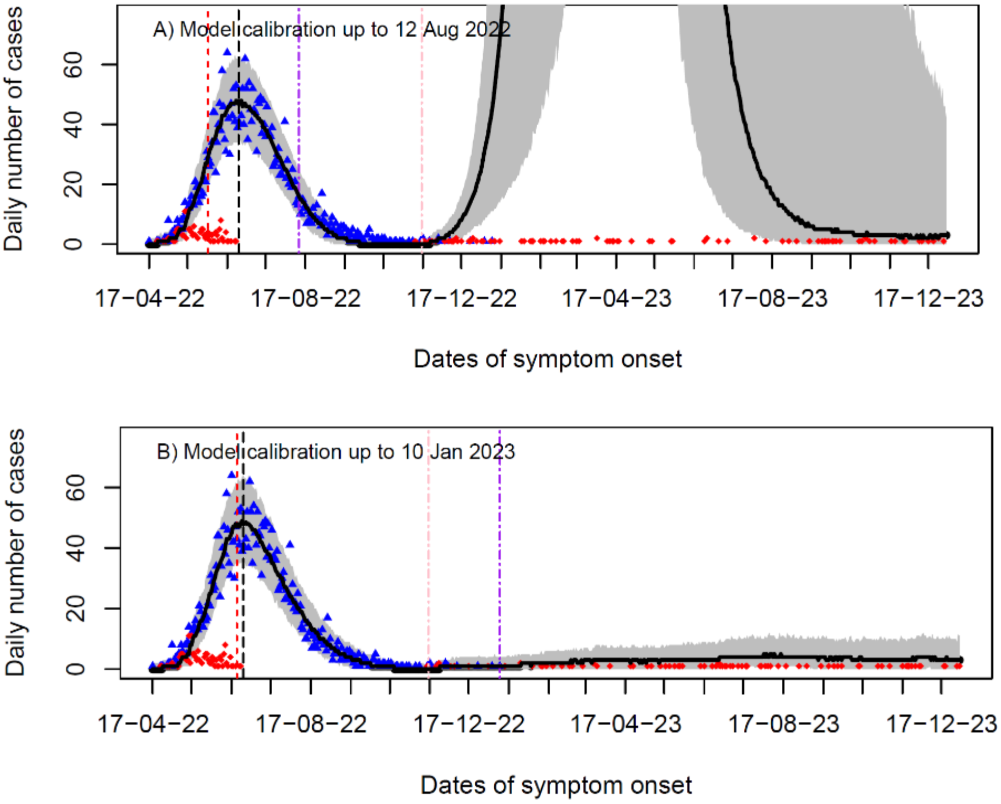

**Figure S8** Threshold vaccine prices for baseline scenario. A) calculation of the threshold price at which the median ICER is below £20k/QALY (V50), or the threshold price at which the 90% of ICER is below £30k/QALY (V90) under pre-emptive vaccination at 41 doses per day; B) Comparison of threshold vaccine price under different vaccination scenarios and vaccination rates. The threshold vaccine price is lower for pre-emptive vaccination scenarios and higher vaccination rates.

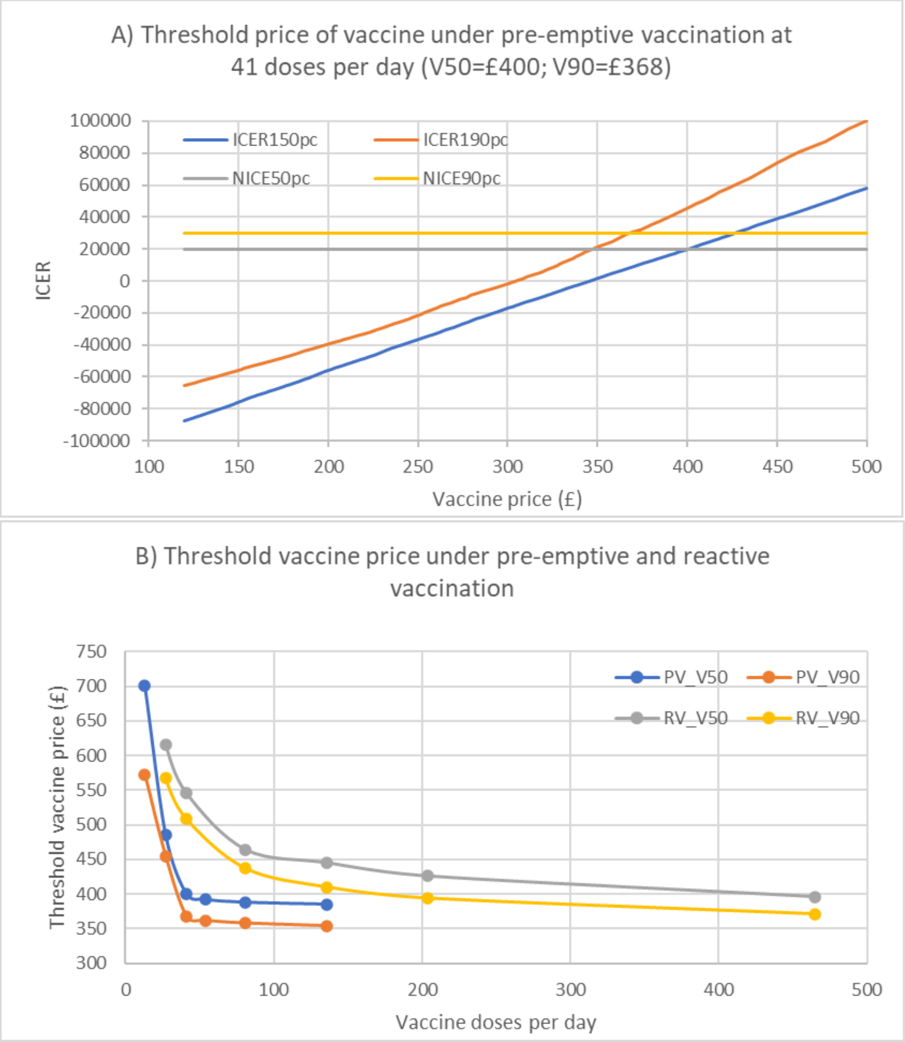

#### TABLES S13-S28 – SENSITIVITY ANALYSES

Table S13 Breakdown of total costs in the baseline scenario. The mean and 95%CrI are listed. The units for all costs are 1000s £.

| scenario | Outbreak size, number of case | Outbreak duration (days) | Vaccination costs | CCM cost | PHR cost | PL costs | total costs (with PHR but not PL) | total costs exclusive of PHR | total costs inclusive of PL |
|---|---|---|---|---|---|---|---|---|---|
| No Vacc | 54891 (26246,67256) | 5472 (5141,5676) | 0 (0,0) | 25043 (13228, 31583) | 61085 (50759,73068) | 61206 (32116, 74885) | 86127 (67671, 102039) | 25043 (13228, 31583) | 147333 (101595, 176598) |
| Pre-emptive vaccination of GBMSM at high risk for mpox who attend SHS | | | | | | | | | |
| PV 13 vaccines per day | 1309 (8224,18452) | 4219 (3494,4673) | 14041 (14041,14041) | 7275 (5202, 9636) | 37867 (28834, 44692) | 17780 (13104, 23052) | 59184 (48271, 68408) | 21316 (19243, 23677) | 76964 (61821, 90753) |
| PV 27/day | 2143 (637,3860) | 2210 (802,3198) | 28082 (28082,28082) | 2978 (2534, 3599) | 18260 (6003, 27833) | 7280 (6359, 8587) | 49320 (36731, 59342) | 31060 (30616, 31681) | 56600 (43129, 68195) |
| PV 41 vaccines per day | 254 (0,1143) | 369 (0,1719) | 40698 (40511,40735) | 2412 (1760, 2955) | 3417 (0, 14870) | 5896 (4357, 6967) | 46527 (42497, 58089) | 43110 (42494, 43627) | 52423 (46855, 64735) |
| PV 54 vaccines per day | 234 (0,1101) | 340 (0,1641) | 41576 (41272,41637] | 2372 (1727, 2967) | 3104 (0, 14233) | 5798 (4269, 6980) | 47052 (43363, 58261) | 43948 (43356, 44489) | 52850 (47636, 64662) |
| PV 81 vaccines per day | 221 (0,1064) | 325 (0,1594) | 42077 (41728,42146) | 2348 (1705, 2936) | 2920 (0, 14182) | 5740 (4202, 6945) | 47345 (43851, 58526) | 44425 (43839, 44954) | 53085 (48055, 65012) |
| PV 135 vaccines per day | 203 (0,1022) | 305 (0,1523) | 42474 (42078,42553) | 2337 (1693, 2933) | 2675 (0, 13239) | 5713 (4163, 6909) | 47487 (44244, 58324) | 44812 (44227, 45336) | 53201 (48396, 65003) |
| Reactive vaccination of GBMSM at high risk for mpox who attend SHS | | | | | | | | | |
| RV 27 vaccines per day | 9841 (7021,13064) | 4164 (3745,4450) | 15407 (13510,16667) | 6035 (4858, 7563) | 37273 (31761, 42042) | 14749 (12162, 18133) | 58714 (50229, 65689) | 21442 (18514, 24141) | 73463 (62597, 83515) |
| RV 41 vaccines per day | 5597 (4074,7416) | 3596 (3171,3909) | 20059 (17290,22078) | 4274 (3639, 5237) | 31486 (26446, 35797) | 10446 (9100, 12389) | 55820 (47818, 62590) | 24333 (21146, 27267) | 66266 (56994, 74805) |
| RV 81 vaccines per day | 2157 (1747,2722) | 2487 (2060,2842) | 28076 (22677,32446) | 2903 (2635, 3205) | 21509 (17318, 25098) | 7096 (6811, 7580) | 52487 (43539, 60384) | 30979 (25823, 35351) | 59584 (50623, 67844) |
| RV135 vaccines per day | 1284 (1071,1605) | 1699 (1370,2233) | 32012 (25221,39010) | 2773 (2456, 3095) | 14776 (11597, 19168) | 6781 (6254, 7355) | 49561 (40180, 60425) | 34785 (28352, 41729) | 56342 (47477, 66815) |
| RV 204 vaccines per day | 960 (729,1565) | 1328 (966,2148) | 34651 (27125,40265) | 2707 (2448, 2993) | 11555 (8230, 18061) | 6618 (6262, 7127) | 48913 (38452, 61210) | 37358 (30062, 42898) | 55531 (45324, 67622) |
| RV 465 vaccines per day | 811 (462,1530) | 1130 (630,2111) | 37669 (33635,41068) | 2643 (2340, 2919) | 9684 (5274, 17782) | 6462 (5803, 6857) | 49996 (41530, 61447) | 40311 (36155, 43787) | 56457 (47905, 67990) |

Abbreviations: CCM, Clinical case management; PHR, Public health response; PL, Productivity losses.; PV, pre-emptive vaccination; RV, reactive vaccination

Table S14 Cost effectiveness of vaccination program under annual discount rate of 1.5%. The values listed are mean and 95% CrI.

| Scenario | Total number of infections | Outbreak duration (days) | Total Cost (1000s £) | Total QALYs | Incremental costs compared to no vaccination (1000s £) | Incremental QALY compared to no vaccination | ICER compared to no vaccination | Fully incremental analysis | INMB compared to No Vaccination at £20,000/QALY | INMB compared to no vaccination at £30,000/QALY |
|---|---|---|---|---|---|---|---|---|---|---|
| No Vaccination counterfactual | 54891 [26246,67256] | 5472 [5141,5676] | 106803 (83594, 126730) | 11248670 (11248346, 11249036) | – | – | – | Dominated | – | – |
| Pre-emptive vaccination (PV) of GBMSM at high risk for mpox who attend SHS | | | | | | | | | | |
| PV 13 vaccines per day | 13099 [8224,18452] | 4219 [3494,4673] | 72363 (59468, 83382) | 11249213 (11249101, 11249311) | -34440 (-48029,-18309) | 542.35 (220.82, 777.87) | Cost-saving | Dominated | 45286918 (21680433, 61355269) | 50710413 (23513116, 68385200) |
| PV 27 vaccines per day | 2143 [637,3860] | 2210 [802,3198] | 59367 (44300, 71041) | 11249345 (11249308, 11249378) | -47436 (-63893,-33523) | 674.97 (322.11,971.53) | Cost-saving | Dominated | 60935785 (41690837, 80471553) | 67685500 (44281760, 88688560) |
| **PV 41 vaccines per day** | **254 [0,1143]** | **369 [0,1719]** | **55037 (50388, 68815)** | **11249362 (11249331, 11249391)** | **-51767 (-70450,-32655)** | **691.89 (340.01,988.93)** | **Cost-saving** | **Lowest cost scenario** | **65604499 (39548722, 86494121)** | **72523434 (43319700, 95256343)** |
| PV 54 vaccines per day | 234 [0,1101] | 340 [0,1641] | 55546 (51254, 69157) | 11249363 (11249332, 11249392) | -51257 (-69648,-32042) | 692.92 (341.06,989.07) | Cost-saving | Extendedly dominated | 65115210 (38739358, 85769175) | 72044430 (42529320, 94541826) |
| PV 81 vaccines per day | 221 [0,1064] | 325 [0,1594] | 55819 (51721, 69602) | 11249364 (11249332, 11249392) | -50984 (-69268,-31616) | 693.54 (341.69,990.59) | Cost-saving | Extendedly dominated | 64855010 (38434977, 85365542) | 71790365 (42126202, 94470822) |
| PV 135 vaccines per day | 203 [0,1022] | 305 [0,1523] | 55937 (52091, 69054) | 11249364 (11249332, 11249392) | -50866 (-69366,-31562) | 693.81 (342.11,991.26) | Cost-saving | 469,171/QALY compared to PV 41/day | 64741961 (38276996, 85404615) | 71680031 (41939734, 94628481) |
| Reactive vaccination (RV) of GBMSM at high risk for mpox who attend SHS | | | | | | | | | | |
| RV 27 vaccines per day | 9841 [7021,13064] | 4164 [3745,4450] | 71336 (61702, 79518) | 11249253 (11249174, 11249324) | -35468 (-49843,-17058) | 582.20 (248.87,835.36) | Cost-saving | Dominated | 47112380 (21522760, 65341813) | 52934331 (24018780, 72199674) |
| RV 41 vaccines per day | 5597 [4074,7416] | 3596 [3171,3909] | 67469 (58414, 75314) | 11249306 (11249255, 11249354) | -39334 (-54577,-20770) | 636.02 (283.78,915.49) | Cost-saving | Dominated | 52054532 (25996865, 71214390) | 58414758 (28857042, 79563566) |
| RV 81 vaccines per day | 2157 [1747,2723] | 2487 [2060,2842] | 62940 (52764, 72063) | 11249348 (11249316, 11249380) | -43863 (-59407,-27877) | 677.78 (313.03,979.49) | Cost-saving | Dominated | 57418970 (33462080, 76892754) | 64196772 (36817085, 85867340) |

| | | | | | | | | | | |
|---|---|---|---|---|---|---|---|---|---|---|
| RV 135 vaccines per day | 1284 [1071,1605] | 1699 [1370,2233] | 59097 (48719, 71958) | 11249352 (11249322, 11249383) | -47706 (-64057,-31558) | 681.78 (317.00,988.13) | Cost-saving | Dominated | 61342013 (38060217, 80820324) | 68159842 (41348524, 89922723) |
| RV 204 vaccines per day | 960 [729,1565] | 1328 [966,2148] | 58108 (46070, 72592) | 11249354 (11249327, 11249383) | -48695 (-65793,-34316) | 683.77 (319.35,987.23) | Cost-saving | Dominated | 62370473 (41417668, 80930669) | 69208189 (44716860, 90590305) |
| RV 465 vaccines per day | 811 [462,1530] | 1130 [630,2111] | 59240 (50016, 72731) | 11249356 (11249327, 11249384) | -47564 (-65145,-31640) | 685.22 (326.32,988.04) | Cost-saving | Dominated | 61268046 (39093258, 80641798) | 68120249 (42233645, 89083241) |

Cost-saving scenarios save money and gain QALYs compared to the no vaccination counterfactual scenario. Dominated scenarios have higher mean costs and save fewer QALYs (just comparison of mean) than another scenario. Extended dominated scenarios have a higher mean ICER and save fewer QALYs (just comparison of mean) than another scenario. The scenario in bold is the most cost-effective vaccination scenario – it saves most money and no other scenario that saves more QALYs is cost-effective compared to it, it also has the largest INMB. ICER is calculated using the mean incremental QALY and costs.

Abbreviations: QALY, Quality Adjusted Life Years; ICER, incremental cost-effectiveness ratio; INMB, incremental net monetary benefit; PV, pre-emptive vaccination; RV, reactive vaccination

Table S15 Threshold vaccine price in British pounds 2022 that meet the NICE standard for ICER.

| Daily rate of vaccination | Baseline | | Discount 1.5% | | Exclusive of PHR | | Inclusive of PL | | Long immunity | | Short immunity | | Low importation | | High importation | | Low OTB threshold | | High OTB threshold | | No behavior change | | Physical contact | | Less severe | |
|---|---|---|---|---|---|---|---|---|---|---|---|---|---|---|---|---|---|---|---|---|---|---|---|---|---|---|
| | V50 | V90 | V50 | V90 | V50 | V90 | V50 | V90 | V50 | V90 | V50 | V90 | V50 | V90 | V50 | V90 | V50 | V90 | V50 | V90 | V50 | V90 | V50 | V90 | V50 | V90 |
| Pre-emptive vaccination of GBMSM at high risk for mpox who attend SHS | | | | | | | | | | | | | | | | | | | | | | | | | | |
| 13 | 701 | 573 | 687 | 624 | 356 | 295 | 1351 | 1060 | 514 | 493 | 621 | 483 | 696 | 676 | 506 | 490 | 564 | 299 | 715 | 685 | 2095 | 1998 | 153 | 123 | 650 | 593 |
| 27 | 485 | 455 | 511 | 482 | 196 | 171 | 867 | 774 | 412 | 389 | 515 | 296 | 515 | 488 | 368 | 172 | 416 | 273 | 530 | 515 | 2105 | 1943 | 181 | 160 | 484 | 455 |
| 41 | 400 | 368 | 423 | 390 | 129 | 113 | 668 | 589 | 401 | 378 | 421 | 285 | 367 | 335 | 323 | 221 | 353 | 258 | 424 | 394 | 1718 | 1549 | 209 | 185 | 400 | 367 |
| 54 | 392 | 362 | 415 | 384 | 126 | 110 | 655 | 576 | 396 | 373 | 361 | 265 | 357 | 326 | 321 | 220 | 348 | 252 | 415 | 385 | 1688 | 1515 | 168 | 161 | 392 | 362 |
| 81 | 388 | 358 | 412 | 380 | 124 | 109 | 648 | 570 | 392 | 370 | 296 | 223 | 352 | 322 | 317 | 219 | 346 | 250 | 411 | 381 | 1675 | 1500 | 108 | 103 | 387 | 357 |
| 136 | 385 | 354 | 409 | 379 | 123 | 108 | 643 | 568 | 389 | 369 | 292 | 220 | 349 | 319 | 317 | 219 | 344 | 248 | 408 | 380 | 1662 | 1489 | 96 | 91 | 385 | 354 |
| Reactive vaccination of GBMSM at high risk for mpox who attend SHS | | | | | | | | | | | | | | | | | | | | | | | | | | |
| 27 | 616 | 567 | 647 | 590 | 325 | 296 | 1220 | 1076 | 514 | 499 | 577 | 329 | 734 | 711 | 419 | 102 | 517 | 253 | 696 | 666 | 2131 | 1978 | 150 | 127 | 538 | 493 |
| 41 | 546 | 509 | 572 | 534 | 267 | 247 | 1051 | 943 | 477 | 457 | 520 | 301 | 654 | 636 | 375 | 136 | 453 | 242 | 613 | 590 | 1876 | 1731 | 160 | 143 | 479 | 449 |
| 81 | 464 | 437 | 488 | 461 | 194 | 186 | 839 | 782 | 467 | 419 | 412 | 276 | 562 | 537 | 328 | 191 | 386 | 255 | 518 | 499 | 1843 | 1723 | 185 | 165 | 413 | 392 |
| 136 | 445 | 410 | 470 | 433 | 167 | 162 | 778 | 717 | 439 | 404 | 356 | 256 | 532 | 500 | 310 | 194 | 367 | 268 | 499 | 473 | 1795 | 1723 | 196 | 181 | 399 | 370 |
| 204 | 426 | 394 | 450 | 417 | 151 | 147 | 730 | 676 | 426 | 394 | 337 | 250 | 523 | 489 | 308 | 196 | 352 | 260 | 483 | 451 | 1770 | 1638 | 200 | 182 | 383 | 357 |
| 465 | 396 | 371 | 422 | 394 | 139 | 129 | 679 | 629 | 394 | 375 | 307 | 224 | 437 | 422 | 307 | 194 | 341 | 246 | 433 | 418 | 1745 | 1604 | 187 | 176 | 363 | 338 |

Two NICE prices are obtained: V50 represents the price at which the median ICER is just below £20k/QALY, and V90 the price at which the 90% of ICER is just below £30k/QALY. Scenarios are defined briefly below:

1. Discount 1.5%: Lower annual discount rate (1.5%) on costs and utilities;
2. Exclusive of PHR: Only include direct health care costs so remove public health response costs from the baseline;
3. Inclusive of PL: Include productivity losses due to mpox disease instead of just health care and public health response costs;
4. Long immunity: Long duration of protection (Limm1=10 years and Limm2=Limm0=20 years);
5. Short immunity: Short duration of protection (Limm1=2.5 years and Limm2=Limm0=5.0 years);
6. Low importation: Low importation rate (seeding rate=1 case per month);
7. High importation: High importation rate (seeding rate=10 cases per month);
8. Low OTB threshold: Low outbreak criteria;
9. High OTB threshold: High outbreak criteria;
10. No behaviour change: No responsive behaviour changes during outbreaks instead of assuming a decrease in sexual risk behaviour during each outbreak;
11. Physical contact: Using data from the RiiSH MPOX survey in December 2022 to define the contact matrix;
12. Less severe: Breakthrough infections have less severe symptoms - half the chance of experiencing moderate and severe mpox disease compared to the baseline scenario

Table S16 Cost effectiveness of vaccination in the baseline scenario with the total costs excluding Public Health Response (PHR) costs. The values listed are mean and 95%CrI.

| Scenario | Total number of infections | Outbreak duration (days) | Total Cost (1000s £) | Total QALYs (1000s QALY) | Incremental costs compared to no vaccination (1000s £) | Incremental QALY compared to no vaccination | ICER compared to no vaccination | Fully incremental analysis | INMB compared to No Vaccination at £20,000/QALY | INMB compared to no vaccination at £30,000/QALY |
|---|---|---|---|---|---|---|---|---|---|---|
| No Vaccination counterfactual | 54891 [26246,67256] | 5472 [5141,5676] | 25043 (13228, 31583) | 9493798 (9493540, 9494085) | – | – | – | Dominated | – | – |
| Pre-emptive vaccination (PV) of GBMSM at high risk for mpox who attend SHS | | | | | | | | | | |
| PV 13 vaccines per day | 13099 [8224,18452] | 4219 [3494,4673] | 21316 (19243, 23677) | 9494228 (9494138, 9494307) | -3727 (-8262, 6596) | 429.59 (182.58,614.47) | Cost-saving | Lowest cost scenario | 12318458 (-3025803, 19743681) | 16614387 (-1247030, 25908743) |
| PV 27 vaccines per day | 2143 [637,3860] | 2210 [802,3198] | 31060 (30616, 31681) | 9494331 (9494299, 9494358) | 6017 (-33, 17634) | 532.62 (259.29,767.60) | Not Cost-saving | £212,868/QALY compared to RV41 | 4635435 (-12183024, 14664829) | 9961661 (-9484032, 21962679) |
| PV 41 vaccines per day | 254 [0,1143] | 369 [0,1719] | 43110 (42494, 43627) | 9494345 (9494318, 9494369) | 18067 (11865, 29407) | 546.26 (278.56,780.80) | Not Cost-saving | Extended dominated | -7141685 (-23480194, 3140300) | -1679094 (-20722994, 10590648) |
| PV 54 vaccines per day | 234 [0,1101] | 340 [0,1641] | 43948 (43356, 44489) | 9494346 (9494319, 9494369) | 18905 (12631, 30268) | 547.23 (279.58,780.85) | Not Cost-saving | Extendedly dominated | -7960175 (-24318269, 2376985) | -2487831 (-21539234, 9645257) |
| PV 81 vaccines per day | 221 [0,1064] | 325 [0,1594] | 44425 (43839, 44954) | 9494346 (9494319, 9494370) | 19382 (13088, 30752) | 547.84 (280.23,782.34) | Not Cost-saving | £93,027/QALY compared to RV 135/day | -8425454 (-24797615, 1909729) | -2947101 (-22001629, 9218818) |
| PV 135 vaccines per day | 203 [0,1022] | 305 [0,1523] | 44812 (44227, 45336) | 9494346 (9494320, 9494370) | 19769 (13434, 31140) | 548.10 (280.66,783.00) | Not Cost-saving | £1,486,765/QALY compared to PV 81/day | -8806621 (-25182151, 1466922) | -3325571 (-22405152, 8858433) |
| Reactive vaccination (RV) of GBMSM at high risk for mpox who attend SHS | | | | | | | | | | |
| **RV 27 vaccines per day** | **9841 [7021,13064]** | **4164 [3745,4450]** | **21442 (18514, 24141)** | **9494257 (9494192, 9494315)** | **-3601 (-8176, 6101)** | **458.51 (202.78,658.39)** | **Cost-saving** | **£4,335/QALY compared to PV13** | **12771380 (-2249939, 20566422)** | **17356447 (-213042, 27014527)** |
| RV 41 vaccines per day | 5597 [4074,7416] | 3596 [3171,3909] | 24333 (21146, 27267) | 9494299 (9494257, 9494339) | -710 (-5501, 8770) | 501.02 (231.93,722.24) | Cost-saving | £68,023/QALY compared to RV27 | 10730068 (-4315754, 19089368) | 15740302 (-1868298, 26120098) |

| | | | | | | | | | | |
|---|---|---|---|---|---|---|---|---|---|---|
| RV 81 vaccines per day | 2157 [1747,2723] | 2487 [2060,2842] | 30979 (25823, 35351) | 9494333 (9494306, 9494360) | 5936 (1363, 13493) | 534.38 (253.95,772.71) | Not Cost-saving | Extendedly dominated | 4751873 (-8929097, 12702524) | 10095629 (-6485071, 20327076) |
| RV 135 vaccines per day | 1284 [1071,1605] | 1699 [1370,2233] | 34785 (28352, 41729) | 9494336 (9494311, 9494362) | 9742 (5218, 15887) | 537.53 (256.33,780.15) | Not Cost-saving | £758,658/QALY compared to PV27 | 1008525 (-11177673, 8343789) | 6383803 (-9006781, 16326526) |
| RV 204 vaccines per day | 960 [729,1565] | 1328 [966,2148] | 37358 (30062, 42898) | 9494338 (9494315, 9494362) | 12315 (7407, 17706) | 539.28 (259.57,777.96) | Not Cost-saving | Extended dominated | -1529734 (-13074335, 5710612) | 3863035 (-10481092, 13499388) |
| RV 465 vaccines per day | 811 [462,1530] | 1130 [630,2111] | 40311 (36155, 43787) | 9494339 (9494315, 9494363) | 15269 (10379, 23496) | 540.67 (262.46,778.53) | Not Cost-saving | Extended dominated | -4455271 (-18013933, 4138630) | 951390 (-1553317, 11904118) |

Cost-saving scenarios save money and gain QALYs compared to the no vaccination counterfactual scenario. Dominated scenarios have higher mean costs and save fewer QALYs (just comparison of mean) than another scenario. Extended dominated scenarios have a higher mean ICER and save fewer QALYs (just comparison of mean) than another scenario. The scenario in bold is the most cost-effective vaccination scenario. Note that PV13 saves most money, but RV27 gains more QALYs, is cost-effective compared to PV13 and has the largest INB and so RV27 is the most cost-effective vaccination scenario.

Abbreviations: QALY, Quality Adjusted Life Years; ICER, incremental cost-effectiveness ratio; INMB, incremental net monetary benefit; PV, pre-emptive vaccination; RV, reactive vaccination

Table S17 Cost effectiveness of vaccination in the baseline scenario with the total costs including Productivity losses (PL). The values listed are mean and 95% CrI.

| Scenario | Total number of infections | Outbreak duration (days) | Total Cost (1000s £) | Total QALYs | Incremental costs compared to no vaccination (1000s £) | Incremental QALY compared to no vaccination | ICER compared to no vaccination | Fully incremental analysis | INMB compared to No Vaccination at £20,000/QALY | INMB compared to no vaccination at £30,000/QALY |
|---|---|---|---|---|---|---|---|---|---|---|
| No Vaccination counterfactual | 54891 [26246,67256] | 5472 [5141,5676] | 147333 (101595, 176598) | 9493798 (9493540, 9494085) | – | – | – | Dominated | – | – |
| Pre-emptive vaccination (PV) of GBMSM at high risk for mpox who attend SHS | | | | | | | | | | |
| PV 13 vaccines per day | 13099 [8224,18452] | 4219 [3494,4673] | 76964 (61821, 90753) | 9494228 (9494138, 9494307) | -70370 (-88693,-32093) | 429.59 (182.58,614.47) | Cost-saving | Dominated | 78961397 (34870135, 99786313) | 83257326 (36429607, 105296668) |
| PV 27 vaccines per day | 2143 [637,3860] | 2210 [802,3198] | 56600 (43129, 68195) | 9494331 (9494299, 9494358) | -90733 (-111895,-52828) | 532.62 (259.29,767.60) | Cost-saving | Dominated | 101385791 (57649900, 126181083) | 106712018 (60557098, 132681917) |
| **PV 41 vaccines per day** | **254 [0,1143]** | **369 [0,1719]** | **52423 (46855, 64735)** | **9494345 (9494318, 9494369)** | **-94910 (-118102,-53538)** | **546.26 (278.56,780.80)** | **Most cost-effective** | **Lowest cost scenario** | **105835068 (58457980, 132346837)** | **111297659 (61012615, 140105486)** |
| PV 54 vaccines per day | 234 [0,1101] | 340 [0,1641] | 52850 (47636, 64662) | 9494346 (9494319, 9494369) | -94483 (-117435,-52811) | 547.23 (279.58,780.85) | Cost-saving | Extendedly dominated | 105427416 (57706416, 132090743) | 110899760 (60269552, 139700982) |
| PV 81 vaccines per day | 221 [0,1064] | 325 [0,1594] | 53085 (48055, 65012) | 9494346 (9494319, 9494370) | -94248 (-117511,-52901) | 547.84 (280.23,782.34) | Cost-saving | 418,780/QALY compared to PV41 | 105204891 (57307675, 131900237) | 110683245 (59859076, 139584315) |
| PV 135 vaccines per day | 203 [0,1022] | 305 [0,1523] | 53201 (48396, 65003) | 9494346 (9494320, 9494370) | -94133 (-117675,-52561) | 548.10 (280.66,783.00) | Cost-saving | 444,642/QALY compared to PV81 | 105094677 (57043593, 131616252) | 110575727 (60066695, 139310911) |
| Reactive vaccination (RV) of GBMSM at high risk for mpox who attend SHS | | | | | | | | | | |
| RV 27 vaccines per day | 9841 [7021,13064] | 4164 [3745,4450] | 73463 (62597, 83515) | 9494257 (9494192, 9494315) | -73870 (-4164,-32670) | 458.51 [202.78,658.39] | Cost-saving | Dominated | 83040035 (36806093, 105678638) | 87625102 (38921420, 112018696) |
| RV 41 vaccines per day | 5597 [4074,7416] | 3596 [3171,3909] | 66266 (56994, 74805) | 9494299 (9494257, 9494339) | -81067 (-103325,-39129) | 501.02 [231.93,722.24] | Cost-saving | Dominated | 91087659 (43793042, 115676015) | 96097894 (46119456, 122808498) |

| | | | | | | | | | | |
|---|---|---|---|---|---|---|---|---|---|---|
| RV 81 vaccines per day | 2157 [1747,2723] | 2487 [2060,2842] | 59584 (50623, 67844) | 9494333 (9494306, 9494360) | -87749 (-110894,-46647) | 534.38 [253.95,772.71] | Cost-saving | Dominated | 98436973 (51723115, 124729343) | 103780729 (54439133, 132267893) |
| RV 135 vaccines per day | 1284 [1071,1605] | 1699 [1370,2233] | 56342 (47477, 66815) | 9494336 (9494311, 9494362) | -90991 (-113712,-50483) | 537.53 [256.33,780.15] | Cost-saving | Dominated | 101741742 (55771817, 128588592) | 107117019 (58284579, 136423880) |
| RV 204 vaccines per day | 960 [729,1565] | 1328 [966,2148] | 55531 (45324, 67622) | 9494338 (9494315, 9494362) | -91802 (-113229,-53292) | 539.28 [259.57,777.96] | Cost-saving | Dominated | 102587469 (58498152, 127811749) | 107980238 (60803667, 135703148) |
| RV 465 vaccines per day | 811 [462,1530] | 1130 [630,2111] | 5646 (47905, 67990) | 9494339 (9494315, 9494363) | -90876 (-112564,-51792) | 540.67 [262.46,778.53] | Cost-saving | Dominated | 101689035 (57123872, 127072912) | 107095695 (59525929, 135030064) |

Cost-saving scenarios save money and gain QALYs compared to the no vaccination counterfactual scenario. Dominated scenarios have higher mean costs and save fewer QALYs (just comparison of mean) than another scenario. Extended dominated scenarios have a higher mean ICER and save fewer QALYs (just comparison of mean) than another scenario. The scenario in bold is the most cost-effective vaccination scenario – it saves most money and no other scenario that saves more QALYs is cost-effective compared to it. PV 41 also has the highest INMB.

Abbreviations: QALY, Quality Adjusted Life Years; ICER, incremental cost-effectiveness ratio; INMB, incremental net monetary benefit; PV, pre-emptive vaccination; RV, reactive vaccination.

Table S18 Cost effectiveness of vaccination under long immunity with $L_{imm1}$=10 and $L_{imm2}$=$L_{imm0}$=20 years

| Scenario | Total number of infections | Outbreak duration (days) | Total Cost (1000s £) | Total QALYs | Incremental costs compared to no vaccination (1000s £) | Incremental QALY compared to no vaccination | ICER compared to no vaccination | Fully incremental analysis | INMB compared to No Vaccination at £20,000/QALY | INMB compared to no vaccination at £30,000/QALY |
|---|---|---|---|---|---|---|---|---|---|---|
| No Vaccination counterfactual | 26601 (20789,31794) | 4653 (4152,4958) | 57379 (48234, 66194) | 9494106 (9493967, 9494224) | – | – | – | Dominated | – | – |
| Pre-emptive vaccination (PV) of GBMSM at high risk for mpox who attend SHS | | | | | | | | | | |
| PV 13 vaccines per day | 2154 (743,3732) | 2271 (1029,3157) | 35964 (24122, 45387) | 9494332 (9494299, 9494360) | -21415 (-27798,-15410) | 225.60 (135.25,336.78) | Cost-saving | Dominated | 25926941 (20064182, 32123704) | 28182957 (22218528, 34556183) |
| **PV 27 vaccines per day** | **261 (0,1023)** | **379 (0,1585)** | **30194 (25987, 40401)** | **9494344 (9494314, 9494369)** | **-27185 (-33851, -19872)** | **237.50 (142.62,355.27)** | **Cost-saving** | **Lowest cost scenario** | **31934667 (24747126, 39044114)** | **34309634 (27039150, 41881949)** |
| PV 41 vaccines per day | 226 (0,986) | 327 (0,1490) | 30455 (26848, 40453) | 9494345 (9494316, 9494370) | -26924 (-33945, -19744) | 238.70 (142.87,354.59) | Cost-saving | £217,500/QALY compared to PV27 | 31697959 (24660396, 38990331) | 34084951 (26541244, 41930864) |
| PV 54 vaccines per day | 216 (0,965) | 315 (0,1443) | 30668 (27210, 40279) | 9494346 (9494316, 9494371) | -26711 (-33801, -19636) | 239.23 (143.15,355.37) | Cost-saving | Extendedly dominated | 31495534 (24310508, 38962409) | 33887794 (26317583, 41919995) |
| PV 81 vaccines per day | 202 (0,936) | 298 (0,1395) | 30800 (27522, 40101) | 9494346 (9494317, 9494371) | -26579 (-33518, -19483) | 239.61 (143.39,355.44) | Cost-saving | Extendedly dominated | 31371255 (24012832, 38708080) | 33767356 (25985908, 41676631) |
| PV 135 vaccines per day | 187 (0,894) | 282 (0,1338) | 30875 (27793, 39775) | 9494346 (9494316, 9494371) | -26504 (-33334, -19305) | 239.86 (143.58,355.93) | Cost-saving | 362,069/QALY compared to PV41 | 31301452 (23780413, 38518162) | 33700078 (25722569, 41494161) |
| Reactive vaccination (RV) of GBMSM at high risk for mpox who attend SHS | | | | | | | | | | |
| RV 27 vaccines per day | 3639 (2788,4697) | 3044 (2611,3424) | 40800 (34370, 46980) | 9494320 (9494284, 9494352) | -16579 (-20963,-12856) | 213.47 (124.45,323.13) | Cost-saving | Dominated | 20848611 (16362169, 25904084) | 22983340 (17885998, 28498052) |
| RV 41 vaccines per day | 2253 (1786,2893) | 2530 (2137,2905) | 38710 (32269, 45438) | 9494333 (9494306, 9494359) | -18669 (-23370, -14488) | 226.33 (131.76,344.43) | Cost-saving | Dominated | 23195497 (18434339, 28532858) | 25458756 (20042351, 31460308) |

| | | | | | | | | | | |
|---|---|---|---|---|---|---|---|---|---|---|
| RV 81 vaccines per day | 1120 (926,1429) | 1572 (1248,1992) | 34215 (27169, 42669) | 9494337 (9494313, 9494362) | -23164 (-28782,-17521) | 230.23 (133.58,356.15) | Cost-saving | Dominated | 27768527 (22550400, 33641271) | 30070786 (24374523, 36365355) |
| RV 135 vaccines per day | 765 (587,1292) | 1121 (817,1851) | 32565 (25093, 42763) | 9494337 (9494314, 9494363) | -24814 (-30731, -18736) | 231.14 (135.43,355.34) | Cost-saving | Dominated | 29437024 (23669095, 35644439) | 31748434 (25935370, 38418098) |
| RV 204 vaccines per day | 644 (421,1254) | 938 (619,1842) | 32317 (25358, 43020) | 9494337 (9494312, 9494362) | -25062 (-31122, -18102) | 231.00 (135.07,356.48) | Cost-saving | Dominated | 29682142 (23234768, 35979476) | 31992124 (25699199, 38862872) |
| RV 465 vaccines per day | 573 (259,1231) | 848 (384,1827) | 33189 (25838, 43162) | 9494340 (9494315, 9494365) | -24190 (-30873, -17811) | 233.52 (138.71,356.49) | Cost-saving | Dominated | 28860843 (22427845, 35614122) | 31196024 (24615828, 38738830) |

Cost-saving scenarios save money and gain QALYs compared to the no vaccination counterfactual scenario. Dominated scenarios have higher mean costs and save fewer QALYs (just comparison of mean) than another scenario. Extended dominated scenarios have a higher mean ICER and save fewer QALYs (just comparison of mean) than another scenario. The scenario in bold is the most cost-effective vaccination scenario – it saves most money and no other scenario that saves more QALYs is cost-effective compared to it; PV27 also has the highest INMB.

Abbreviations: QALY, Quality Adjusted Life Years; ICER, incremental cost-effectiveness ratio; INMB, incremental net monetary benefit; PV, pre-emptive vaccination; RV, reactive vaccination

Table S19 Cost effectiveness of vaccination under short immunity with $L_{imm1}$=2.5 and $L_{imm2}$=$L_{imm0}$=5.0 years

| Scenario | Total number of infections | Outbreak duration (days) | Total Cost (1000s £) | Total QALYs | Incremental costs compared to no vaccination (1000s £) | Incremental QALY compared to no vaccination | ICER compared to no vaccination | Fully incremental analysis | INMB compared to No Vaccination at £20,000/QALY | INMB compared to no vaccination at £30,000/QALY |
|---|---|---|---|---|---|---|---|---|---|---|
| No Vaccination counterfactual | 82021 (20560,108195) | 6084 (5836,6597) | 115004 (75405,141882) | 9493465 (9492889, 9494131) | – | – | – | Dominated | – | – |
| Pre-emptive vaccination (PV) of GBMSM at high risk for mpox who attend SHS | | | | | | | | | | |
| PV 13 vaccines per day | 44117 (33050,58818) | 5669 (5426,5889) | 96033 (84380, 110084) | 9493886 (9493581, 9494109) | -18971 (-33113,16296) | 421.45 (-105.08,723.73) | Cost-saving | Dominated | 27400267 (-18401811, 44956858) | 31614781 (-19457023, 51282082) |
| PV 27 vaccines per day | 16823 (11283,24683) | 4825 (4439,5208) | 82421 (74219, 92178) | 9494181 (9494036, 9494282) | -32583 (-53378,7222) | 715.56 (64.25,1174.41) | Cost-saving | Dominated | 46894254 (-6089066, 72828740) | 54049855 (-5643951, 83343889) |
| PV 41 vaccines per day | 5742 (3394,9406) | 3667 (2922,4284) | 78648 (70020, 86703) | 9494297 (9494232, 9494343) | -36356 (-59197, 13220) | 832.28 (176.04,1354.56) | Cost-saving | Dominated | 53001926 (2075367, 82336684) | 61324709 (3751320, 91944861) |
| **PV 54 vaccines per day** | **1937 (655,3706)** | **2187 (885,3266)** | **77724 (65971, 88174)** | **9494331 (9494301, 9494360)** | **-37280 (-60290, -2660)** | **866.31 (208.01,1412.63)** | **Cost-saving** | **Lowest cost scenario** | **54606422 (6100753, 83741379)** | **63269504 (8012726, 96417055)** |
| PV 81 vaccines per day | 181 (0,850) | 239 (0,1139) | 81333 (78500, 89271) | 9494345 (9494316, 9494371) | -33671 (-57511, 4506) | 880.35 (222.72,1427.94) | Cost-saving | Extendedly dominated | 51278248 (251012, 81900742) | 60081792 (2478644, 93713287) |
| PV 135 vaccines per day | 156 (0,780) | 215 (0,1054) | 82065 (79477, 89516) | 9494346 (9494318, 9494371) | -32939 (-56934, 5370) | 881.32 (223.20,1428.29) | Cost-saving | Extendedly dominated | 50565710 ( -629267, 80970157) | 59378916 (1608181, 92842374) |
| Reactive vaccination (RV) of GBMSM at high risk for mpox who attend SHS | | | | | | | | | | |
| RV 27 vaccines per day | 28202 (20657,38238) | 5390 (5141,5636) | 88545 (79064, 99916) | 9494053 (9493845, 9494202) | -26459 (-44657,14150) | 587.80 (-35.59,981.13) | Cost-saving | Dominated | 38215567 (-14895102, 61023132) | 44093612 (-15267429, 69874977) |
| RV 41 vaccines per day | 16312 (12056,22294) | 4879 (4613,5140) | 82691 (74912, 91403) | 9494182 (9494051, 9494278) | -32313 (-53215,8110) | 716.86 (70.40,1186.48) | Cost-saving | Dominated | 46650110 (-7033023, 72921142) | 53818716 (-6385214, 82571598) |

| | | | | | | | | | | |
|---|---|---|---|---|---|---|---|---|---|---|
| RV 81 vaccines per day | 5659 (4412,7366) | 3750 (3453,4046) | 80378 (73249, 87676) | 9494297 (9494251, 9494340) | -34627 (-57596, 4134) | 832.36 (176.56, 1360.96) | Cost-saving | Dominated | 51273703 (-1531029, 80981727) | 5959729 (-61698, 90982279) |
| RV 135 vaccines per day | 2665 (2234,3349) | 2812 (2522,3093) | 81488 (72902, 89953) | 9494329 (9494302, 9494359) | -33516 (-56517, 4031) | 864.25 (205.48,1414.23) | Cost-saving | Dominated | 5080089 (-167059, 81148369) | 59443391 (1488039, 91214937) |
| RV 204 vaccines per day | 1743 (1542,2015) | 2083 (1821,2411) | 80991 (70978, 92531) | 9494335 (9494309, 9494361) | -34013 (-56251, 2314) | 869.60 (207.71,1433.19) | Cost-saving | Extendedly dominated | 51405521 (838051, 80373550) | 60101556 (3058947, 92404004) |
| RV 465 vaccines per day | 1175 (832,1812) | 1484 (1024,2251) | 83550 (73380, 95882) | 9494337 (9494312, 9494363) | -31454 (-54343, 5067) | 872.04 (212.12,1432.77) | Cost-saving | £1016,764/QALY compared to PV54 | 48895042 (-328887, 78539649) | 57615362 (1758239, 88897623) |

Cost-saving scenarios save money and gain QALYs compared to the no vaccination counterfactual scenario. Dominated scenarios have higher mean costs and save fewer QALYs (just comparison of mean) than another scenario. Extended dominated scenarios have a higher mean ICER and save fewer QALYs (just comparison of mean) than another scenario. The scenario in bold is the most cost-effective vaccination scenario – it saves most money and no other scenario that saves more QALYs is cost-effective compared to it; PV54 also has the highest INMB.

Abbreviations: QALY, Quality Adjusted Life Years; ICER, incremental cost-effectiveness ratio; INMB, incremental net monetary benefit; PV, pre-emptive vaccination; RV, reactive vaccination

Table S20 Cost effectiveness of vaccination under the low importation rate with seeding rate =1 case per month

| Scenario | Total number of infections | Outbreak duration (days) | Total Cost (1000s £) | Total QALYs | Incremental costs compared to no vaccination (1000s £) | Incremental QALY compared to no vaccination | ICER compared to no vaccination | Fully incremental analysis | INMB compared to No Vaccination at £20,000/QALY | INMB compared to no vaccination at £30,000/QALY |
|---|---|---|---|---|---|---|---|---|---|---|
| No Vaccination counterfactual | 52295 (40702,63264) | 4476 (4105,4908) | 74445 (58982, 90504) | 9493836 (9493607, 9494072) | – | – | – | Dominated | – | – |
| Pre-emptive vaccination (PV) of GBMSM at high risk for mpox who attend SHS | | | | | | | | | | |
| PV 13 vaccines per day | 9168 (4137,14894) | 2860 (2084,3647) | 43810 (33583, 56611) | 9494281 (9494197, 9494350) | -30635 (-40008,-23307) | 444.80 (268.50,618.14) | Cost-saving | Dominated | 39530928 (30360695, 50865374) | 43978909 (33726990, 56135163) |
| **PV 27 vaccines per day** | **392 (0,1706)** | **521 (0,1808)** | **33344 (28638, 44983)** | **9494371 (9494346, 9494393)** | **-41101 (-53031, -30191)** | **535.23 (317.56,741.04)** | **Cost-saving** | **Lowest cost scenario** | **51805344 (38539168, 65936712)** | **57157659 (42561418, 72782168)** |
| PV 41 vaccines per day | 8 (0,73) | 6 (0,97) | 41395 (41125, 42821) | 9494391 (9494379, 9494398) | -33049 (-48824, -17806) | 555.23 (324.39,776.37) | Cost-saving | 402,600/QALY compared to PV27 | 44153757 (26187697, 63360732) | 49706020 (30678358, 70421491) |
| PV 54 vaccines per day | 8 (0,71) | 6 (0,97) | 42340 (42089, 43693) | 9494392 (9494380, 9494398) | -32105 (-47886, -16843) | 555.67 (324.54,777.33) | Cost-saving | Extendedly dominated | 43217998 (25224542, 62436290) | 48774661 (29721433, 69503491) |
| PV 81 vaccines per day | 7 (0,68) | 6 (0,95) | 42836 (42598, 44165) | 9494392 (9494380, 9494398) | -31609 (-47391, -16334) | 555.91 (324.62,777.83) | Cost-saving | £2117,647/QALY compared to PV41 | 42726799 (24714904, 61949472) | 48285934 (29215086, 69020389) |
| PV 135 vaccines per day | 7 (0,62) | 5 (0,94) | 43228 (43004, 44526) | 9494392 (9494380, 9494398) | -31216 (-46997, -15928) | 556.09 (324.68,778.01) | Cost-saving | £2183,333/QALY compared to PV81 | 42338336 (24309719, 61561398) | 47899273 (28812151, 68634638) |
| Reactive vaccination (RV) of GBMSM at high risk for mpox who attend SHS | | | | | | | | | | |
| RV 27 vaccines per day | 10372 (6863,14054) | 3297 (2870,3753) | 47310 (38707, 57308) | 9494261 (9494188, 9494325) | -27134 (-36366,-19210) | 424.81 (252.26,593.90) | Cost-saving | Dominated | 35630698 (25835893, 46529080) | 39878839 (28548658, 52123343) |
| RV 41 vaccines per day | 5706 (3743,7997) | 2841 (2404,3259) | 43953 (35699, 53194) | 9494309 (9494262, 9494351) | -30492 (-40774, -22022) | 473.16 (276.05,660.57) | Cost-saving | Dominated | 39955047 (29253262, 52394336) | 44686606 (32291501, 59094571) |

| | | | | | | | | | | |
|---|---|---|---|---|---|---|---|---|---|---|
| RV 81 vaccines per day | 1859 (1402,2530) | 1946 (1558,2346) | 40035 (31856, 49598) | 9494350 (9494327, 9494370) | -34409 (-45644, -25446) | 514.00 (296.59,721.37) | Cost-saving | Dominated | 44689246 (33248210, 58004033) | 49829294 (36807596, 65180504) |
| RV 135 vaccines per day | 919 (789,1102) | 1332 (1020,1704) | 37693 (28831, 48433) | 9494358 (9494340, 9494376) | -36751 (-47736, -27666) | 522.09 (300.88,738.88) | Cost-saving | Dominated | 47193206 (35817899, 60917532) | 52414088 (39765211, 67971872) |
| RV 204 vaccines per day | 617 (524,729) | 944 (751,1180) | 36036 (28387, 45839) | 9494360 (9494342, 9494377) | -38409 (-50057, -28924) | 524.16 (303.94,641.29) | Cost-saving | Dominated | 48892094 (37422423, 63250514) | 54133677 (41191372, 70447698) |
| RV 465 vaccines per day | 343 (270,472) | 546 (403,888) | 38159 (30399, 47435) | 9494371 (9494354, 9494384) | -36286 (-48852, -26342) | 534.82 (310.39,750.27) | Cost-saving | Dominated | 46982204 (35089050, 61980846) | 52330415 (38700808, 69137634) |

Cost-saving scenarios save money and gain QALYs compared to the no vaccination counterfactual scenario. Dominated scenarios have higher mean costs and save fewer QALYs (just comparison of mean) than another scenario. Extended dominated scenarios have a higher mean ICER and save fewer QALYs (just comparison of mean) than another scenario. The scenario in bold is the most cost-effective vaccination scenario – it saves most money and no other scenario that saves more QALYs is cost-effective compared to it; PV27 also has the highest INMB.

Abbreviations: QALY, Quality Adjusted Life Years; ICER, incremental cost-effectiveness ratio; INMB, incremental net monetary benefit; PV, pre-emptive vaccination; RV, reactive vaccination.

Table S21 Cost effectiveness of vaccination under the high importation rate with seeding rate =10 cases per month

| Scenario | Total number of infections | Outbreak duration (days) | Total Cost (1000s £) | Total QALYs | Incremental costs compared to no vaccination (1000s £) | Incremental QALY compared to no vaccination | ICER compared to no vaccination | Fully incremental analysis | INMB compared to No Vaccination at £20,000/QALY | INMB compared to no vaccination at £30,000/QALY |
|---|---|---|---|---|---|---|---|---|---|---|
| No Vaccination counterfactual | 39725 (9933,65949) | 6270 (5912,6739) | 82740 (60755 ,105361) | 9493959 (9493557, 9494298) | – | – | – | Dominated | – | – |
| Pre-emptive vaccination (PV) of GBMSM at high risk for mpox who attend SHS | | | | | | | | | | |
| PV 13 vaccines per day | 14208 (8354,20506) | 5538 (5111,6034) | 71664 (63887, 78945) | 9494219 (9494113, 9494307) | -11077 (-29800,7473) | 260.26 (-4.44,558.73) | Cost-saving | Dominated | 16281841 (-7516403, 38836488) | 18884474 (-7545618, 43847520) |
| PV 27 vaccines per day | 4215 (2571,6046) | 3887 (2840,4619) | 65152 (53978, 73115) | 9494312 (9494273, 9494346) | -17588 (-37617, 3658) | 353.22 (37.33,722.36) | Cost-saving | Dominated | 24652096 (-2403887, 50014713) | 28184322 (-1886713, 56473014) |
| PV 41 vaccines per day | 1371 (121,2777) | 1724 (168,3238) | 58881 (45397, 72494) | 9494327 (9494296, 9494356) | -23859 (-44064, -4514) | 367.68 (44.53,751.26) | Cost-saving | Dominated | 31212910 (6977473, 54274311) | 34889756 (7740281, 60319858) |
| PV 54 vaccines per day | 1291 (68,2733) | 1626 (100,3206) | 58598 (45022, 72626) | 9494327 (9494297, 9494356) | -24142 (-44659, -5239) | 367.90 (45.47,750.90) | Cost-saving | Dominated | 31500281 (7368563, 55066357) | 3517925 (8066895, 60777964) |
| PV 81 vaccines per day | 1257 (61,2737) | 1585 (93,3226) | 58598 (45315, 72889) | 9494327 (9494298, 9494356) | -24142 (-44298, -5418) | 368.17 (46.25,751.92) | Cost-saving | Dominated | 31505276 (7317676, 54734785) | 35186979 (7977633, 61305227) |
| **PV 135 vaccines per day** | **1224 (29,2718)** | **1547 (44,3169)** | **58532 (45511, 73113)** | **9494327 (9494298, 9494356)** | **-24208 (-44689, -5023)** | **368.22 (46.67,752.39)** | **Cost-saving** | **Lowest cost scenario** | **31572106 (6975525, 55072058)** | **35254353 (7652691, 61845953)** |
| Reactive vaccination (RV) of GBMSM at high risk for mpox who attend SHS | | | | | | | | | | |
| RV 27 vaccines per day | 8882 (6287,12105) | 5133 (4823,5511) | 70265 (63972, 75765) | 9494268 (9494201, 9494325) | -12475 (33210,8044) | 309.14 (18,73,644.15) | Cost-saving | Dominated | 18657476 (-7528232, 44617753) | 21748885 (-7352922, 50091728) |
| RV 41 vaccines per day | 5490 (4316,6865) | 4458 (4171,4805) | 68326 (62376, 73839) | 9494301 (9494258, 9494339) | -14414 (-35732, 6268) | 341.76 (31.09,703.35) | Cost-saving | Dominated | 21249432 (-5374454, 48019212) | 24667046 (-4962037, 54632934) |

| | | | | | | | | | | |
|---|---|---|---|---|---|---|---|---|---|---|
| RV 81 vaccines per day | 2609 (2270,3124) | 3020 (2631,3452) | 64030 (55398, 72542) | 9494325 (9494295, 9494355) | -18710 (-38555, -235) | 365.80 (40.16,747.36) | Cost-saving | Dominated | 26026118 (1567584, 51115664) | 29684107 (2306932, 57977203) |
| RV 135 vaccines per day | 2056 (1451,2966) | 2490 (1788,3334) | 63315 (52357, 73516) | 9494326 (9494297, 9494356) | -19425 (-39206, -1454) | 367.47 (43.15,749.48) | Cost-saving | Dominated | 26774372 (2870945, 50885479) | 30449091 (3568056, 57307435) |
| RV 204 vaccines per day | 1928 (1073,2941) | 2354 (1352,3307) | 62994 (50647, 73226) | 9494327 (9494298, 9494356) | -19746 (-39144, -1027) | 367.76 (43.55,750.01) | Cost-saving | Dominated | 27101336 (2752738, 50949351) | 30778917 (3635085, 57627938) |
| RV 465 vaccines per day | 1861 (936,2932) | 2284 (1219,3297) | 62932 (51086, 73569) | 9494327 (9494298, 9494356) | -19808 (-39418, -1227) | 368.17 (44.05,750.65) | Cost-saving | Dominated | 27171199 (2758659, 51294211) | 30852875 (3739672, 58051950) |

Cost-saving scenarios save money and gain QALYs compared to the no vaccination counterfactual scenario. Dominated scenarios have higher mean costs and save fewer QALYs (just comparison of mean) than another scenario. Extended dominated scenarios have a higher mean ICER and save fewer QALYs (just comparison of mean) than another scenario. The scenario in bold is the most cost-effective vaccination scenario – it saves most money and no other scenario that saves more QALYs is cost-effective compared to it; PV135 also has the highest INMB. This indicates that high importation requires high vaccination rate.

Abbreviations: QALY, Quality Adjusted Life Years; ICER, incremental cost-effectiveness ratio; INMB, incremental net monetary benefit; PV, pre-emptive vaccination; RV, reactive vaccination.

Table S22 Cost effectiveness of vaccination under the low outbreak criteria (an outbreak starts with ≥96 cases and ends with ≤48 cases in 3 months)

| Scenario | Total number of infections | Outbreak duration (days) | Total Cost (1000s £) | Total QALYs | Incremental costs compared to no vaccination (1000s £) | Incremental QALY compared to no vaccination | ICER compared to no vaccination | Fully incremental analysis | INMB compared to No Vaccination at £20,000/QALY | INMB compared to no vaccination at £30,000/QALY |
|---|---|---|---|---|---|---|---|---|---|---|
| No Vaccination counterfactual | 43813 (7116,61224) | 5804 (5521,6282) | 80719 (57203 ,98793) | 9493925 (9493641, 9494310) | – | – | – | Dominated | – | – |
| Pre-emptive vaccination (PV) of GBMSM at high risk for mpox who attend SHS | | | | | | | | | | |
| PV 13 vaccines per day | 11833 (7370,16675) | 4708 (4094,5089) | 62267 (52785, 70079) | 9494250 (9494171, 9494319) | -18452 (-31311,1126) | 324.82 (-24.35,557.59) | Cost-saving | Dominated | 24948944 (-1543626, 40570526) | 28197167 (-1752669, 45531363) |
| PV 27 vaccines per day | 2339 (1141,3755) | 2806 (1727,3647) | 54246 (43805, 62442) | 9494340 (9494310, 9494364) | -26474 (-40662,-7138) | 414.27 (33.80, 681.26) | Cost-saving | Dominated | 34758904 (10095353, 52933037) | 38901596 (11046087, 58849956) |
| **PV 41 vaccines per day** | **496 (59,1463)** | **841 (102,2372)** | **50364 (43727, 63292)** | **9494350 (9494329, 9494372)** | **-30355 (-46675, -12329)** | **425.19 (46.27,694.04)** | **Cost-saving** | **Lowest cost scenario** | **38858794 (13407960, 57057780)** | **43110700 (14079501, 63543458)** |
| PV 54 vaccines per day | 465 (53,1416) | 788 (97,2298) | 50641 (44538, 63211) | 9494351 (9494329, 9494372) | -30078 (-46037, -11518) | 425.76 (47.72,695.41) | Cost-saving | £485,965/QALY compared to PV41 | 38593238 (12623496, 57196971) | 42850791 (13296718, 63667716) |
| PV 81 vaccines per day | 447 (0, 1392) | 763 (0,2270) | 50837 (43885, 63364) | 9494351 (9494329, 9494372) | -29882 (-45708, -11238) | 426.07 (48.35,696.33) | Cost-saving | £632,258/QALY compared to PV54 | 38403746 (12288196, 57172775) | 42664478 (12891425, 63747960) |
| PV 135 vaccines per day | 431 (0,1364) | 744 (0,2231) | 51003 (44259, 63615) | 9494352 (9494329, 9494372) | -29716 (-45395, -11110) | 426.28 (48.16,696.81) | Cost-saving | £790,476/QALY compared to PV81 | 38241856 (12714710, 56864203) | 42504622 (13426323, 63439656) |
| Reactive vaccination (RV) of GBMSM at high risk for mpox who attend SHS | | | | | | | | | | |
| RV 27 vaccines per day | 7862 (5600,10524) | 4486 (4173,4762) | 61066 (54214, 67522) | 9494286 (9494233, 9494334) | -19653 (-34175,1572) | 360.69 (-5.96,606.46) | Cost-saving | Dominated | 26866666 (-1639937, 44558665) | 30473587 (-1675591, 49921204) |
| RV 41 vaccines per day | 4515 (3430,5798) | 3873 (3498,4165) | 58813 (51489, 64866) | 9494320 (9494285, 9494351) | -21906 (-36453, -1559) | 394.30 (14.69,652.39) | Cost-saving | Dominated | 29792138 (1738557, 48101352) | 33735089 (1873961, 54712734) |

| | | | | | | | | | | |
|---|---|---|---|---|---|---|---|---|---|---|
| RV 81 vaccines per day | 1863 (1577,2286) | 2669 (2275,3013) | 55806 (47142, 63077) | 9494344 (9494321, 9494367) | -24914 (-39581, -5729) | 419.05 (32.80,689.21) | Cost-saving | Dominated | 33294805 (7269604, 52043310) | 37485343 (7851718, 58345327) |
| RV 135 vaccines per day | 1219 (1019,1674) | 1886 (1534,2591) | 53388 (44393, 65051) | 9494346 (9494324, 9494368) | -27331 (-42610, -9478) | 420.92 (34.22,692.48) | Cost-saving | Dominated | 35749180 (10629733, 54085740) | 39958353 (11056695, 60189284) |
| RV 204 vaccines per day | 1005 (662,1651) | 1621 (1079,2562) | 53441 (42633, 65244) | 9494348 (9494327, 9494368) | -27278 (-42391, -9920) | 422.62 (38.44,692.80) | Cost-saving | Dominated | 35730426 (11327893, 53338425) | 39956654 (12053877, 60148712) |
| RV 465 vaccines per day | 929 (485,1629) | 1510 (778,2517) | 54139 (44500, 65532) | 9494349 (9494328, 9494369) | -26580 (-41942, -8301) | 423.26 (40.48,694.46) | Cost-saving | Dominated | 35045124 (9635016, 52817970) | 39277760 (10575563, 59865523) |

Cost-saving scenarios save money and gain QALYs compared to the no vaccination counterfactual scenario. Dominated scenarios have higher mean costs and save fewer QALYs (just comparison of mean) than another scenario. Extended dominated scenarios have a higher mean ICER and save fewer QALYs (just comparison of mean) than another scenario. The scenario in bold is the most cost-effective vaccination scenario – it saves most money and no other scenario that saves more QALYs is cost-effective compared to it; PV41 also has the highest INMB.

Abbreviations: QALY, Quality Adjusted Life Years; ICER, incremental cost-effectiveness ratio; INMB, incremental net monetary benefit; PV, pre-emptive vaccination; RV, reactive vaccination.

Table S23 Cost effectiveness of vaccination under the high outbreak criteria (an outbreak starts with ≥144 cases and ends with ≤72 cases in 3 months)

| Scenario | Total number of infections | Outbreak duration (days) | Total Cost (1000s £) | Total QALYs | Incremental costs compared to no vaccination (1000s £) | Incremental QALY compared to no vaccination | ICER compared to no vaccination | Fully incremental analysis | INMB compared to No Vaccination at £20,000/QALY | INMB compared to no vaccination at £30,000/QALY |
|---|---|---|---|---|---|---|---|---|---|---|
| No Vaccination counterfactual | 60592 (47166,71777) | 5249 (4852,5476) | 88641 (71803 ,105278) | 9493728 (9493461, 9494004) | – | – | – | Dominated | – | – |
| Pre-emptive vaccination (PV) of GBMSM at high risk for mpox who attend SHS | | | | | | | | | | |
| PV 13 vaccines per day | 14181 (8660,20074) | 3901 (3181,4445) | 57471 (45998, 67683) | 9494208 (9494112, 9494297) | -31169 (-41634,-23100) | 480.65 (291.65,670.96) | Cost-saving | Dominated | 40782500 (30931505, 52873922) | 45589012 (34552425, 58628558) |
| PV 27 vaccines per day | 1987 (369,4075) | 1800 (412,2955) | 46099 (33700, 57631) | 9494323 (9494287, 9494354) | -42541 (-56022,-32461) | 594.84 (347.38, 835.16) | Cost-saving | Dominated | 54438201 (42325896, 69511778) | 60386647 (47167791, 76455628) |
| **PV 41 vaccines per day** | **142 (0,822)** | **177 (0,1081)** | **44967 (42495, 53631)** | **9494341 (9494304, 9494368)** | **-43674 (-58525, -29409)** | **613.33 (364.99,854.74)** | **Cost-saving** | **Lowest cost scenario** | **55940378 (38130487, 72893818)** | **62073685 (42831062, 80948754)** |
| PV 54 vaccines per day | 130 (0,779) | 163 (0,1060) | 45631 (43362, 53941) | 9494343 (9494306, 9494369) | -43010 (-57787, -28526) | 614.74 (365.52,855.79) | Cost-saving | Extendedly dominated | 55304881 (37264374, 72671571) | 61452240 (42017482, 80799160) |
| PV 81 vaccines per day | 117 (0, 745) | 150 (0,998) | 45942 (43851, 53929) | 9494343 (9494308, 9494370) | -42699 (-57671, -28031) | 615.43 (365.86,855.81) | Cost-saving | £464,286/QALY compared to PV41 | 55007798 (36779590, 72504224) | 61162096 (41562088, 80507853) |
| PV 135 vaccines per day | 105 (0,736) | 138 (0,995) | 46182 (44245, 54233) | 9494344 (9494308, 9494370) | -42459 (-57365, -27634) | 615.84 (366.10,856.35) | Cost-saving | £585,366/QALY compared to PV81 | 54775959 (36392215, 72171422) | 60934311 (41190964, 80225804) |
| Reactive vaccination (RV) of GBMSM at high risk for mpox who attend SHS | | | | | | | | | | |
| RV 27 vaccines per day | 11660 (8408,15341) | 3968 (3531,4295) | 57813 (48424, 65717) | 9494229 (9494157, 9494298) | -30828 (-41684,-21383) | 501.42 (295.90,703.60) | Cost-saving | Dominated | 40856158 (28999232, 53939731) | 45870386 (32700298, 60211120) |
| RV 41 vaccines per day | 6612 (4859,8733) | 3433 (3010,3788) | 54437 (46001, 61903) | 9494280 (9494230, 9494327) | -34204 (-46430, -23944) | 552.52 (321.70,775.18) | Cost-saving | Dominated | 45254554 (32248420, 59853333) | 50779743 (36317063, 66865260) |

| | | | | | | | | | | |
|---|---|---|---|---|---|---|---|---|---|---|
| RV 81 vaccines per day | 2451 (1981,3126) | 2374 (1945,2729) | 50583 (41620, 58609) | 9494322 (9494292, 9494353) | -38058 (-51095, -27705) | 593.75 (340.32,833.17) | Cost-saving | Dominated | 49932954 (37334511, 65224981) | 55870495 (41423171, 72763927) |
| RV 135 vaccines per day | 1387 (1183,1635) | 1606 (1295,1968) | 47440 (38272, 57664) | 9494326 (9494296, 9494356) | -41200 (-54284, -30787) | 598.15 (341.26,841.65) | Cost-saving | Dominated | 53163418 (40628137, 68624532) | 59144938 (44773422, 76253640) |
| RV 204 vaccines per day | 995 (809,1474) | 1195 (907,1848) | 46197 (36394, 58261) | 9494327 (9494298, 9494356) | -42444 (-55643, -31694) | 598.99 (344.11,840.76) | Cost-saving | Dominated | 54424024 (41777507, 69844604) | 60413948 (46279619, 77034395) |
| RV 465 vaccines per day | 761 (437,1449) | 933 (497,1823) | 47620 (37936, 58666) | 9494330 (9494303, 9494358) | -41021 (-55054, -29765) | 602.38 (350.50,840.90) | Cost-saving | Dominated | 53068804 (39696339, 68249374) | 59092643 (44670332, 76056564) |

Cost-saving scenarios save money and gain QALYs compared to the no vaccination counterfactual scenario. Dominated scenarios have higher mean costs and save fewer QALYs (just comparison of mean) than another scenario. Extended dominated scenarios have a higher mean ICER and save fewer QALYs (just comparison of mean) than another scenario. The scenario in bold is the most cost-effective vaccination scenario – it saves most money and no other scenario that saves more QALYs is cost-effective compared to it; PV41 also has the highest INMB.

Abbreviations: QALY, Quality Adjusted Life Years; ICER, incremental cost-effectiveness ratio; INMB, incremental net monetary benefit; PV, pre-emptive vaccination; RV, reactive vaccination.

Table S24 Cost effectiveness of vaccination assuming there is no responsive behaviour change upon outbreaks. In view of no responsive behaviour change, vaccination scenarios at two high rates (203 and 271 doses per day) for PV and one high rate (558 doses per day) for RV are added.

| Scenario | Total number of infections | Outbreak duration (days) | Total Cost (1000s £) | Total QALYs | Incremental costs compared to no vaccination (1000s £) | Incremental QALY compared to no vaccination | ICER compared to no vaccination | Fully incremental analysis | INMB compared to No Vaccination at £20,000/QALY | INMB compared to no vaccination at £30,000/QALY |
|---|---|---|---|---|---|---|---|---|---|---|
| No Vaccination counterfactual | 328100 (252055,423779) | 7197 (6909,7281) | 325806 (235073,445005) | 9490571 (9488411,9492344) | – | – | – | Dominated | – | – |
| Pre-emptive vaccination (PV) of GBMSM at high risk for mpox who attend SHS | | | | | | | | | | |
| PV 13 vaccines per day | 184699 (113558,278205) | 7074 (6208,7281) | 218247 (143599, 313687) | 9492314 (9490745, 9493450) | -107558 (-143562,-78576) | 1742.63 (1053.08,2456.44) | Cost-saving | Dominated | 142411006 (108103454, 185057029) | 159837284 (120185362, 207414114) |
| PV 27 vaccines per day | 35415 (7455,99738) | 6319 (5209,7281) | 106508 (73168, 170290) | 9494018 (9493229, 9494326) | -219298 ( -299838,-158290) | 3446.68 (1942.19,5011.82) | Cost-saving | Dominated | 288231463 (213173352, 390113677) | 322698224 (237961089, 436006091) |
| PV 41 vaccines per day | 2673 (0,11411) | 2264 (0,7258) | 63797 (42793, 113151) | 9494328 (9494261, 9494366) | -262008 (-360090,-188198) | 3757.50 (2022.17,5850.84) | Cost-saving | Dominated | 337158361 (247640322, 459125609) | 374733360 (273530949, 509465085) |
| PV 54 vaccines per day | 2414 (0,10031) | 2099 (0,6569) | 62680 (43577, 105950) | 9494332 (9494269, 9494367) | -263126 (-362676,-187821) | 3760.71 (2023.04,5859.88) | Cost-saving | Dominated | 338340035 (247296489, 464681360) | 375947109 (272779581, 514900426) |
| PV 81 vaccines per day | 2230 (0,9601) | 1990 (0,6565) | 61779 (44035, 106093) | 9494334 (9494276, 9494367) | -264027 (-366243,-188151) | 3762.84 (2023.59,5866.21) | Cost-saving | Dominated | 339283786 (247331255, 465971074) | 376912228 (272320199, 515670530) |
| PV 135 vaccines per day | 2108 (0,8963) | 1918 (0,5990) | 61238 (44414, 98748) | 9494335 (9494279, 9494368) | -264568 (-366523,-188641) | 3764.26 (2023.98,5870.31) | Cost-saving | Dominated | 339853438 (247269305, 467125358) | 377496023 (271938361, 519047843) |
| PV 203 vaccines per day | 2047 (0,8717) | 1880 (0,5889) | 60891 (44545, 98253) | 9494336 (9494280, 9494368) | -264915 (-366774,-188694) | 3764.90 (2024.16,5872.12) | Cost-saving | Dominated | 340212519 (247282162, 467196689) | 377861472 (272505676, 519085598) |
| **PV 271 vaccines per day** | **2021 (0,8677)** | **1865 (0,5885)** | **60774 (44620, 98242)** | **9494336 (9494282, 9494368)** | **-265032 (-366820,-188621)** | **3765.22 (2024.25,5873.03)** | **Cost-saving** | **Lowest cost scenario** | **340336449 (247551681, 467215658)** | **377988664 (272435962, 519103572)** |
| Reactive vaccination (RV) of GBMSM at high risk for mpox who attend SHS | | | | | | | | | | |

| | | | | | | | | | | |
|---|---|---|---|---|---|---|---|---|---|---|
| RV 27 vaccines per day | 38194 (10487,100656) | 7052 (6705,7281) | 115103 (88178, 171259) | 9493989 (9493215, 9494305) | -210703 (-296406,-143566) | 3417.55 (1921.08,4994.76) | Cost-saving | Dominated | 279054028 (197335027, 383426241) | 313229554 (223571092, 426570237) |
| RV 41 vaccines per day | 7468 (5745,12095) | 6041 (4732,7259) | 91818 (73715, 113632) | 9494298 (9494238, 9494341) | -233988 (-341506,-157032) | 3727.46 (1996.99,5837.67) | Cost-saving | Dominated | 308536954 (211816711, 441279286) | 345811543 (237921412, 497369259) |
| RV 81 vaccines per day | 4655 (2833,10271) | 4156 (2773,6772) | 76487 (58226, 109052) | 9494313 (9494268, 9494351) | -249319 (-352713,-172609) | 3742.43 (2005.77,5859.09) | Cost-saving | Dominated | 324167493 (229702002, 453641951) | 361591785 (256502940, 508352715) |
| RV 135 vaccines per day | 3954 (1741,9946) | 3594 (1764,6740) | 72280 (51492, 108179) | 9494318 (9494272, 9494354) | -253526 (-354904,-179304) | 3747.31 (2009.36,5862.26) | Cost-saving | Dominated | 328471724 (236269653, 454903135) | 365944774 (262976403, 509329241) |
| RV 204 vaccines per day | 3639 (1288,9652) | 3347 (1330,6463) | 70401 (49428, 105318) | 9494321 (9494274, 9494356) | -255405 (-358426,-181157) | 3749.63 (2011.52,5865.07) | Cost-saving | Dominated | 330397335 (237920747, 456073024) | 367893659 (264828853, 511817326) |
| RV 465 vaccines per day | 3360 (927,9428) | 3191 (974,6555) | 69553 (47232, 105652) | 9494323 (9494277, 9494357) | -256253 (-358592,-183188) | 3751.88 (2012.66,5867.73) | Cost-saving | Dominated | 331290675 (239149717, 456155423) | 368809475 (265320726, 511890223) |
| RV558 vaccines per day | 3323 (894,9409) | 3168 (951,6556) | 69430 (47054, 105650) | 9494323 (9494277, 9494358) | -256376 (-358591,-183102) | 3752.07 (2012.76,5868.08) | Cost-saving | Dominated | 331417095 (239769656, 456136094) | 368937816 (265341196, 511886543) |

Cost-saving scenarios save money and gain QALYs compared to the no vaccination counterfactual scenario. Dominated scenarios have higher mean costs and save fewer QALYs (just comparison of mean) than another scenario. Extended dominated scenarios have a higher mean ICER and save fewer QALYs (just comparison of mean) than another scenario. The scenario in bold is the most cost-effective vaccination scenario – it saves most money and no other scenario that saves more QALYs is cost-effective compared to it; PV 271 has the highest INMB. It worth noting that without responsive behaviour change and up to vaccination rate of 271 doses per day (i.e., 100% coverage after one year vaccination), increasing vaccination rates are better.

Abbreviations: QALY, Quality Adjusted Life Years; ICER, incremental cost-effectiveness ratio; INMB, incremental net monetary benefit; PV, pre-emptive vaccination; RV, reactive vaccination.

**Table S25** Estimates of model parameters based on calibration of the transmission model to case data of the whole GBMSM population (considering 3023 male cases among which 148 cases were imported) up to day 118 (12 August 2022) and day 267 (10 January 2023) using physical sex contact data from the 2022 RIISH-MPOX survey, with a cut-off of 10 sexual contacts in the last 4 months to define the high risk GBMSM. The risk behaviours of GBMSM decrease in the outbreak and then revert to pre-outbreak level from 16 November 2022. The high-risk proportion includes 23.3% of GBMSM with the respective mean contact rates of 3.7 and 25.6 in the last 4 months for low and high-risk groups, respectively.

| Model parameter | Prior | Posterior; point estimates are mean and range in brackets is 95% credibility interval | |
|---|---|---|---|
| | | Calibration up to 12 August 2022 | Calibration up to 10 January 2023 |
| Initial growth rate ($\psi_r$) | U[0.01,0.15] | 0.059 [0.038,0.074] | 0.017 [0.014,0.023] |
| Transmission coefficients ($\beta$) | – | 0.58[0.43,0.71] | 0.41 [0.36,0.44] |
| $R_0$ (early stage) | – | 1.58 [1.47,1.84] | 1.26 [1.20,1.35] |
| $R_0$ after $t_c$ | – | 0.85 [0.77,0.94] | 0.82[0.79,0.83] |
| Time $t_c$ after which risk behaviour and infectious period changes (days from 17/04/2022) | U[40,80] | 47.41 [40.3,67.54] | 66.02 [59.04,69.85] |
| Reduction in contact rate after tc (1-*w*) | U[0.00,0.70] | 0.447 [0.300,0.551] | 0.07 [0.00,0.20] |
| Infectious period before $t_c$ ($D_1$) (days) | U[2.0,21.0] | 3.05 [2.56,4.31] | 3.09 [3.00,3.52] |
| Infectious period after $t_c$ ($D_2$) (days) | U[1.5,10.0] | 2.31 [1.95,3.17] | 2.18 [1.94, 2.53] |
| *C* (parameter defining the rate of change in behaviour and effective infectious period after $t_c$) | U[0.01,2.5] | 0.017 [0.012,0.037] | 0.090 [0.061,0.146] |

U[a,b] refers to uniform distribution with minimum value a and maximum value b

**Table S26** Cost effectiveness of vaccination based on the transmission model that uses 2022 RiiSH physical contact to construct contact matrix

| Scenario | Total number of infections | Outbreak duration (days) | Total Cost (1000s £) | Total QALYs | Incremental costs compared to no vaccination (1000s £) | Incremental QALY compared to no vaccination | ICER compared to no vaccination | Fully incremental analysis | INMB compared to No Vaccination at £20,000/QALY | INMB compared to no vaccination at £30,000/QALY |
|---|---|---|---|---|---|---|---|---|---|---|
| No Vaccination counterfactual | 13803 (9471,19659) | 6081 (5858,6322) | 61591 (56368 ,67436) | 9494230 (9494133, 9494308) | – | – | – | Dominated | – | – |
| Pre-emptive vaccination (PV) of GBMSM at high risk for mpox who attend SHS | | | | | | | | | | |
| PV 13 vaccines per day | 9184 (7093,12229) | 4885 (4453,5269) | 62779 (56169, 69384) | 9494267 (9494200, 9494325) | 1188 (-2298,4213) | 37.38 (3.84,84.22) | Not cost-saving | Dominated | -440642 (-4043974, 3115011) | -66817 (-3965202, 3530077) |
| PV 27 vaccines per day | 3667 (2271,5348) | 3249 (2223,4100) | 59651 (49502, 68665) | 9494315 (9494278, 9494349) | -1940 (-9884, 4301) | 85.19 (35.36, 155.88) | Cost-saving | Dominated | 3644212 (-2543030, 11203456) | 4496086 (-1902910, 11787941) |
| **PV 41 vaccines per day** | 815 (170,2055) | 867 (155,2100) | **53241 (46848, 64827)** | **9494331 (9494302, 9494361)** | **-8351 (-15390, 517)** | **100.63 (48.58,176.53)** | **Cost-saving** | **Lowest cost scenario** | **10363334 (1556804, 17572198)** | **11369683 (2578168, 18727244)** |
| PV 54 vaccines per day | 270 (165,521) | 232 (153,448) | 61292 (59992, 64209) | 9494346 (9494319, 9494370) | -300 (-4847, 4282) | 115.91 (59.87,194.48) | Cost-saving | £602,394/QALY compared to RV465 | 2617765 (-2620416, 7996819) | 3776868 (-1750782, 9722202) |
| PV 81 vaccines per day | 213 (156, 390) | 175 (150,323) | 86170 (85628, 88144) | 9494361 (9494343, 9494378) | 24579 (19895, 29313) | 131.08 (69.87,216.69) | Not cost-saving | £1640,013/QALY compared to PV54 | -21957026 (-27692707,-15988499) | -20646225 (-26727694, -13895321) |
| PV 135 vaccines per day | 188 (136,248) | 159 (142,174) | 94472 (94093, 94912) | 9494365 (9494349, 9494381) | 32881 (27348, 37747) | 135.45 (72.23,222.01) | Not cost-saving | £1899,771/QALY compared to PV81 | -30171610 (-36093456,-23661908) | -28817125 (-35102783, -21706980) |
| Reactive vaccination (RV) of GBMSM at high risk for mpox who attend SHS | | | | | | | | | | |
| RV 27 vaccines per day | 7405 (6134,9438) | 4671 (4369,4973) | 63587 (57718, 70062) | 9494284 (9494231, 9494332) | 1995 (-967,5170) | 54.11 (14.50,107.33) | Not cost-saving | Dominated | -913135 (-4577799, 568715) | -371994 (-4145758, 3467734) |
| RV 41 vaccines per day | 55220 (4369,6494) | 4084 (3716,4427) | 62940 (56414, 69607) | 9494304 (9494266, 9494343) | 1349 (-2844, 5253) | 74.21 (26.52,138.36) | Not cost-saving | Dominated | 135489 (-3994462, 4413825) | 877540 (-3339181, 5328978) |

| | | | | | | | | | | |
|---|---|---|---|---|---|---|---|---|---|---|
| RV 81 vaccines per day | 2915 (2496,3376) | 2757 (2359,3239) | 59349 (50941, 69635) | 9494319 (9494289, 9494350) | -2242 (-8119, 4188) | 89.24 (34.37,167.29) | Cost-saving | Dominated | 4026765 (-2101198, 9657286) | 4919210 (-1162370, 10542971) |
| RV 135 vaccines per day | 1913 (1696,2222) | 1900 (1727,2140) | 56564 (50769, 64347) | 9494323 (9494294, 9494354) | -5027 (-10410, -936) | 93.52 (39.98,172.34) | Cost-saving | Dominated | 6897425 (1435927, 12260859) | 7832655 (2296586, 13332953) |
| RV 204 vaccines per day | 1336 (1167,1614) | 1375 (1248,1661) | 55522 (50286, 66108) | 9494329 (9494300, 9494357) | -6069 (-11167, 1479) | 98.58 (44.06,176.87) | Cost-saving | Dominated | 8040576 (1414546, 13500715) | 9026333 (2564600, 14597058) |
| RV 465 vaccines per day | 688 (590,792) | 693 (621,775) | 57015 (51410, 63286) | 9494339 (9494314, 9494364) | -4577 (-9871, 683) | 108.81 (52.23,188.88) | Cost-saving | £461,369/QALY compared to PV41 | 6752868 (1467659, 12373263) | 7841013 (2407424, 13676640) |

Cost-saving scenarios save money and gain QALYs compared to the no vaccination counterfactual scenario. Dominated scenarios have higher mean costs and save fewer QALYs (just comparison of mean) than another scenario. Extended dominated scenarios have a higher mean ICER and save fewer QALYs (just comparison of mean) than another scenario. The scenario in bold is the most cost-effective vaccination scenario – it saves most money and no other scenario that saves more QALYs is cost-effective compared to it; PV41 also has the highest INMB.

Abbreviations: QALY, Quality Adjusted Life Years; ICER, incremental cost-effectiveness ratio; INMB, incremental net monetary benefit; PV, pre-emptive vaccination; RV, reactive vaccination.

**Table S27** Cost effectiveness of vaccination in the situation where breakthrough infections have half the chance of experiencing moderate and severe mpox disease compared to the baseline scenarios.

| Scenario | Total number of infections | Outbreak duration (days) | Total Cost (1000s £) | Total QALYs | Incremental costs compared to no vaccination (1000s £) | Incremental QALY compared to no vaccination | ICER compared to no vaccination | Fully incremental analysis | INMB compared to No Vaccination at £20,000/QALY | INMB compared to no vaccination at £30,000/QALY |
|---|---|---|---|---|---|---|---|---|---|---|
| No Vaccination counterfactual | 54891 (26246,67256) | 5472 (5141,5676) | 86065 (67633, 101969) | 9493801 (9493545, 9494086) | – | – | – | Dominated | – | – |
| Pre-emptive vaccination (PV) of GBMSM at high risk for mpox who attend SHS | | | | | | | | | | |
| PV 13 vaccines per day | 13099 (8224,18452) | 4219 (3494,4673) | 59116 (48230, 68299) | 9494231 (9494145, 9494308) | -26949 (-37751,-15237) | 429.75 (182.94,614.41) | Cost-saving | Dominated | 35543870 (17867415, 48126309) | 39841339 (19401435, 53759254) |
| PV 27 vaccines per day | 2143 (637,3860) | 2210 (802,3198) | 49271 (36700, 59268) | 9494333 (9494303, 9494359) | -36794 (-49957, -26184) | 532.03 (259.06,766.50) | Cost-saving | Dominated | 47434553 (32688165, 62871192) | 52754881 (35041654, 69268528) |
| **PV 41 vaccines per day** | **254 (0,1143)** | **369 (0,1719)** | **46488 (42485, 58035)** | **9494346 (9494320, 9494370)** | **-39577 (-54334, -24465)** | **545.26 (277.83, 779.06)** | **Cost-saving** | **Lowest cost scenario** | **50481845 (30113639, 67202994)** | **55934446 (33172011, 74064497)** |
| PV 54 vaccines per day | 234 (0,1101) | 340 (0,1641) | 47015 (43349, 58208) | 9494347 (9494321, 9494371) | -39049 (-53527,-24059) | 546.14 (278.76,779.05) | Cost-saving | Extendedly dominated | 49972292 (29250271, 66466143) | 55433738 (32318112, 73287505) |
| PV 81 vaccines per day | 221 (0,1064) | 325 (0,1594) | 47310 (43833, 58478) | 9494348 (9494321, 9494370) | -38755 (-53126,-23608) | 546.69 (279.35,780.39) | Cost-saving | £574,825/QALY compared to PV41 | 49688804 (29155059, 66039547) | 55155680 (31839685, 73234353) |
| PV 135 vaccines per day | 203 (0,1022) | 305 (0,1523) | 47453 (44226, 58265) | 9494348 (9494322, 9494371) | -38612 (-53340,-23260) | 546.93 (279.75, 780.98) | Cost-saving | £595,833/QALY compared to PV81 | 49550693 (28775445, 66064748) | 55020042 (31455462, 73391493) |
| Reactive vaccination (RV) of GBMSM at high risk for mpox who attend SHS | | | | | | | | | | |
| RV 27 vaccines per day | 9841 (7021,13064) | 4164 (3745,4450) | 58651 (50190, 65579) | 9494259 (9494198, 9494316) | -27414 (-38690,-13753) | 458.49 (203.07,658.04) | Cost-saving | Dominated | 36583822 (17541190, 50918377) | 41168724 (19579907, 56346333) |
| RV 41 vaccines per day | 5597 (4074,7416) | 3596 (3171,3909) | 55765 (47790, 62507) | 9494302 (9494260, 9494340) | -30300 (-42225,-16535) | 500.65 (232.04,721.40) | Cost-saving | Dominated | 40312678 (20712592, 55310936) | 45319168 (23233789, 61951131) |

| | | | | | | | | | | |
|---|---|---|---|---|---|---|---|---|---|---|
| RV 81 vaccines per day | 2157 (1747,2722) | 2487 (2060,2842) | 52441 (43504, 60315) | 9494335 (9494309, 9494361) | -33624 (-45916,-21849) | 533.68 (253.66,771.68) | Cost-saving | Dominated | 44297800 (26709190, 59542583) | 49634611 (29281661, 66745489) |
| RV 135 vaccines per day | 1284 (1071,1605) | 1699 (1370,2233) | 49514 (40143, 60372) | 9494338 (9494313, 9494363) | -36551 (-49518,-24540) | 536.84 (256.10,778.20) | Cost-saving | Dominated | 47287361 (30164050, 62589746) | 52655736 (32892715, 69810681) |
| RV 204 vaccines per day | 960 (729,1565) | 1328 (966,2148) | 48871 (38419, 61156) | 9494339 (9494317, 9494363) | -37194 (-50707,-25741) | 538.40 (259.22,776.71) | Cost-saving | Dominated | 47962077 (32640630, 62431864) | 53346067 (35296081, 70150567) |
| RV 465 vaccines per day | 811 (462,1530) | 1130 (630,2111) | 49961 (41513, 61394) | 9494340 (9494318, 9494365) | -36104 (-49970,-23823) | 539.51 (261.89,777.55) | Cost-saving | Dominated | 46894311 (30129109, 62080141) | 52289420 (33115402, 68735161) |

Cost-saving scenarios save money and gain QALYs compared to the no vaccination counterfactual scenario. Dominated scenarios have higher mean costs and save fewer QALYs (just comparison of mean) than another scenario. Extended dominated scenarios have a higher mean ICER and save fewer QALYs (just comparison of mean) than another scenario. The scenario in bold is the most cost-effective vaccination scenario – it saves most money and no other scenario that saves more QALYs is cost-effective compared to it; PV41 also has the highest INMB.

Abbreviations: QALY, Quality Adjusted Life Years; ICER, incremental cost-effectiveness ratio; INMB, incremental net monetary benefit; PV, pre-emptive vaccination; RV, reactive vaccination.

Table S28 Cost effectiveness of vaccination in the situation where 50% of vaccine doses are given as fractional quarter doses and half as full doses. We assume the vaccine effectiveness of ¼ fractional dose is reduced by 25% compared to full dose. Point values are means and ranges are 95% CrI.

| Scenario | Total number of infections | Outbreak duration (days) | Total Cost (1000s £) | Total QALYs ( QALY) | Incremental costs compared to no vaccination (1000s £) | Incremental QALY compared to no vaccination | ICER compared to no vaccination | Fully incremental analysis | Incremental NB compared to No Vaccination ( £20,000/QALY) | Incremental Net Benefit compared to no vaccination at £30,000/QALY |
|---|---|---|---|---|---|---|---|---|---|---|
| No Vaccination counterfactual | 54891 (26246,67256 ) | 5472 (5141,5676) | 86127 (67671, 102039) | 9493798 (9493540, 9494085) | – | – | – | Dominated | – | – |
| Pre-emptive vaccination (PV) of GBMSM at high risk for mpox who attend SHS | | | | | | | | | | |
| PV 13 vaccines per day | 17779 (11830,24060 ) | 4727 (4245,5050) | 63125 (53356,72280) | 9494179 (9494067, 9494281) | -23003 (-33080, -8828) | 380.71 (146.26,545.98) | Cost-saving | Dominated | 30616950 (11401508, 42118427) | 34424051 (12631478, 47016147) |
| PV 27 vaccines per day | 3965 (1815,6356) | 3148 (1986,3812) | 50219 (38610,58155) | 9494316 (9494274, 9494351) | -35909 (-47931, -24363) | 517.26 (250.37,739.96) | Cost-saving | Dominated | 46253812 (30347710, 61066876) | 51426405 (32630099, 67516089) |
| PV 41 vaccines per day | 1101 (98,2622) | 1411 (115,2784) | 43260 (31658,55609) | 9494336 (9494310, 9494362) | -42868 (-56997,-31356) | 537.62 (263.12,775.78) | Cost-saving | Dominated | 53620065 (39793945, 68848246) | 58996257 (42912951, 75410953) |
| PV 54 vaccines per day | 1039 (92,2525) | 1337 (113,2745) | 43012 (32172,55278) | 9494336 (9494311, 9494363) | -43115 (-57633,-31296) | 538.10 (264.62,776.90) | Cost-saving | Dominated | 53877412 (39398311, 68990184) | 59258453 (42455361, 75572607) |
| PV 81 vaccines per day | 1008 (80,2501) | 1304 (106,2716) | 42958 (32395,55441) | 9494337 (9494312, 9494363) | -43170 (-57572,-30982) | 538.42 (265.42,776.93) | Cost-saving | Dominated | 53938397 (39142793, 69232762) | 59322637 (42251437, 75848791) |
| PV 135 vaccines per day | 978 (0,2474) | 1276 (0,2699) | 42891 (31484,55529) | 9494337 (9494311, 9494363) | -43236 (-57411, -30975) | 538.45 (265.88,777.25) | Cost-saving | Dominated | 54005398 (39162164, 69397171) | 59389888 (42711659, 75857726) |
| PV203 vaccines per day | 965 (0,2461) | 1268 (0,2692) | 42894 (31524,55583) | 9494337 (9494312, 9494363) | -43234 (-57388,-30940) | 538.61 (265.76,777.52) | Cost-saving | Dominated | 54005809 (39085426, 69374364) | 59391925 (42964525, 75819007) |

| | | | | | | | | | | |
|---|---|---|---|---|---|---|---|---|---|---|
| **271 vaccines per day** | **955 (0,2454)** | **1262 (0,2692)** | **42864 (31562,55608)** | **9494337 (9494312, 9494363)** | **-43265 (-57394, -30917)** | **538.70 (265.49,777.65)** | **Cost-saving** | **Lowest cost scenario** | **54038929 (39396727, 69433225)** | **59425931 (42930056, 75820041)** |
| Reactive vaccination (RV) of GBMSM at high risk for mpox who attend SHS | | | | | | | | | | |
| RV 27 vaccines per day | 12343 (8623,16297) | 4499 (4201,4757) | 59937 (52223,67082) | 9494230 (9494151, 9494303) | -26190 (-37191,-10971) | 431.56 (187.38,617.37) | Cost-saving | Dominated | 34821474 (14605790, 48054824) | 39137046 (16398449, 53504351) |
| RV 41 vaccines per day | 7077 (5148,9405) | 3939 (3594,4214) | 55359 (48383,61352) | 9494284 (9494232, 9494330) | -30768 (-3102,-16013) | 485.70 (225.06,696.79) | Cost-saving | Dominated | 40482292 (20303303, 55887172) | 45339323 (22726416, 62087858) |
| RV 81 vaccines per day | 2695 (2113,3461) | 2814 (2421,3143) | 49871 (42439,55782) | 9494328 (9494299, 9494357) | -36256 (-8910,-22961) | 529.95 (252.69,765.60) | Cost-saving | Dominated | 46855171 (27726609, 62927540) | 52154679 (30161202, 69856545) |
| RV 135 vaccines per day | 1806 (1338,2996) | 2154 (1598,2959) | 47300 (37768,56490) | 9494334 (9494306, 9494360) | -38828 (-1592,-27850) | 535.12 (253.91,773.74) | Cost-saving | Dominated | 49530103 (33429690, 64390600) | 54881329 (35799223, 71229770) |
| RV 204 vaccines per day | 1638 (906,2939) | 1988 (1128,2938) | 46917 (35209,56646) | 9494334 (9494308, 9494361) | -39211 (-2085,-28298) | 535.93 (257.71,775.14) | Cost-saving | Dominated | 49929209 (35454419, 64448517) | 55288485 (38391265, 71388534) |
| RV 465 vaccines per day | 1565 (693,2922) | 1911 (876,2914) | 46866 (34959,56723) | 9494335 (9494309, 9494361) | -39261 (-2273,-27979) | 536.29 (257.81,774.68) | Cost-saving | Dominated | 49987037 (35195462, 64939233) | 55349972 (37885119, 71466010) |
| RV 588 vaccines per day | 1559 (685,2917) | 1907 (881,2912) | 46870 (34972,56722) | 9494335 (9494309, 9494361) | -39257 (-2426,-27979) | 536.36 (257.84,774.74) | Cost-saving | Dominated | 49984303 (35179720, 64954657) | 55347918 (37917297, 71459906) |

Cost-saving scenarios save money and gain QALYs compared to the no vaccination counterfactual scenario. Dominated scenarios have higher mean costs and save fewer QALYs (just comparison of mean) than another scenario. Extended dominated scenarios have a higher mean ICER and save fewer QALYs (just comparison of mean) than another scenario. The scenario in bold is the most cost-effective vaccination scenario – it saves most money and no other scenario that saves more QALYs is cost-effective compared to it; PV271 also has the highest INMB.
Abbreviations: QALY, Quality Adjusted Life Years; ICER, incremental cost-effectiveness ratio; INMB, incremental net monetary benefit; PV, pre-emptive vaccination; RV, reactive vaccination.

We assume 50% using full dose and 50% using 1/4 fractional dose vial of vaccine will be delivered to HR GBMSM attending SHS. It is further assumed that those using ¼ dose for 1st dose will use ¼ dose for their 2nd dose and the same for those using full dose. Price for full dose =£160 and Price of 1/4 dose vial =160/4=£40, with same administration costs. VE1= 0.78(0.54-0.89) and VE2= 0.89(0.78-1.00) for full dose and VE(1/4 dose)=0.75*VE(full dose) (i.e., reducing 25%). We assume the same duration of protection - 5 years for 1 dose and 10 years for 2 doses.

### References

1. World Health Organisation. Monkeypox - United Kingdom of Great Britain and Northern Ireland (https://www.who.int/emergencies/disease-outbreak-news/item/2022-DON383) 2022 [
2. Wearing HJ, Rohani P, Keeling MJ. Appropriate models for the management of infectious diseases. PLoS Med. 2005;2(7):e174.
3. Office of National Statistics. Sexual orientation in the UK from 2012 to 2020 by region. (https://www.ons.gov.uk/datasets/sexual-orientation-by-region/editions/time-series/versions/2) 2022 [
4. Brown JR, Reid D, Howarth AR, Mohammed H, Saunders J, Pulford CV, et al. Sexual behaviour, STI and HIV testing and testing need among gay, bisexual and other men who have sex with men recruited for online surveys pre/post-COVID-19 restrictions in the UK. Sex Transm Infect. 2023.
5. Bertran M, Andrews N, Davison C, Dugbazah B, Boateng J, Lunt R, et al. Effectiveness of one dose of MVA-BN smallpox vaccine against mpox in England using the case-coverage method: an observational study. Lancet Infect Dis. 2023.
6. Dugbazah B, Mindlin M. Monkeypox hospitalisation interim report. 2023.
7. Zhang XS, Mandal S, Mohammed H, Turner C, Florence I, Walker J, et al. Transmission dynamics and effect of control measures on the 2022 outbreak of mpox among gay, bisexual, and other men who have sex with men in England: a mathematical modelling study. Lancet Infect Dis. 2024;24(1):65-74.
8. Ogaz D, Enayat Q, Brown JRG, Phillips D, Wilkie R, Jayes D, et al. Mpox Diagnosis, Behavioral Risk Modification, and Vaccination Uptake among Gay, Bisexual, and Other Men Who Have Sex with Men, United Kingdom, 2022. Emerg Infect Dis. 2024;30(5):916-25.
9. National Health Service England (NHS-England). Vaccinations: Mpox (https://www.england.nhs.uk/statistics/statistical-work-areas/vaccinations-for-mpox/) 2024 [
10. Centers for Disease Control and Prevention. Potential Risk for New Mpox Cases (https://emergency.cdc.gov/han/2023/han00490.asp#print). 2023.
11. Sante publique France. Monkey po Aelion: situation point in France on March 23, 2023 (https://www.santepubliquefrance.fr/les-actualites/2023/variole-du-singe-point-de-situation-en-france-au-23-mars-2023). 2023.
12. Charles H, Thorley K, Turner C, Bennet K, Andrews N, Bertran M, et al. Post-peak mpox in England: epidemiology, reinfection, and vaccine effectiveness – data from 2023 (). medRxiv (https://doiorg/101101/20240226243033362). 2024.
13. Berry MT, Khan SR, Schlub TE, Notaras A, Kunasekaran M, Grulich AE, et al. Predicting vaccine effectiveness for mpox. Nat Commun. 2024;15(1):3856.
14. Department of Health and Social Care. Cost-effectiveness methodology for vaccination programmes (https://assets.publishing.service.gov.uk/media/5afd3c17ed915d0de80ffd6c/cemipp-consultation-document.pdf). 2018.
15. National Institute for Health and Care Excellence (NICE). NICE health technology evaluations: the manual (https://www.nice.org.uk/process/pmg36/chapter/economic-evaluation-2#discounting). 2023.

16. National Institute for Health and Care Excellence (NICE). British National Formulary (BNF; https://bnf.nice.org.uk/drugs/) 2023 [
17. Phillips D. Tariff Calculator for Monkeypox activities in Sexual Health Services using the Integrated Sexual Health Tariff (ISHT) Methodology. 2022.
18. National Health Service England (NHS-England). National Cost Collection: National Schedule of NHS Costs - Year 2021/22 - NHS trusts and NHS foundation trusts 2023 [
19. Jones K, Weatherly H, Birch S, Castelli A, Chalkley M, Dargan A. Unit Costs of Health and Social Care 2022 Manual. Technical report. (doi:10.22024/UniKent/01.02.100519). Personal Social Services Research Unit (University of Kent); Centre for Health Economics (University of York) K, UK., editor2022.
20. Office of National Statistics. Employee earnings in the UK: 2022 (https://www.ons.gov.uk/employmentandlabourmarket/peopleinwork/earningsandworkinghours/bulletins/annualsurveyofhoursandearnings/2022) London, UK2022 [
21. Office of National Statistics. Employment in the UK and employee earnings (https://www.ons.gov.uk/employmentandlabourmarket/peopleinwork/employmentandemployeetypes/bulletins/employmentintheuk/may2022#:~:text=The%20UK%20employment%20rate%20was,December%202019%20to%20February%202020).) 2022 [
22. Uk Health Security Agency. HCID status of mpox (monkeypox) (https://www.gov.uk/guidance/hcid-status-of-monkeypox) 2023 [
23. Uk Health Security Agency. Mpox (monkeypox): diagnostic testing (https://www.gov.uk/guidance/monkeypox-diagnostic-testing) 2024 [
24. UK Health Security Agency. Investigation into monkeypox outbreak in England: technical briefing 8 (https://www.gov.uk/government/publications/monkeypox-outbreak-technical-briefings) https://www.gov.uk/government/publications/monkeypox-outbreak-technical-briefings 2022 [
25. National Health Service (NHS). Chief Medical Officer Alert: Tecovirimat as a Treatment for Patients Hospitalised due to Monkeypox Viral Infection (https://www.google.com/url?sa=t&source=web&rct=j&opi=89978449&url=https://www.cas.mhra.gov.uk/ViewandAcknowledgment/ViewAttachment.aspx%3FAttachment_id%3D104081&ved=2ahUKEwiDtKWF1OWGAxVoQUEAHcs3DaQQFnoECA8QAQ&usg=AOvVaw1BozWk0DGCQbiRkvEHHFi4 2022 [
26. National Health Service England (NHS-England). NHS Drug Tariff (https://www.drugtariff.nhsbsa.nhs.uk/#/00837338-DC/DD00837329/Home) 2023 [
27. Fink DL, Callaby H, Luintel A, Beynon W, Bond H, Lim EY, et al. Clinical features and management of individuals admitted to hospital with monkeypox and associated complications across the UK: a retrospective cohort study. Lancet Infect Dis. 2022.
28. Global Burden of Disease Forecasting Collaborators. Burden of disease scenarios for 204 countries and territories, 2022-2050: a forecasting analysis for the Global Burden of Disease Study 2021. Lancet. 2024;403(10440):2204-56.
29. Gater A, Abetz-Webb L, Carroll S, Mannan A, Serpell M, Johnson R. Burden of herpes zoster in the UK: findings from the zoster quality of life (ZQOL) study. BMC Infect Dis. 2014;14:402.
30. Xu M, Liu C, Du Z, Bai Y, Wang Z, Gao C. Real-world effectiveness of mpox (monkeypox) vaccines: a systematic review. J Travel Med. 2023.

31. Hazra A, Zucker J, Bell E, Flores J, Gordon L, Mitja O, et al. Mpox in people with past infection or a complete vaccination course: a global case series. Lancet Infect Dis. 2024;24(1):57-64.
32. Dalton AF, Diallo AO, Chard AN, Moulia DL, Deputy NP, Fothergill A, et al. Estimated Effectiveness of JYNNEOS Vaccine in Preventing Mpox: A Multijurisdictional Case-Control Study - United States, August 19, 2022-March 31, 2023. MMWR Morb Mortal Wkly Rep. 2023;72(20):553-8.
